# Quantifying antibody dynamics of severe and non-severe patients with COVID-19

**DOI:** 10.1101/2022.02.23.22271403

**Authors:** Fernanda Ordoñez-Jiménez, Rodolfo Blanco-Rodríguez, Alexis Erich S. Almocera, Gustavo Chinney-Herrera, Esteban Hernández-Vargas

**Affiliations:** Instituto de Matemáticas, Universidad Nacional Autónoma de México, Boulevard Juriquilla 3001, Querétaro, Qro., 76230, México; Division of Physical Sciences and Mathematics, College of Arts and Sciences, University of the Philippines Visayas, Philippines; Frankfurt Institute for Advanced Studies, Frankfurt am Main, Germany

**Keywords:** Topological Data Analysis, Immune Responses, Mathematical Modeling, SARS-CoV-2, COVID-19

## Abstract

COVID-19 pandemic is a major public health threat with unanswered questions regarding the role of the immune system in the severity level of the disease. In this paper, based on antibody kinetic data of patients with different disease severity, topological data analysis highlights clear differences in the shape of antibody dynamics between three groups of patients, which were non-severe, severe, and one intermediate case of severity. Subsequently, different mathematical models were developed to quantify the dynamics between the different severity groups. The best model was the one with the lowest media value of Akaike Information Criterion for all groups of patients. Although it has been reported high IgG level in severe patients, our findings suggest that IgG antibodies in severe patients may be less effective than non-severe patients due to early B cell production and early activation of the seroconversion process from IgM to IgG antibody.

## 1. INTRODUCTION

Acute Respiratory Syndrome Coronavirus 2 (SARS-CoV-2) is the causative agent of one of the largest pandemic in history. COVID-19 pandemic has resulted in about 5 million of deaths and more than 365 million of infected people. Previous coronavirus outbreaks SARS-CoV (2003) and MERS-CoV (2012) showed similar qualitative and quantitative clinical aspects to SARS-CoV-2. In particular, patients with MERS-CoV [29] had viral levels peak in the second week with a median value of 7.21 (log10 copies/mL) in the severe patient group, and approximately 5.54 (log10 copies/mL) in the mild group. On the other hand, in patients with SARS were reported that the virus peaked at 5.7 (log10 copies/mL) between 7 to 10 days after onset of symptoms (dpso) [33].

Several multifactorial and complex mechanisms are implicated during the course of COVID-19, which consequently lead to the nature of pathogenesis. COVID-19 patients can be classified into mild, moderate, severe, and critical cases. In patients with SARS-CoV-2 the viral peak was approximately 8.85 (log10 copies/mL) around 5 dpso [49], and viral load persisted for 12 days after onset [20]. Severe disease cases reported a mean viral load on admission 60 times higher than the mean of mild disease cases [20]. Remarkably, prolonged viral shedding was presented in severe patients than that of non-severe patients [45].

The immune system has been pointed out by the scientific community as a key factor between severe and non-severe patients Figure1. Among the immune cells recruited to tackle SARS-CoV-2 in the lungs are virus-specific T cells *e*.*g* CD4+ T and CD8+ T cells. The study by Oja *et al*. [30] highlighted that B and T cell responses of critical patients are imbalanced, exhibiting high titers and low virus-specific CD4+ T cell responses. Interestingly, the CD4+ T cell response was impaired as well as the functionality [30], that is SARS-CoV-2-specific CD4+ T cells showed decreased production of Interferon-*γ* (IFN-*γ*), Interleukin 4 (IL-4) and Interleukin 21 (IL-21). While other studies [47] has shown high levels IL-2, IL-10 and IL-6, COVID-19 patients are less inflamed than influenza patients [25].

**Figure 1:**
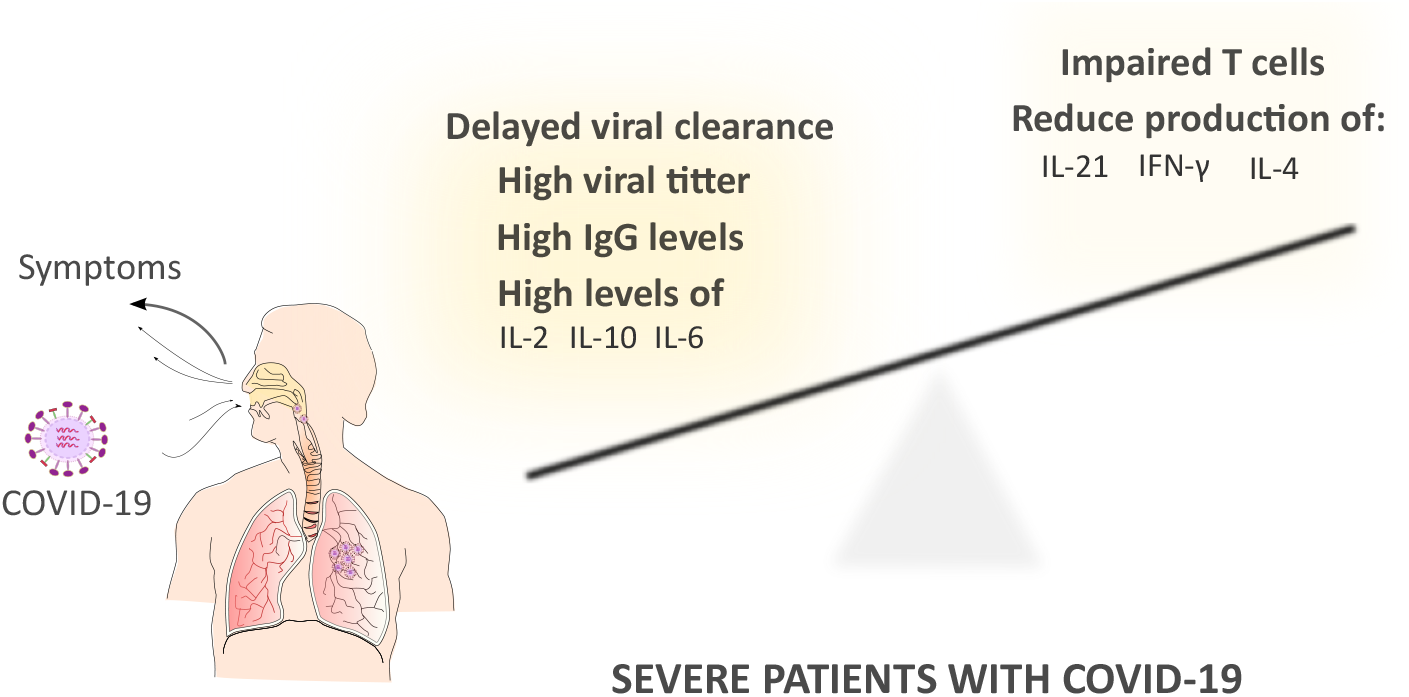
Immune responses associated for the poor clinical outcome in severe COVID-19 patients.

B cell responses could be indicative of a deregulated immune response in severe COVID-19 patients. Timing is central as the ratio between viral loads and antibody titers during the early phase of disease may be predictive for disease severity [30]. Prospective cohort studies with COVID-19 patients highlight that immunoglobulin M (IgM) started on day 7 and peaked on day 28, while that of immunoglobulin G (IgG) was on day 10 and day 49 after illness onset [45, 30]. Strikingly, these studies revealed that IgG antibody are significantly higher in severe patients than non-severe patients [45].

Dissecting the contributions of the identified players in antibody dynamics to severe and non-severe patients with COVID-19 is crucial to develop prophylactic and therapeutic strategies. In this regard, it is important to understand the relationship between data taken from experimental samples of patients with COVID-19 and the severity of the disease. Topological Data Analysis (TDA) is a set of tools based on algebraic topology and whose objective is to retrieve information from high-dimensional databases which are complex to study with traditional statistical methods. The particular tool we used from TDA is the Mapper algorithm. The algorithm was proposed by Singh et al. [41] and has since gained relevance in recent years as it offers graphs with clear interpretations that represent a general map of the phenomenon being studied, simplifying and preserving important features of the data. This algorithm has proven its usefulness in various situations such as the study of breast cancer data [28] or image processing [38].

On the other hand, mathematical modeling has made valuable contributions to our quantitative understanding of different viral infection such as for Influenza [4, 24, 42, 32, 14, 8, 17], HIV [39, 34, 15, 35], Hepatitis[37, 13], Ebola [26, 27], among many others. For COVID-19, there are a few models at within-host level to quantify SARS-CoV-2 dynamics in the host [9, 10, 11, 12, 16, 1, 2, 5]. However, untangling the contributions of different mechanisms by which changes in the immune response in non-severe and severe patients in a temporal manner has not been proposed until now. Therefore, by combining the results of different data sets, topological data analysis and mathematical modelling approaches, the present study aims at clarifying the relative contributions of antibodies between severe and non-severe COVID-19 patients.

## 2. RESULTS

### Topological data analysis

We decided to explore the data using the mapper algorithm for topological analysis of the data in order to find a distinction between severe and non-severe cases that the usual statistical tools fail to recover. The mapper algorithm is a method of replacing a topological space by a simpler one, which captures topological and geometric features of the original space. In the graph resulting from the mapper, the nodes represent the group of data that the algorithm considered similar while the edges between two of them mean that they have non-empty intersection. The size of the nodes is proportional to the number of elements it contains.

First we use the data from 262 patients with a single sample per patient, this means that we have a set of 262 points in ℝ^4^ where the entries correspond to the days after onset of symptoms, IgG antibody level, IgM antibody level and the severity with 0 indicating a non-severe case and 1 a severe case. In Figure 2 shows the resulting graph colored according to the severity of the patients described by the data set. The algorithm distinguishes three main groups, labeled A, B and C. In group A the nodes have at least 70% of severe patients, in group B the nodes have between 20% and 60% and in group C there are no severe patients.

**Figure 2:**
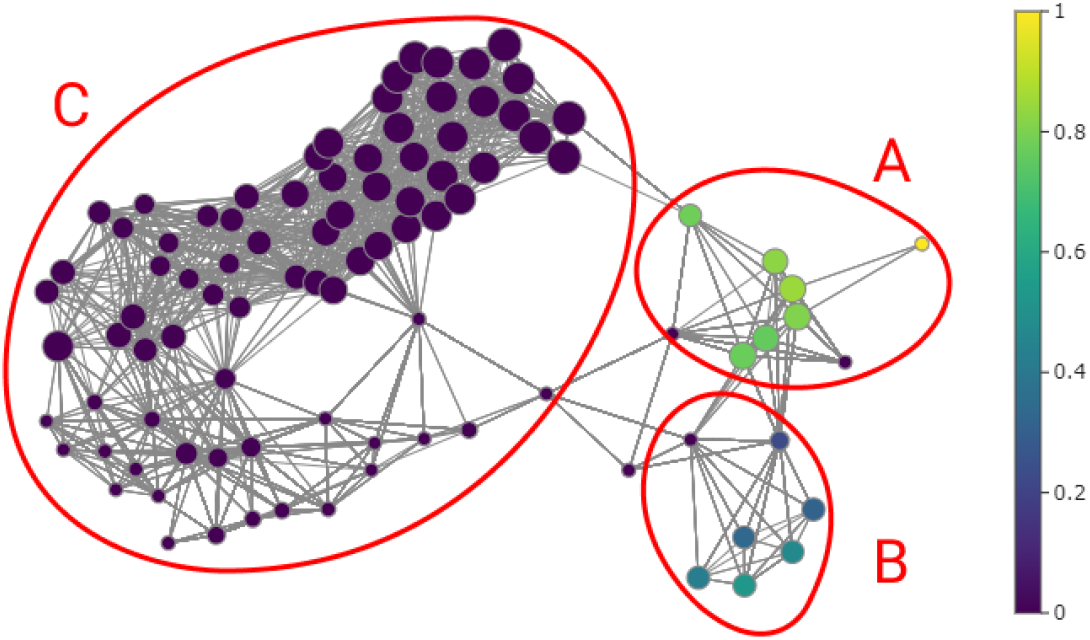
Graph generated with the mapper algorithm using 262 samples with the nodes colored by severity percentage. The algorithm distinguishes three main groups, in group A the nodes have at least 70% of samples of severe patients, in group B the nodes have between 20% and 60% and in group C there are no severe patients.

In order to study these distinctions, we show the violin plots in Figure 3. We first compared the IgG and IgM antibody levels of the data from node groups A, B, and C; and then comparing groups A ⋃ B and C separated by week. In the violin plots presented on the left of Figure 3 we observe that group B has clearly higher and less dispersed IgG antibody levels than the other groups. As for IgM in group A, it is generally higher than in the other groups. This may indicate a difference between severe cases, however it is insufficient with this information. The following two graphs compares IgG and IgM antibody levels between groups A ⋃ B and C per week. The A ⋃ B group represents mostly severe patients. The first three weeks the group A ⋃ B IgG antibody levels are higher than group C. As for IgM the diagrams are inconclusive. From the above we can conclude that high IgG antibody levels in the first weeks may be an indicator of a tendency towards a more severe disease state.

**Figure 3:**
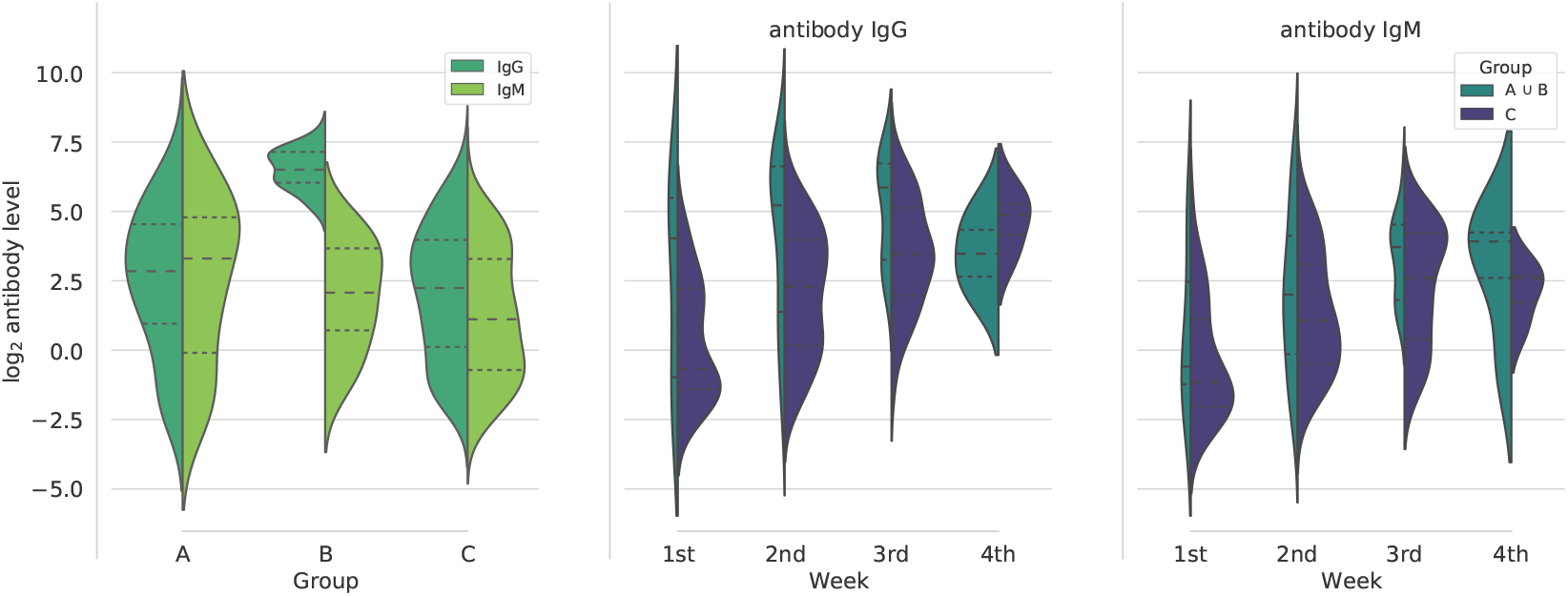
Violin plots comparing IgG and IgM antibody levels for each group A, B and C distinguished from the previous graph. The graph on the left compares IgG and IgM antibody levels between groups A, B and C. The following two graphs compares IgG and IgM antibody levels between groups A ⋃ B and C per week. Dashed lines are the median of the data and dotted lines indicate the interquartile range.

Of the 262 patients considered in the previous analysis, longitudinal samples were obtained from 41 of them. For the most part, samples were taken at three-day intervals. In this dataset the ID of each patient was also considered so that the mapper algorithm recognizes the samples coming from the same person, so the points are in ^5^. In Figure 4 shows the resulting graph colored according to the severity of the patients, the tail (group F) containing nodes with a large number of samples followed by a circular shape (group G) that ends with two distinct protrusions (group D and E). The group E contains the samples with high IgG antibody values, although there are some nodes with medium levels near the circle. In this same area, nodes with high levels of IgM antibodies are found. In group D, all nodes contain samples of severe patients, while in group E the percentage of severity varies between 0% and 90%. On the other hand, in groups F and G there are no samples of severe patients, however they differ with respect to the number of elements and the form in which they are presented. In group F all nodes have at least 12 elements (except for a single node) and those in group G have at most 11.

**Figure 4:**
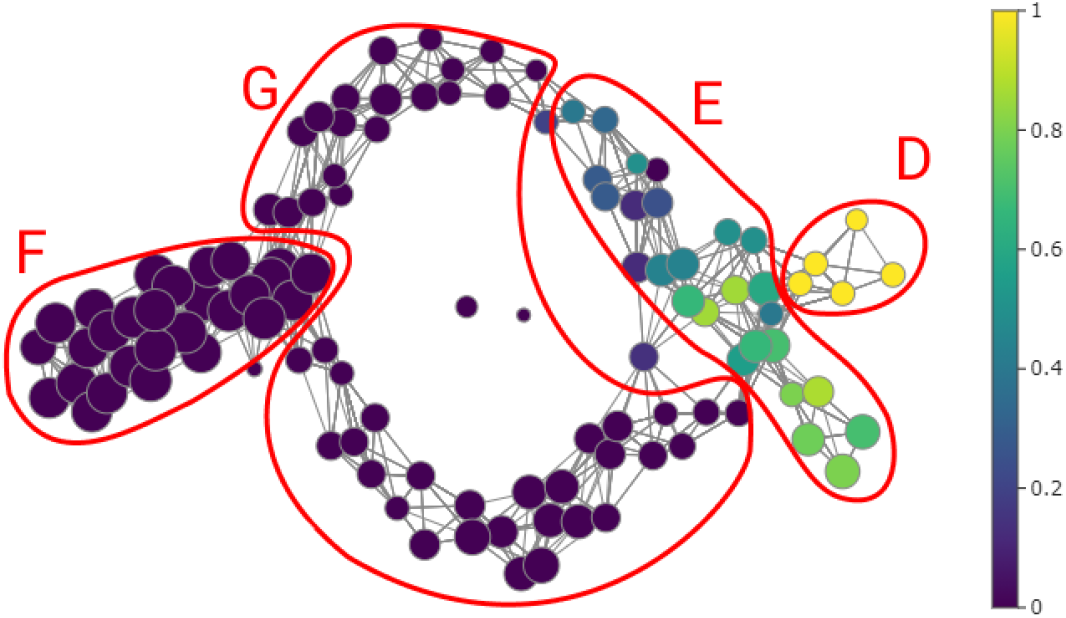
Graph generated with the Mapper algorithm using 41 longitudinal samples with the nodes colored by severity percentage. We can observe four main groups, in group D all nodes contain samples of severe patients, while in group E the percentage of severity varies between 0% and 90%. in groups F and G there are no samples of severe patients, however they differ with respect to the number of elements.

### 2.1. Mathematical models

The complexity and, at times, redundancy of immune responses to infections often result in arduous and expensive experimental settings when attempting to identify the key components and their temporal contributions. To dissect the dynamics observed in patients with COVID-19, mathematical modelling was employed not only as a quantitative recapitulation of experimental data but as a tool to support or reject less favourable hypotheses on the basis of various mathematical models as “thought experiments” using the Akaike Information Criterion (AIC) for the model selection process. We considered six different models. We will now describe three models that seem relevant to us; models 2, 4 and 5 are presented in the Supplementary Material.

**Model 1:** This model represents the viral dynamics of SARS-CoV-2 (*V*) and the dynamics of the T cell (*T*), B cell (*B*), IgM (*M*) and IgG (*G*) antibody response. The model is given by:

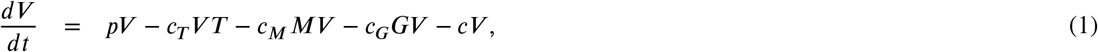

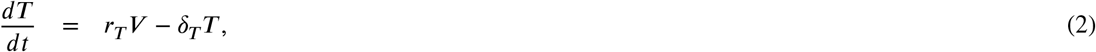

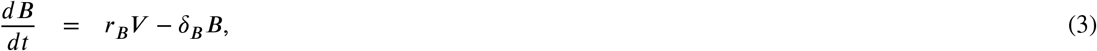

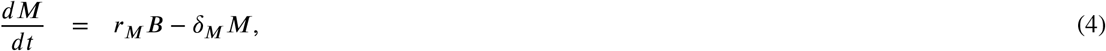

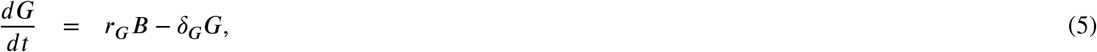

where *p* is the rate of viral replication, the terms *c*_*T*_ *V T, c*_*M*_ *MV* and *c*_*G*_*GV* are the clearance rate of the virus by the immune response T cells, IgM and IgG respectively. Clearance rate of the virus is *c*. Previous modelling work has suggested using half of the detection levels (less than 50 copies/ml) as initial viral concentration *V* (0); however, for patients with COVID-19, *V* (0) has been estimated approximately in 0.31 copies/ml using a regression model [16]. The proliferation rates of T cell and B cell are *r*_*T*_ and *r*_*B*_ respectively and are mediated by the viral load. The proliferation rate of IgM and IgG antibody are *r*_*M*_ and *r*_*G*_ respectively, and are mediated by the level of B cell. *δ*_*T*_, *δ*_*B*_, *δ*_*M*_ and *δ*_*G*_ are the half life of T cell, B cell, IgM antibody and IgM antibody, respectively. The initial levels of T cell, B cell, IgM antibody nd IgG antibody were set to 10^6^, 10, 0.1 and 0.1, respectively.

**Model 3:** In this model we considered the viral replication rate modelled by a logistic function with a replication rate *p* and a maximum carrying capacity *K*_*V*_ ; this is the maximum viral load. The model is given by:

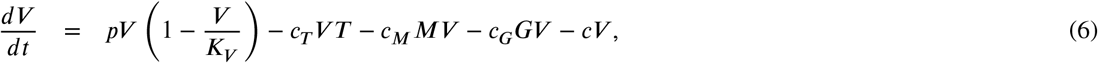

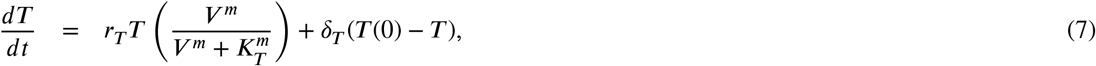

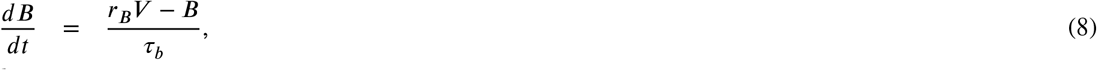

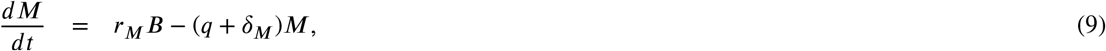

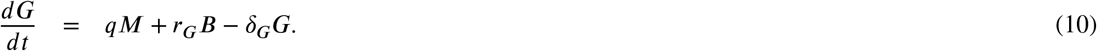

Equation (7) refers to the T cells response against SARS-CoV-2. T cells proliferate at a rate *r*_*T*_ and it is assumed that the activation of T cell proliferation by *V* follows a log-sigmoidal form with a half saturation constant *K*_*T*_. The coefficient *m* relates to the width of the sigmoidal function, different values of *m* were tested, *m* = 2 rendered a better fit [16]. *T* (0) is the initial T cells concentration, this represents the T cell homeostasis and it was set to 10^6^.

Equation (8) represent the B cells production, which activates the immunoglobulins response against the virus. This dynamic is modelled by the proliferation of the cells at a rate *r*_*B*_ minus the B cells already produce since these cells are directly involved in the immunoglobulins proliferation. The parameter τ_*b*_ correspond to the activation delay of the cells production.

Equation (9) refers to the IgM dynamic. The model infers that the production of antibodies is not simultaneous but independent, because IgM is considered a temporary antibody, in contrast to IgG, which shows prolonged immunity, and that the seroconversion process suggests the early production of IgM for its later conversion to IgG. This serocon-version process is represented by *q*. The dynamic of IgG antibody proliferation is represent in Eq. (10), where *q* is the growth rate associated with seroconversion.

**Model 6:** The dynamic of IgG antibody proliferation is modified. Here, the process in which seroconversion regulates the proliferation of IgG is modeled with a logistic function, where *r*_*q*_ is the growth rate associated with seroconversion and *K*_*q*_ is the maximum capacity of this process. Figure 5 shows the complexity of the process described by the following model:

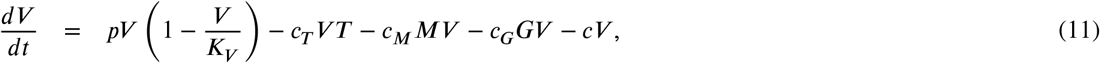

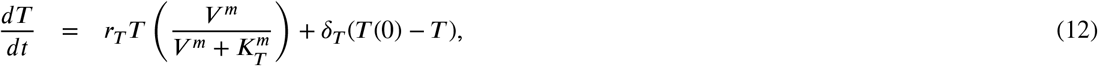

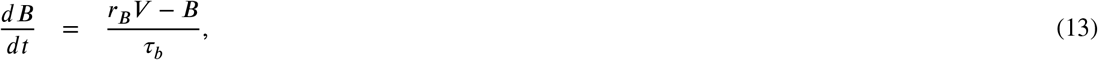

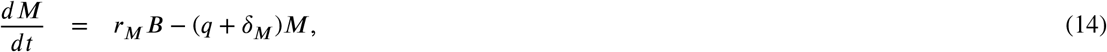

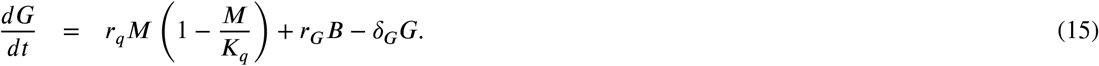

**Figure 5:**
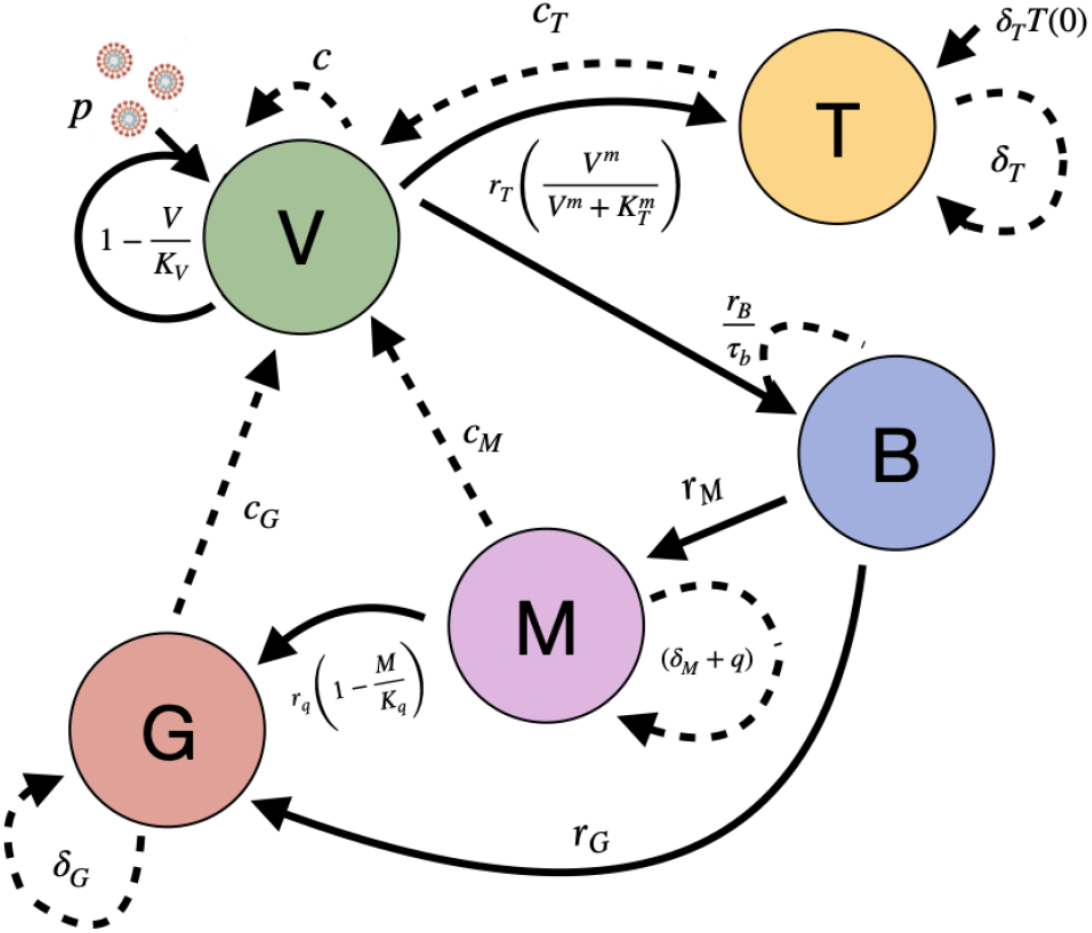
Schematic representation of the Model 6 proposed. Virus (*V*) are replicated at a rate of *p* and inhibited by T cells (*T*), B cells (*B*) and antibodies IgM (M) and IgG (G). *V* induces *B* and *T* proliferation. On the other hand, *B* induces *M* and *G* proliferation, while that seroconversion process regulates *G* proliferation due to *M*. Positive regulations between processes are highlighted with solid arrows, while negative regulations are marked with dotted arrows.

### 2.2. Stability analysis

In this section we perform a sensitivity analysis of Model 6, since it was the one with the best AIC score. To begin with, Eqs. (11)-(15) can be transformed into the following dimensionless form:

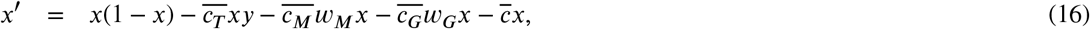

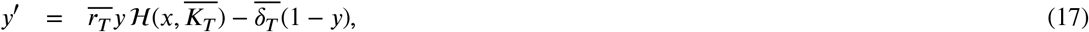

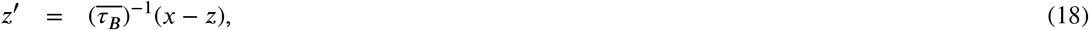

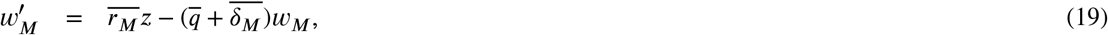

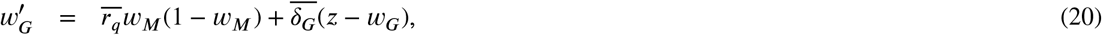

where the function ℋ is defined by ℋ (*x, a*) = *x*^*m*^/(*x*^*m*^ + *a*^*m*^) with *m* ≥ 2, and the derivatives are with respect to dimensionless time *τ* = *pt*. And the overline symbols are the dimensionless parameter that can be obtained by substituting in Eqs. (11)-(15) the variables defined by:

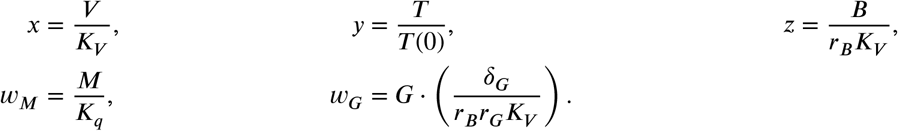

For convenience, we call an equilibrium point with viral load *x virus-free* when *x* = 0 and *virus-positive* when *x* > 0. We will only consider equilibrium points with non-negative coordinates. Basic computation yields exactly one virus-free equilibrium which is *E*_0_ := (0, 1, 0, 0, 0).

Let *J* (*p*) denote the following Jacobian matrix:

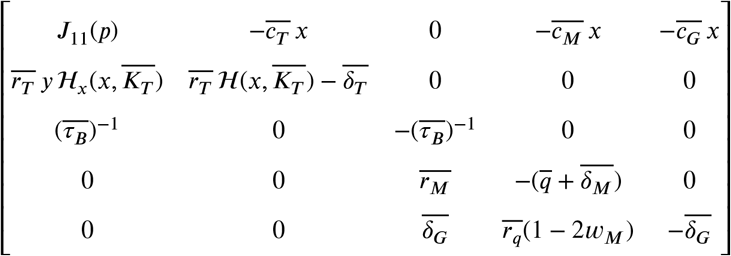

where ℋ*x* = *δ*ℋ/*δx* and

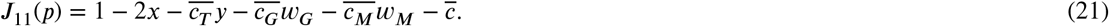

#### Theorem 1.

*Let*

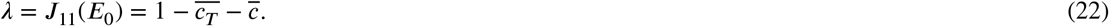

*Then E*_0_ *is asymptotically stable for λ <* 0 *and non-hyperbolic for λ* = 0. *If λ >* 0, *then E*_0_ *is an unstable saddle with a four-dimensional stable manifold and a one-dimensional unstable manifold*.

*Proof*. Evaluating the Jacobian matrix at *E*_0_ yields

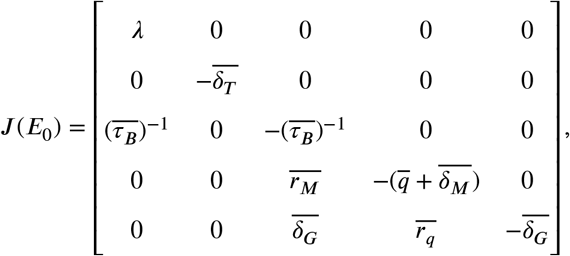

which implies that *E*_0_ is asymptotically stable for *λ <* 0 and is non-hyperbolic for *λ* = 0. If *λ* > 0, then *J* (*E*_0_) has exactly one positive eigenvalue *λ* and four negative eigenvalues. Hence by the Stable Manifold Theorem, *E*_0_ is an unstable saddle with a four-dimensional stable manifold and a one-dimensional unstable manifold.

*Remark*. In the case where *E*_0_ becomes a saddle for *λ >* 0, the solutions *ϕ*_1_(*x*) such that *ϕ*_1_(*x*) → *E*_0_ at *t* → ∞ form a four-dimensional invariant manifold, called the stable manifold of *E*_0_.

Our dimensionless model, Eqs. (16) to (20), admits virus-positive equilibrium points only if *λ >* 0. This point with viruses is more complicated to obtain. In the Supplementary Material we present an analysis of the different scenarios in which this equilibrium point can occur.

### 2.3. Numerical simulations

In order to study the antibody response during the course of SARS-CoV-2 infection, we fixed the parameters related to the dynamics of the virus and T cell with values obtained from [16] and [23]. We estimated the parameters related to the dynamics of the B cells and antibodies IgM and IgG. To estimate these parameters we selected 39 patients who presented a significant temporal change of IgG antibody data and fit our models to that data; of these 39 patients, 3 were severe cases found in group D, 4 were severe cases from group E, and 32 were non-severe cases belonging to groups F and G, identified by the topological analysis presented above (Figure 4). The fixed parameters are displayed in Table 1, and they are taken the same for all cases.

**Table 1.** Model Parameters.

| Parameter | Value |
| --- | --- |
| $p$ | 6.11 [16] |
| $K_V$ | $1 \times 10^8$ [16] |
| $c_T$ | $7.8 \times 10^{-7}$ [16] |
| $c_M$ | Fitted |
| $c_G$ | Fitted |
| $c$ | 2.4 [16] |
| $\delta_T$ | 0.1 [16] |
| $r_T$ | 0.281 [16] |
| $K_T$ | $3.16 \times 10^4$ [16] |
| $r_B$ | Fitted |
| $\tau_b$ | Fitted |
| $r_M$ | Fitted |
| $q$ | Fitted |
| $\delta_M$ | 0.0693 [23] |
| $r_q$ | Fitted |
| $K_q$ | Fitted |
| $r_G$ | Fitted |
| $\delta_G$ | 0.0277 [23] |

Parameter optimization was carried out with data from each of the 39 patients considered. The six models were fitted to the data of the 39 patients, and we calculated the AIC score for each patient from Eq. (24). The results showed that Model 6 had a better score for non-severe patients, but was not the best for severe patients. However, the difference between the score of Model 6 in non-severe with the rest of the models is greater than comparing Model 4 to 6 in the severe patient group, therefore we chose Model 6 as the one that best explains the viral dynamic and immune response. The AIC scores can be consulted in the Supplementary Material.

Figure 6 display the dynamics of the viral load and the immune response for one severe patient from group D (severe D), one severe patient from group E (severe E), and one non-severe patient from group F and G, using Model 6. In Figure 6e we can observe the fit of the IgG response of Model 6 with the experimental data points. In these particular cases, the peak of the viral load for the severe E patient in Figure 6a was lower than other two, while severe D patient took more days to clear the virus. The peak of T cell level for the severe D patient in Figure 6b was higher than other two cases, and severe E patient had the lowest T cell response in accordance with her/his lower viral load. The B cell level for severe D patient in Figure 6c maintained a higher level until the end of the disease, while the severe E had the lowest level again. The non-severe patient had the highest IgM antibody level in Figure 6d and severe D patient had the lowest IgG antibody level in Figure 6e.

**Figure 6:**
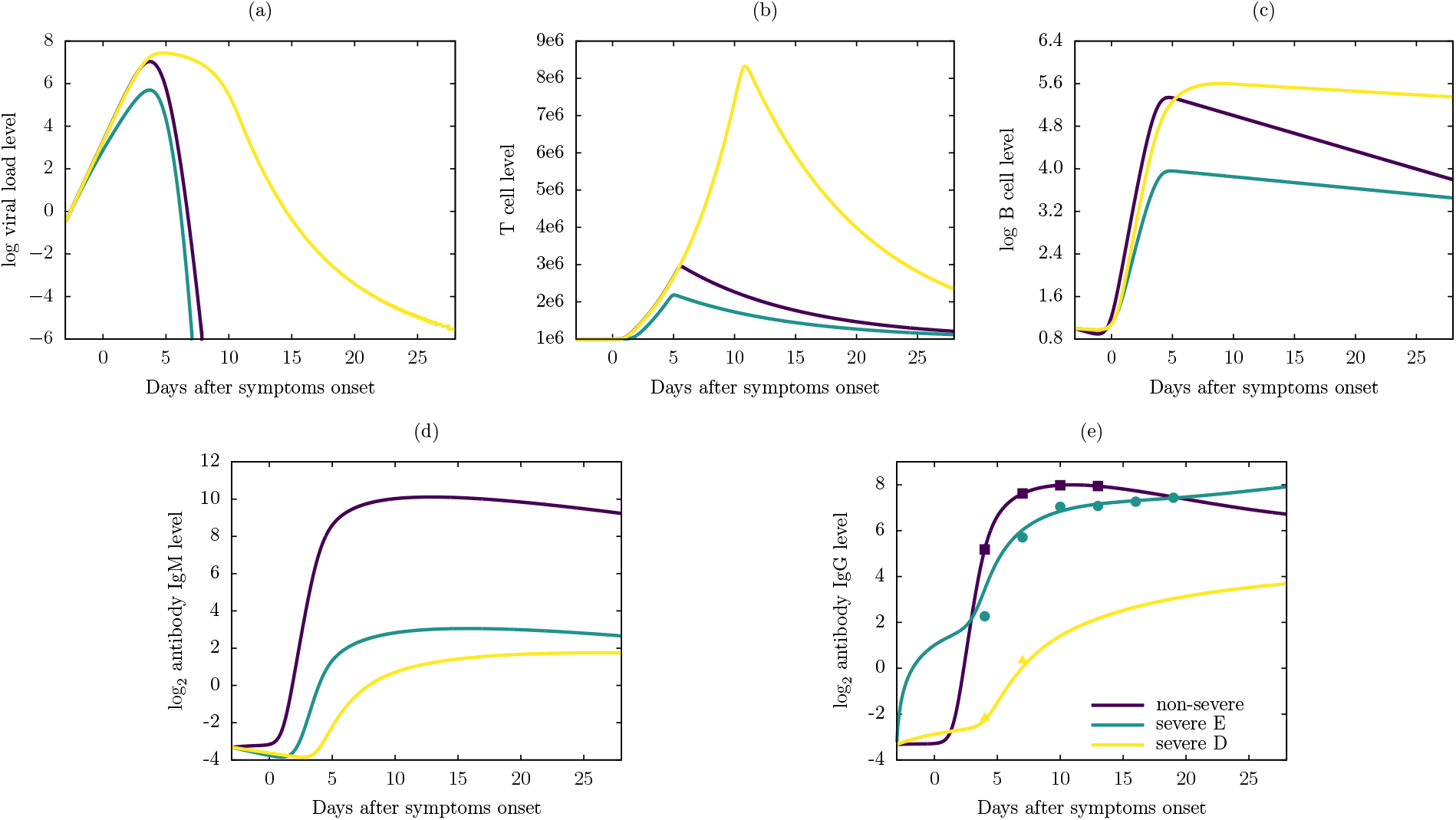
Numerical results of the Model 6 using antibody IgG level data from one severe patient from group D, one severe patient from group E, and non-severe patient. (a) Viral load level, (b) T cell level, (c) B cell level, (d) antibody IgM level and (e) antibody IgG level. The points in (e) are experimental data of antibody IgG level reported in [21], triangle for severe D case (Patient ID: 5), circle for a severe E case (Patient ID: 3) and square for a non-severe case (Patient ID: 1). Notice the log base 10 scale in (a) and (c), and log base 2 scale in (d) and (e).

A total of 9 parameters were optimized in Model 6 and each patient had a set of parameters that best fit their antibody IgG response data. The distribution of each parameter for all patients can be consulted in the Supplementary Material. The parameters *r*_*B*_, *r*_*M*_ and *r*_*G*_ presented little variation between patients, even between severe and non-severe patients; these parameters represent the proliferation of B cells, antibodies IgM and antibodies IgG, respectively. This result suggests that the B cell response and the antibody IgM level do not play an important role in identifying a severe patient. Although *r*_*G*_ does not vary sufficiently between two groups of patients, *q* show a difference where the non-severe group has lower values, while severe group has uniformly distributed values. The rest of the parameters do not show clear differences between groups, tending to a uniform distribution in both groups. It should be noticed that the group of severe patients is not large enough to give conclusive results.

In Table 2 is displayed the median of the estimated parameters of the patients in the non-severe group and the two severe groups. The parameters that varied the most in their medians were *c*_*G*_, *r*_*M*_, and *r*_*G*_, with orders of magnitude among patient groups. It can be noticed that *r*_*q*_ and *r*_*G*_ values are lower in non-severe patients than severe D patients, however these patients have a viral load clearance similar to severe D patients; this suggests that severe D patients do not produce “quality” antibodies that help to viral shedding, which consequently leads to more severe symptoms. We can also note that although severe D patients have generally better parameters than severe E patients they have a early B-cell proliferation (low values of *τ*_*B*_); this result support the idea of quality antibody production, having early antibody production lowers the efficiency in clearing viral load.

**Table 2.**
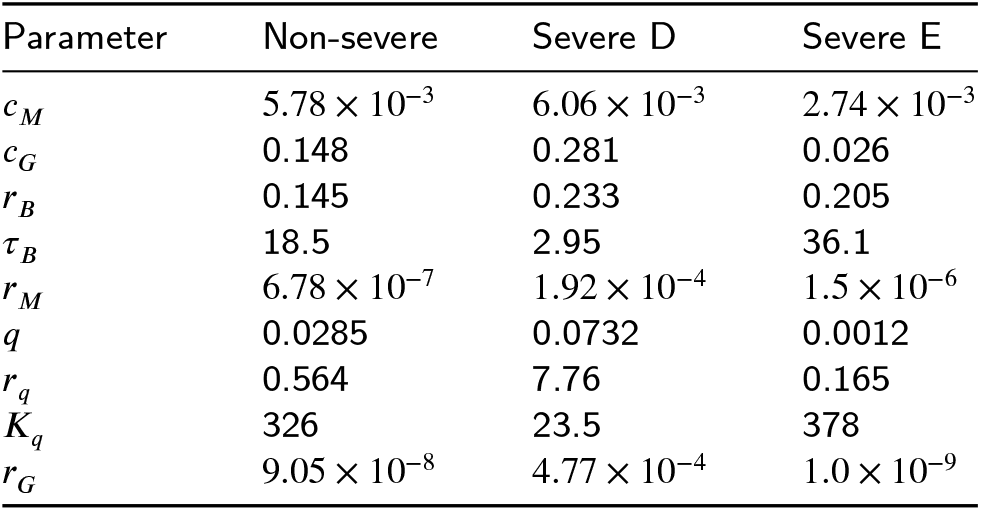
Median of the optimized parameters of severe D patients, severe E patients and non-severe patients.

| Parameter | Non-severe | Severe D | Severe E |
| --- | --- | --- | --- |
| $c_M$ | $5.78 \times 10^{-3}$ | $6.06 \times 10^{-3}$ | $2.74 \times 10^{-3}$ |
| $c_G$ | 0.148 | 0.281 | 0.026 |
| $r_B$ | 0.145 | 0.233 | 0.205 |
| $\tau_B$ | 18.5 | 2.95 | 36.1 |
| $r_M$ | $6.78 \times 10^{-7}$ | $1.92 \times 10^{-4}$ | $1.5 \times 10^{-6}$ |
| $q$ | 0.0285 | 0.0732 | 0.0012 |
| $r_q$ | 0.564 | 7.76 | 0.165 |
| $K_q$ | 326 | 23.5 | 378 |
| $r_G$ | $9.05 \times 10^{-8}$ | $4.77 \times 10^{-4}$ | $1.0 \times 10^{-9}$ |

## 3. Discussion

In this work, we used the mapper algorithm for the exploration of antibody level data of patients with COVID-19. Using this tool, we identified three groups of patients: non-severe, severe and a group with intermediate severity. We found that the last two groups have a notable difference in the level of IgG antibodies, being higher in the intermediate group.

On the other hand, we set out to model the immune system response against SARS-CoV-2 viral infection by focusing on the antibody response. It is not yet clear how the immune system of a patient presenting with severe symptoms fails to clear the viral load from the body, however, it has been reported that severe COVID-19 patients have high IgG antibody levels [45, 21]. Therefore, we proposed six mathematical models that represent the response of the immune system against viral infection; we estimated the parameters related to B cell response and IgM and IgG antibodies using IgG antibody data reported COVID-19 patients. Among all models, Model 6 provided the best fit to the IgG data. Our results show that the key parameters between the dynamics of severe and non-severe patients are those related to antibody proliferation and viral clearance by IgG antibody.

In Table 2 we can observe that the severe D patients have a early B cell proliferation, this confers an early response by B cells, which in turn trigger the production of antibodies. This early B cell production in severe patients leads to a rapid proliferation of antibodies, as we can see from the higher values of *r*_*M*_ and *r*_*G*_ in the severe D group. Due to this short response time, the antibodies produced by these severe patients could not have enough time to carry out the selection process of the optimal receptor for binding with the S protein of the virus, and therefore they would be considered of low quality or even not having neutralizing capacity. This could be the reason why they do not have a great contribution in viral clearance, since the parameter *c*_*G*_ corresponding to viral clearance by IgG antibody is lower in severe patients. This suggest that antibodies produced by severe patients do not present the expected neutralizing response, considered of low quality as reported by Vanshylla et al. [48]. Different works have identified antibodies that potently can neutralize SARS-CoV-2 derived from individuals infected with COVID-19 [18, 19, 36]. In fact, there is evidence that robust neutralizing antibodies to SARS-CoV-2 infection could persist for several months [25]. Furthermore, cross-reactivity in antibody binding to the spike protein is common, but cross-neutralization of the live viruses is rare[22]. Nevertheless, antibody affinity may have several implications. In fact, limited antibody protection against any virus have a theoretical potential to amplify the infection [3].

Another key process in differentiating severe and non-severe patients is the ancillary process in IgG production by the seroconversion period. This process proved to be key to emulate the SARS-CoV-2 infection process and the adaptive immune response, since considering a sigmoidal activation function of the seroconversion process from IgM to IgG improved the score of the Model 6. The rate of antibody IgM converted to IgG due to the seroconversion process, represented by *q*, was higher in severe D patients, which implies that there is a delay in the seroconversion process in non-severe patients. The latter suggests that in non-severe patients there persists a level of IgM antibodies that help in viral clearance before activating the seroconversion process; in severe patients there is an early production of IgG antibodies that are not efficient in neutralizing viral particles.

Since model 6 was the best fit to the data, we performed a stability analysis. This analysis can help us understand how the immune system, and especially the antibodies, clear the viral load. If we take *m* = 2 then we can expect up to 5 equilibrium points in the 5-dimensional Model 6, and the existence criteria of each type of equilibrium is studied. A point of stability is when there are no viral particles, and it is asymptotically stable. There is at least a virus-positive equilibrium point, *λ >* 0, this means that the viral proliferation rate *p* is not overwhelmed by the elimination rates *c* and *c*_*T*_.

## 4. Methodologies

### 4.1. Experimental data details

In this work we used experimental data reported by Long *et al*. [21]. In that study the authors reported the antibody response of 285 COVID-19 patients of which 70 had sequential samples available. The patients were enrolled from three hospitals in Chongqing, a municipality adjacent to Hubei province. The median age of the patients was 47 years and 54.5% were males. Serological samples were collected from symptoms onset and the detection of IgM and IgG levels in response to SARS-Cov-2 was carried out using MCLIA (Magnetic Chemiluminescence Immunoassay) kits. The antibody levels were presented as the measured chemiluminescence values divided by the cutoff value which was defined by receiver operating characteristic curves.

Of the 285 patients, 39 were classified as severe or in critical condition and 63 patients were followed up for serological sampling, where samples were taken at 3-day intervals between February 8, 2020 and the patient’s discharge from the hospital. Of the latter group, 100% tested positive for the presence of IgG antibody approximately 17-19 days after the symptoms onset (daso), while 94% of the patients reached peak IgM levels approximately 20-22 daso. IgG and IgM levels in the group of patients classified as severe were higher than the non-severe group, however a significant difference was only observed in IgG levels. More details can be found in the original paper [21].

### 4.2. Mapper algorithm

For the topological analysis of the data, the *giotto-tda* package was used [46]. This package integrates various TDA tools with machine learning using an API compatible with scikit-learn and C++ implementations. This package has the great advantage of providing a balance between interoperability and computational efficiency, which is useful given the sensitivity of the mapper to parameter changes. The latter is still an important problem, as there is still no clear methodology for parameter selection. In [6, 44] the authors explore ways to attack this problem using category theory.

As mentioned above, the choice of parameters of the mapper algorithm directly influences the results obtained. Among the parameters that can be modified are the metric space, lens, clustering algorithm, number of intervals and percentage overlap. In a previos work, Sasaki et al. [40] studied with TDA the shape of the immune response during co-infections, in this work the mapper algorithm and each of the listed parameters are described in detail; it is recommended to refer to that work to obtain a broader overview of this analysis tool.

In Table 3 is displayed the parameters used for the analysis of the two data sets, considering a sample per patient for 262 patients, and longitudinal samples of 41 patients. It should be pointed out that we tested different choices of parameters, but we only present the ones with which we obtained the best results for this data set.

**Table 3.** Parameters used in mapper algorithm for each data set.

| Parameter | 262-patients dataset | 41-patients dataset |
| --- | --- | --- |
| Metric space | cosine | cosine |
| Lens | Eccentricity (exp = 2) | Eccentricity (exp = 2) |
| Clustering algorithm | DBSCAN (eps = 0.01) | DBSCAN (eps = 0.1) |
| Number of intervals | 100 | 100 |
| Percentage overlap | 95% | 88% |

### 4.3. Parameter estimation

Mathematical models proposed in this work is based on Ordinary Differential Equations (ODEs). The ODEs were solved using the PDEparams module [31], which integrates the differential equations using SciPy’s odeint. We used different mechanistic modeling strategies, a total of 6 models were tested. The estimation of the free parameters was performed using the same PDEparams module where the parameter estimation is carried out using Differential Evolution (DE) algorithm [43], which minimize a cost function. In this work we used experimental data that are given in logarithmic scale, therefore we use the Root Mean Square Log Error (RMSLE) as cost function and is given by:

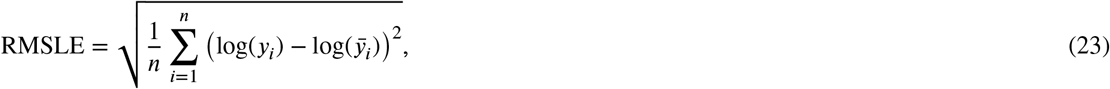

where *n* is the number of data points (samples), *y*_*i*_ is the experimental measure of the *i*-th sample and 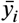 is the predictive output of the model.

As several models can provide the same fit with observed experimental data, it becomes necessary to choose between different models. The standard approach to model selection is first estimate all model parameters from the data, then select the model with the best-fit error and some penalties on model complexity. In this work we used the next model selection criteria.

**Definition 1. Akaike Information Criterion (AIC)**. The corrected (AIC) writes as follows:

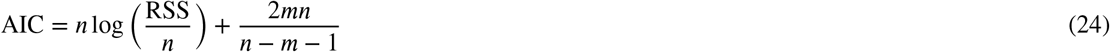

where *n* is the number of data points, *m* is the unknown parameters and RSS is the residual sum of squares obtained from the fitting routine. The lowest AIC value of a given model, the best description of the data better respect the other models. Small differences in AIC scores (*e*.*g. <*2) are not significant [7].

## Data Availability

All data produced in the present study are available upon reasonable request to the authors

## Acknowledgment

This research was funded by the Universidad Nacional Autonoma de México (UNAM) and the Alfons und Gertrud Kassel-Stiftung.

## References

[1] Abuin, P., Anderson, A., Ferramosca, A., Hernandez-vargas, E.A., Gonzalez, A.H., 2020. Annual Reviews in Control Characterization of SARS-CoV-2 dynamics in the host. Annual Reviews in Control URL: https://doi.org/10.1016/j.arcontrol.2020.09.008, doi:10.1016/j.arcontrol.2020.09.008.

[2] Almocera, A.E.S., Quiroz, G., Hernandez-Vargas, E.A., 2020. Stability analysis in COVID-19 within-host model with immune response. Communications in Nonlinear Science and Numerical Simulation doi:10.1016/j.cnsns.2020.105584.

[3] Arvin, A.M., Fink, K., Schmid, M.A., Cathcart, A., Spreafico, R., Havenar-Daughton, C., Lanzavecchia, A., Corti, D., Virgin, H.W., 2020. A perspective on potential antibody-dependent enhancement of SARS-CoV-2. Nature 584, 353–363. URL: http://dx.doi.org/10.1038/s41586-020-2538-8, doi:10.1038/s41586-020-2538-8.

[4] Baccam, P., Beauchemin, C., Macken, C.a., Hayden, F.G., Perelson, A.S., 2006. Kinetics of influenza A virus infection in humans. Journal of virology 80, 7590–9. URL: http://www.ncbi.nlm.nih.gov/pubmed/16840338, doi:10.1128/JVI.01623-05.

[5] Blanco-Rodríguez, R., Du, X., Hernández-Vargas, E., 2021. Computational simulations to dissect the cell immune response dynamics for severe and critical cases of sars-cov-2 infection. Computer methods and programs in biomedicine 211, 106412.

[6] Bui, Q.T., Vo, B., Do, H.A.N., Hung, N.Q.V., Snasel, V., 2020. F-mapper: A fuzzy mapper clustering algorithm. Knowledge-Based Systems 189, 105107.

[7] Burnham, K.P., Anderson, D.R., 2002. Model selection and multimodel inference: a practical information-theoretic approach. Springer Science & Business Media.

[8] Dobrovolny, H.M., Reddy, M.B., Kamal, M.a., Rayner, C.R., Beauchemin, C.a.a., 2013. Assessing Mathematical Models of Influenza Infections Using Features of the Immune Response. PLoS ONE 8, e57088.

[9] Du, S.Q., Yuan, W., 2020. Mathematical Modeling of Interaction between Innate and Adaptive Immune Responses in COVID-19 and Implications for Viral Pathogenesis. Journal of Medical Virology, 0–2doi:10.1002/jmv.25866.

[10] Ejima, K., Kim, K.S., Ito, Y., Iwanami, S., Ohashi, H., Koizumi, Y., Watashi, K., Bento, A.I., Aihara, K., Iwami, S., 2020. Inferring Timing of Infection Using Within-host SARS-CoV-2 Infection Dynamics Model: Are “Imported Cases” Truly Imported? medRxiv 4297, 2020.03.30.20040519. doi:10.1101/2020.03.30.20040519.

[11] Gonçalves, A., Bertrand, J., Ke, R., Comets, E., de Lamballerie, X., Malvy, D., Pizzorno, A., Terrier, O., Calatrava, M.R., Mentré, F., Smith, P., Perelson, A.S., Guedj, J., 2020. Timing of antiviral treatment initiation is critical to reduce SARS-Cov-2 viral load. medRxiv URL: http://www.ncbi.nlm.nih.gov/pubmed/32511641http://www.pubmedcentral.nih.gov/articlerender.fcgi?artid= PMC7276997, doi:10.1101/2020.04.04.20047886.

[12] Goyal, A., Cardozo-Ojeda, E., Schiffer, J.T., 2020. Potency and timing of antiviral therapy as determinants of duration of SARS CoV-2 shedding and intensity of inflammatory response. medRxiv, 2020.04.10.20061325URL: http://medrxiv.org/content/early/2020/04/14/2020.04.10.20061325.abstract, doi:10.1101/2020.04.10.20061325.

[13] Graw, F., Perelson, A.S., 2015. Modeling Viral Spread. Annual Review of Virology, 1–18URL: http://www.annualreviews.org/doi/abs/10.1146/annurev-virology-110615-042249, doi:10.1146/annurev-virology-110615-042249.

[14] Hernandez-Vargas, A.E., Meyer-Hermann, M., 2012. Innate immune system dynamics to influenza virus. IFAC Proceedings Volumes 45, 260–265.

[15] Hernandez-Vargas, E., 2017. Modeling kick-kill strategies toward HIV cure. Frontiers in Immunology 8. doi:10.3389/fimmu.2017.00995.

[16] Hernandez-Vargas, E.A., Velasco-Hernandez, J.X., 2020. In-host Mathematical Modelling of COVID-19 in Humans. Annual Reviews in Control doi:10.1016/j.arcontrol.2020.09.006.

[17] Hernandez-Vargas, E.A., Wilk, E., Canini, L., Toapanta, F.R., Binder, S.C., Uvarovskii, A., Ross, T.M., Guzman, C., Perelson, A.S., Meyer-Hermann, M., 2014. Effects of aging on influenza virus infection dynamics. Journal of Virology 88, 4123–31.

[18] Ju, B., Zhang, Q., Ge, J., Wang, R., Sun, J., Ge, X., Yu, J., Shan, S., Zhou, B., Song, S., Tang, X., Yu, J., Lan, J., Yuan, J., Wang, H., Zhao, J., Zhang, S., Wang, Y., Shi, X., Liu, L., Zhao, J., Wang, X., Zhang, Z., Zhang, L., 2020. Human neutralizing antibodies elicited by SARS-CoV-2 infection. Nature 584, 115–119. URL: http://dx.doi.org/10.1038/s41586-020-2380-z, doi:10.1038/s41586-020-2380-z.

[19] Liu, L., Wang, P., Nair, M.S., Yu, J., Rapp, M., Wang, Q., Luo, Y., Chan, J.F., Sahi, V., Figueroa, A., Guo, X.V., Cerutti, G., Bimela, J., Gorman, J., Zhou, T., Chen, Z., Yuen, K.Y., Kwong, P.D., Sodroski, J.G., Yin, M.T., Sheng, Z., Huang, Y., Shapiro, L., Ho, D.D., 2020. Potent neutralizing antibodies against multiple epitopes on SARS-CoV-2 spike. Nature 584, 450–456. URL: http://dx.doi.org/10.1038/s41586-020-2571-7, doi:10.1038/s41586-020-2571-7.

[20] Liu, Y., Yan, L.M., Wan, L., Xiang, T.X., Le, A., Liu, J.M., Peiris, M., Poon, L.L.M., Zhang, W.,. Viral dynamics in mild and severe cases of COVID-19. The Lancet Infectious Diseases 0. URL: https://www.thelancet.com/journals/laninf/article/PIIS1473-3099(20)30232-2/fulltext, doi:10.1016/S1473-3099(20)30232-2.

[21] Long, Q.X., Liu, B.Z., Deng, H.J., Wu, G.C., Deng, K., Chen, Y.K., Liao, P., Qiu, J.F., Lin, Y., Cai, X.F., et al., 2020. Antibody responses to sars-cov-2 in patients with covid-19. Nature Medicine 26, 845–848.

[22] Lv, H., Wu, N.C., Tsang, O.T.Y., Yuan, M., Perera, R.A.P.M., Leung, W.S., So, R.T.Y., Chan, J.M.C., Yip, G.K., Chik, T.S.H., Wang, Y., Choi, C.Y.C., Lin, Y., Ng, W.W., Zhao, J., Poon, L.L.M., Peiris, J.S.M., Wilson, I.A., Mok, C.K.P., 2020. Cross-reactive antibody response between SARS-CoV-2 and SARS-CoV infections. bioRxiv, 2020.03.15.993097URL: https://www.biorxiv.org/content/10.1101/2020.03.15.993097v1, doi:10.1101/2020.03.15.993097.

[23] Mankarious, S., Lee, M., Fischer, S., Pyun, K., Ochs, H., Oxelius, V., Wedgwood, R., 1988. The half-lives of igg subclasses and specific antibodies in patients with primary immunodeficiency who are receiving intravenously administered immunoglobulin. The Journal of Laboratory and Clinical Medicine 112, 634–640.

[24] Miao, H., Hollenbaugh, J.A., Zand, M.S., Holden-Wiltse, J., Mosmann, T.R., Perelson, A.S., Wu, H., Topham, D.J., 2010. Quantifying the Early Immune Response and Adaptive Immune Response Kinetics in Mice Infected with Influenza A Virus. Journal of Virology 84, 6687–6698. URL: http://jvi.asm.org/cgi/doi/10.1128/JVI.00266-10, doi:10.1128/JVI.00266-10.

[25] Mudd, P.A., Adv, S., Mudd, P.A., Crawford, J.C., Turner, J.S., Souquette, A., Reynolds, D., Bosanquet, J.P., Anand, N.J., Striker, D.A., Martin, R.S., Boon, A.C.M., House, S.L., Remy, K.E., Hotchkiss, R.S., Presti, R.M., Halloran, J.A.O., Powderly, W.G., Thomas, P.G., Ellebedy, A.H., 2020. Distinct inflammatory profiles distinguish COVID-19 from influenza with limited contributions from cytokine storm. Science 3024, 1–21. doi:10.1126/sciadv.abe3024.

[26] Nguyen, V.K., Binder, S.C., Boianelli, A., Meyer-Hermann, M., Hernandez-Vargas, E.A., 2015. Ebola virus infection modeling and identifiability problems. Frontiers in Microbiology 6, 1–11.

[27] Nguyen, V.K., Hernandez-Vargas, E.A., 2017. Windows of opportunity for Ebola virus infection treatment and vaccination. Scientific reports 7, 8975.

[28] Nicolau, M., Levine, A.J., Carlsson, G., 2011. Topology based data analysis identifies a subgroup of breast cancers with a unique mutational profile and excellent survival. Proceedings of the National Academy of Sciences 108, 7265–7270.

[29] Oh, M.d., Park, W.B., Choe, P.G., Choi, S.J., Kim, J.I., Chae, J., Park, S.S., Kim, E.C., Oh, H.S., Kim, E.J., Nam, E.Y., Na, S.H., Kim, D.K., Lee, S.M., Song, K.H., Bang, J.H., Kim, E.S., Kim, H.B., Park, S.W., Kim, N.J., 2016. Viral Load Kinetics of MERS Coronavirus Infection. New England Journal of Medicine 375, 1303–1305. URL: http://www.nejm.org/doi/10.1056/NEJMc1511695, doi:10.1056/NEJMc1511695.

[30] Oja, A.E., Saris, A., Ghandour, C.A., Kragten, N.A., Hogema, B.M., Nossent, E.J., Heunks, L.M., Cuvalay, S., Slot, E., Linty, F., Swaneveld, F.H., Vrielink, H., Vidarsson, G., Rispens, T., Schoot, E., Lier, R.A., Brinke, A.T., Hombrink, P., 2020. Divergent SARS-CoV-2-specific T and B cell responses in severe but not mild COVID-19 patients. European Journal of Immunology doi:10.1002/eji.202048908.

[31] Parra-Rojas, C., Hernandez-Vargas, E.A., 2020. Pdeparams: Parameter fitting toolbox for partial differential equations in python. Bioinformatics 36, 2618–2619.

[32] Pawelek, K.A., Huynh, G.T., Quinlivan, M., Cullinane, A., Rong, L., Perelson, A.S., 2012. Modeling Within-Host Dynamics of Influenza Virus Infection Including Immune Responses. PLoS Comput Biol 8, e1002588.

[33] Peiris, J.S.M., Chu, C.M., Cheng, V.C.C., Chan, K.S., Hung, I.F.N., Poon, L.L.M., Law, K.I., Tang, B.S.F., Hon, T.Y.W., Chan, C.S., Chan, K.H., Ng, J.S.C., Zheng, B.J., Ng, W.L., Lai, R.W.M., Guan, Y., Yuen, K.Y., HKU/UCH SARS Study Group, 2003. Clinical progression and viral load in a community outbreak of coronavirus-associated SARS pneumonia: a prospective study. Lancet (London, England) 361, 1767–72. URL: http://www.ncbi.nlm.nih.gov/pubmed/12781535, doi:10.1016/s0140-6736(03)13412-5.

[34] Perelson, A.S., Ribeiro, R.M., 2013. Modeling the within-host dynamics of HIV infection. BMC Biology 11, 96. URL: http://bmcbiol.biomedcentral.com/articles/10.1186/1741-7007-11-96, doi:10.1186/1741-7007-11-96, 9903017.

[35] Pinkevych, M., Kent, S.J., Tolstrup, M., Lewin, S.R., Cooper, D.A., Søgaard, O.S., Rasmussen, T.A., Kelleher, A.D., Cromer, D., Davenport, M.P., 2016. Modeling of Experimental Data Supports HIV Reactivation from Latency after Treatment Interruption on Average Once Every 5–8 Days. PLoS Pathogens 12, 8–11. URL: http://dx.doi.org/10.1371/journal.ppat.1005740, doi:10.1371/journal.ppat.1005740.

[36] Ravichandran, S., Coyle, E.M., Klenow, L., Tang, J., Grubbs, G., Liu, S., Wang, T., Golding, H., Khurana, S., 2020. Antibody signature induced by SARS-CoV-2 spike protein immunogens in rabbits. Science Translational Medicine 12, 1–9. doi:10.1126/SCITRANSLMED.ABC3539.

[37] Reluga, T.C., Dahari, H., Perelson, A.S., 2009. Analysis if Hepatitis C Virus Infection Models with Hepatocyte Homeostasis. SIAM journal on applied mathematics 69, 999–1023.

[38] Robles, A., Hajij, M., Rosen, P., 2017. The shape of an image: A study of mapper on images. arXiv preprint 1710.09008.

[39] Rong, L., Ã, A.S.P., 2009. Modeling HIV persistence, the latent reservoir, and viral blips. Journal of Theoretical Biology 260, 308–331. URL: http://dx.doi.org/10.1016/j.jtbi.2009.06.011, doi:10.1016/j.jtbi.2009.06.011.

[40] Sasaki, K., Bruder, D., Hernandez-Vargas, E.A., 2020. Topological data analysis to model the shape of immune responses during co-infections. Communications in Nonlinear Science and Numerical Simulation 85, 105228.

[41] Singh, G., Mémoli, F., Carlsson, G.E., et al., 2007. Topological methods for the analysis of high dimensional data sets and 3d object recognition. Eurographics Symposium on Point-Based Graphics 2.

[42] Smith, A.M., Adler, F.R., McAuley, J.L., Gutenkunst, R.N., Ribeiro, R.M., McCullers, J.A., Perelson, A.S., 2011. Effect of 1918 PB1-F2 expression on influenza A virus infection kinetics. PLoS computational biology 7, e1001081. doi:10.1371/journal.pcbi.1001081.

[43] Storn, R., Price, K., 1997. Differential evolution–a simple and efficient heuristic for global optimization over continuous spaces. Journal of global optimization, 341–359doi:10.1023/A:1008202821328.

[44] Stovner, R.B., 2012. On the mapper algorithm: A study of a new topological method for data analysis. Master’s thesis. Institutt for Matematiske Fag.

[45] Tan, W., Lu, Y., Zhang, J., Wang, J., Dan, Y., Tan, Z., He, X., Qian, C., Sun, Q., Hu, Q., Liu, H., Ye, S., Xiang, X., Zhou, Y., Zhang, W., Guo, Y., Wang, X.H., He, W., Wan, X., Sun, F., Wei, Q., Chen, C., Pan, G., Xia, J., Mao, Q., Chen, Y., Deng, G., 2020. Viral Kinetics and Antibody Responses in Patients with COVID-19. medRxiv, 2020.03.24.20042382URL: http://medrxiv.org/content/early/2020/03/26/2020.03.24.20042382.abstract, doi:10.1101/2020.03.24.20042382.

[46] Tauzin, G., Lupo, U., Tunstall, L., Pérez, J.B., Caorsi, M., Medina-Mardones, A.M., Dassatti, A., Hess, K., 2021. giotto-tda:: A topological data analysis toolkit for machine learning and data exploration. Journal of Machine Learning Research 22, 39–1.

[47] Tay, M.Z., Poh, C.M., Rénia, L., MacAry, P.A., Ng, L.F., 2020. The trinity of COVID-19: immunity, inflammation and intervention. Nature Reviews Immunology 20, 363–374. URL: http://dx.doi.org/10.1038/s41577-020-0311-8, doi:10.1038/s41577-020-0311-8.

[48] Vanshylla, K., Di Cristanziano, V., Kleipass, F., Dewald, F., Schommers, P., Gieselmann, L., Gruell, H., Schlotz, M., Ercanoglu, M.S., Stumpf, R., et al., 2021. Kinetics and correlates of the neutralizing antibody response to sars-cov-2 infection in humans. Cell Host & Microbe 29, 917–929.

[49] Wölfel, R., Corman, V.M., Guggemos, W., Seilmaier, M., Zange, S., Müller, M.A., Niemeyer, D., Jones, T.C., Vollmar, P., Rothe, C., Hoelscher, M., Bleicker, T., Brünink, S., Schneider, J., Ehmann, R., Zwirglmaier, K., Drosten, C., Wendtner, C., 2020. Virological assessment of hospitalized patients with COVID-2019. Nature, 1–10URL: http://www.nature.com/articles/s41586-020-2196-x, doi:10.1038/s41586-020-2196-x.

