## Supplementary for "Quantifying antibody dynamics of severe and non-severe patients with COVID-19"

#### A. Bifurcation analysis

We seek an equilibrium point with virus, denoted $E^*$. To obtain a working expression for $E^*$, we introduce

$$\overline{w_M} = \frac{\overline{r_M}}{\overline{q} + \overline{\delta_M}}, \qquad \overline{w_G} = \frac{\overline{r_q}\,\overline{w_M}^2}{\overline{\delta_G}}.$$

Let us introduce the function

$$f(x,\lambda) = \left(\frac{\overline{c_G}\,\overline{w_G}}{\overline{c_T}}\right)\left[x^2 - \left(\hat{y} + x_G^*\right)x\right] + \left(\frac{\lambda + \overline{c_T}}{\overline{c_T}}\right), \tag{25}$$

$$\mathcal{H}_0(x,\lambda) = 1 - \frac{1}{f(x,\lambda)} = \frac{f(x,\lambda) - 1}{f(x,\lambda)}, \tag{26}$$

where

$$x_G^* = (\overline{w_M})^{-1} + (\overline{w_G})^{-1}, \qquad \hat{y} = \frac{\overline{c_M}\,\overline{w_M} + 1}{\overline{c_G}\,\overline{w_G}}.$$

Writing $E^* = (x^*, y^*, z^*, w_M^*, w_G^*)$ where $x^* > 0$, we establish that $E^*$ is a function of $x^*$.

**Theorem 2.** *A necessary condition for $E^*$ to exist is $\lambda + \overline{c_T} > 0$. Under this condition, $E^*$ takes the form*

$$E^* = (x^*,\ f(x^*,\lambda),\ x^*,\ \overline{w_M}x^*,\ \overline{w_G}x^*(x_G^* - x^*)) \tag{27}$$

*where $x^*$ satisfies*

$$\mathcal{H}(x^*, \overline{K_T}) = \mu\, \mathcal{H}_0(x^*, \lambda), \qquad \mu := \overline{\delta_T}/\overline{r_T}. \tag{28}$$

*Proof.* Eqs. (16)-(20) provide the following equations:

$$0 = \lambda + \overline{c_T} - x^* - \overline{c_T} y^* - \overline{c_M} w_M^* - \overline{c_G} w_G^*, \tag{29}$$

$$0 = \overline{\delta_T} + \overline{r_T} y^*\, \mathcal{H}(x^*, \overline{K_T}) - \overline{\delta_T} y^*, \tag{30}$$

$$0 = (\overline{\tau_B})^{-1}(x^* - z^*), \tag{31}$$

$$0 = \overline{r_M} z^* - (\overline{q} + \overline{\delta_M}) w_M^*, \tag{32}$$

$$0 = \overline{r_q} w_M^*(1 - w_M^*) + \overline{\delta_G}(z^* - w_G^*), \tag{33}$$

with $\lambda + \overline{c_T} = 1 - \overline{c}$. If $E^*$ exists, then we get

$$0 = \lambda + \overline{c_T} - x^* - \overline{c_T} y^* - \overline{c_M} w_M^* - \overline{c_G} w_G^* < \lambda + \overline{c_T}$$

from Eq. 29. Therefore, it is necessary that $\lambda + \overline{c_T} > 0$ for $E^*$ to exist.

Proceeding with the condition $\lambda + \overline{c_T} > 0$, we get $z^* = x^*$ from Eq (31). Now, we can rewrite Eqs. (32) and (33) as

$$0 = \overline{w_M}(\overline{q} + \overline{\delta_M}) x^* - (\overline{q} + \overline{\delta_M}) w_M^*, \tag{34}$$

$$0 = \overline{w_G}\, \overline{\delta_G} w_M^*(1 - w_M^*) + \overline{w_M}^2 \overline{\delta_G}(z^* - w_G^*). \tag{35}$$

Eq. (34) yields

$$w_M^* = \overline{w_M} x^*. \tag{36}$$

Substituting this result into Eq. (35), we obtain

$$\begin{aligned} 0 &= \overline{w_G}\, \overline{\delta_G} \overline{w_M} x^*(1 - \overline{w_M} x^*) + \overline{w_M}^2 \overline{\delta_G}(x^* - w_G^*), \\ 0 &= \overline{w_G}\, x^*(1 - \overline{w_M} x^*) + \overline{w_M}(x^* - w_G^*), \\ w_G^* &= \overline{w_G}\, x^*((\overline{w_M})^{-1} - x^*) + x^*, \end{aligned}$$

$$w_G^* = \overline{w_G}\, x^*((\overline{w_M})^{-1} + (\overline{w_G})^{-1} - x^*),$$
$$w_G^* = \overline{w_G}\, x^*(x_G^* - x^*). \tag{37}$$

From Eq. (29) we have

$$\overline{c_T}\, y^* = \lambda + \overline{c_T} - x^* - \overline{c_M} w_M^* - \overline{c_G} w_G^*. \tag{38}$$

Substituting Eqs. (37) and (36) into Eq. (38), we get

$$\overline{c_T}\, y^* = \lambda + \overline{c_T} - x^* - \overline{c_M}\, \overline{w_M} x^* - \overline{c_G}\, \overline{w_G}\, x^*(x_G^* - x^*),$$
$$\overline{c_T}\, y^* = \overline{c_G}\, \overline{w_G}(x^*)^2 - (1 + \overline{c_M}\, \overline{w_M} + \overline{c_G}\, \overline{w_G}\, x_G^*)(x^*) + \lambda + \overline{c_T},$$
$$\overline{c_T}\, y^* = \overline{c_G}\, \overline{w_G}[(x^*)^2 - (\hat{y} + x_G^*)(x^*)] + \lambda + \overline{c_T}.$$

The last equation is equivalent to $y^* = f(x^*, \lambda)$. Therefore, $E^*$ takes the form in Eq. (27). Moreover, taking $y^* = f(x^*, \lambda)$ in Eq. (30) yields:

$$0 = \overline{\delta_T} + \overline{r_T} f(x^*, \lambda)\, \mathcal{H}(x^*, \overline{K_T}) - \overline{\delta_T} f(x^*, \lambda),$$
$$\mathcal{H}(x^*, \overline{K_T}) = \frac{\overline{\delta_T} f(x^*, \lambda) - \overline{\delta_T}}{\overline{r_T} f(x^*, \lambda)},$$

which is Eq. (28). □

**Theorem 3.** *Assume that $\lambda + \overline{c_T} > 0$ and let $\mathcal{H}_G^* := \mathcal{H}(x_G^*, \overline{K_T})$. Let $\mathcal{I}$ be the set of all $x$ such that*

- $0 < x \leq x_G^*$,
- $f(x, \lambda) > 1$, *and*
- *either $\mu \leq \mathcal{H}_G^*$ or $f(x, \lambda) \leq \frac{\mu}{\mu - \mathcal{H}_G^*}$.*

*Then every virus-positive equilibrium point $E^*$ has the viral load $x^* \in \mathcal{I}$.*

*Proof.* Eq. (27) in Theorem 2 requires that $f(x^*, \lambda) > 0$ and $0 < x^* \leq x_G^*$. Since $\mathcal{H}(x, \lambda)$ increases over positive values of $x$, we have

$$0 < \mathcal{H}(x^*, \lambda) \leq \mathcal{H}_G^*$$
$$\iff 0 < \mathcal{H}_0(x^*, \lambda) \leq \frac{\mathcal{H}_G^*}{\mu} \qquad \text{by Eq. (28)}$$

$$\iff 0 < \frac{f(x^*, \lambda) - 1}{f(x^*, \lambda)} \leq \frac{\mathcal{H}_G^*}{\mu} \qquad \text{by Eq. (26)}$$
$$\iff 1 < f(x^*, \lambda) \leq 1 + \frac{\mathcal{H}_G^*}{\mu} f(x^*, \lambda)$$

The last inequality is equivalent to

$$f(x^*, \lambda) \left[1 - \frac{\mathcal{H}_G^*}{\mu}\right] \leq 1 < f(x^*, \lambda),$$

or

$$f(x^*, \lambda) > 1, \text{ and either } \mu \leq \mathcal{H}_G^* \text{ or } f(x^*, \lambda) \leq \frac{\mu}{\mu - \mathcal{H}_G^*}.$$

Therefore, $x \in \mathcal{I}$. □

The set $\mathcal{I}$ in Theorem 3 locates all possible values of the positive viral load $x^*$ at $E^*$. Note that this set may take different forms according to $\lambda$ and $\mu$. Both quantities put bound restrictions on $f(x, \lambda)$ as follows:

- The parameter $\lambda$ restricts $f(x, \lambda)$ with $f(x, \lambda) > 1$, which is needed for both $y^* > 0$ and $\mathcal{H}_0(x^*) > 0$.
- If $\mu > \mathcal{H}_G^*$, then $\mu$ puts an additional restriction: $f(x, \lambda) \leq \mu/(\mu - \mathcal{H}_G^*)$.

To determine these forms, we collect some properties of $f$ as a quadratic function in $x$ restricted to the interval $(0, x_G^*]$.

1. We have $f(\hat{y}, \lambda) = f(x_G^*, \lambda) < f(0, \lambda)$ due to

$$f(0, \lambda) = \left(\frac{\lambda + \overline{c_T}}{\overline{c_T}}\right),$$
$$f(x_G^*, \lambda) = -\left(\frac{\overline{c_G}\,\overline{w_G}}{\overline{c_T}}\right)(\hat{y} x_G^*) + \left(\frac{\lambda + \overline{c_T}}{\overline{c_T}}\right).$$

2. The graph of $y = f(x, \lambda)$ in the $xy$-plane is concave up, due to the leading coefficient $(\overline{c_G}\,\overline{w_G}/\overline{c_T})$.
3. The same graph also has its vertex point $(\hat{x}_0, f_{\min})$, where

$$\hat{x}_0 := \frac{\hat{y} + x_G^*}{2}, \qquad f_{\min} := f(\hat{x}_0, \lambda) = f(0, \lambda) - \left(\frac{\overline{c_G}\,\overline{w_G}}{\overline{c_T}}\right)\hat{x}_0^2.$$

   Thus, $f(x, \lambda)$ decreases with negative $\frac{\partial f}{\partial x}$ for $x < \hat{x}_0$ and increases with positive $\frac{\partial f}{\partial x}$ for $x > \hat{x}_0$.
4. We have $2(\hat{x}_0 - x_G^*) = \hat{y} - x_G^*$, hence $\text{sgn}(\hat{x}_0 - x_G^*) = \text{sgn}(\hat{y} - x_G^*)$. Moreover, $\hat{x}_0 \leq x_G^*$ if $\hat{y} \leq x_G^*$.

5. Finally, the quadratic formula gives the following roots of $f(\,\cdot\,,\lambda) - 1$:

$$\widehat{x}_1 := \widehat{x}_0 - \sqrt{\widehat{x}_0^2 - \frac{\overline{c_T}}{\overline{c_G}\,\overline{w_G}}\,[f(0,\lambda) - 1]},$$
$$\widehat{x}_2 := \widehat{x}_0 + \sqrt{\widehat{x}_0^2 - \frac{\overline{c_T}}{\overline{c_G}\,\overline{w_G}}\,[f(0,\lambda) - 1]}.$$

Note that $\widehat{x}_1 \leq \widehat{x}_2$.

For a simpler analytical approach, we may consider the following set

$$\mathcal{I}_0 := \{x \mid 0 < x \leq x_G^* \text{ and } f(x,\lambda) > 1\},$$
$$\implies \mathcal{I} = \mathcal{I}_0 \cap \left\{ x \mid \mu \leq \mathcal{H}_G^* \text{ or } f(x,\lambda) \leq \frac{\mu}{\mu - \mathcal{H}_G^*} \right\}.$$

Then Theorem 3 implies that $E^* \in \mathcal{I}_0$. That is, $\mathcal{I}_0$ provides regions to locate $E^*$, while $\mathcal{I}$ provides additional restriction.

To determine the possible sets that $\mathcal{I}_0$ takes, we appeal to the monotone and concave properties of $f(\,\cdot\,,\lambda)$, particularly that $f(x,\lambda)$ decreases for $x < \widehat{x}_0$ and increases for $x > \widehat{x}_0$. Consider partitioning the parameter space into the following cases.

- Case 1: $f(0,\lambda) \leq 1$.
  If $\widehat{x}_0 > x_G^*$, then $f(x,\lambda)$ decreases with $x$ over $(0, x_G^*]$. Otherwise, the image of $(0, x_G^*]$ under $f(\,\cdot\,,\lambda)$ is $[f_{\min}, f(0,\lambda)) \subset (-\infty, 1)$. Therefore, $\mathcal{I}_0$ is empty.

- Case 2: $f(x_G^*, \lambda) \leq 1 < f(0,\lambda)$.
  Restricting $f(x,\lambda)$ for $0 < x < x_G^*$ and following the same cases for $\widehat{x}_0$, we see that $f(x,\lambda) = 1$ uniquely at $x = \widehat{x}_1$. Moreover, $\mathcal{I}_0 = (0, \widehat{x}_1)$, with $f(x,\lambda) > 1$ for $0 < x < \widehat{x}_1$, and $f(x,\lambda) < 1$ for $\widehat{x}_1 < x \leq x_G^*$.

- Case 3: $f(x_G^*, \lambda) > 1$, and either $\widehat{y} \geq x_G^*$ or $f_{\min} > 1$.
  Note in this case that $f(0,\lambda) > f(x_G^*,\lambda) > 1$. If $\widehat{y} \geq x_G^*$ (equivalently, $\widehat{x}_0 \geq x_G^*$), then as $x$ increases from zero to $x_G^*$, the value of $f$ decreases from $f(0,\lambda)$ to $f(x_G^*,\lambda) > 1$. Otherwise, with $f_{\min} > 1$, the image of $(0, x_G^*]$ under $f(\,\cdot\,,\lambda)$ is $[f_{\min}, f(0,\lambda)) \supset (1,\infty)$. Therefore, $\mathcal{I}_0 = (0, x_G^*]$.

- Case 4: $f(x_G^*, \lambda) > 1$, $\widehat{y} < x_G^*$ (equivalently $\widehat{x}_0 < x_G^*$), and $f_{\min} \leq 1$.
  Here, both $f(0,\lambda)$ and $f(x_G^*,\lambda)$ are above one, but $f_{\min} \leq 1$. Thus, the roots $\widehat{x}_1$ and $\widehat{x}_2$ of $f(\,\cdot\,,\lambda)$ exist, with $\widehat{x}_1 \leq \widehat{x}_0 \leq \widehat{x}_2$. Since $f(x,\lambda)$ decreases for $x < \widehat{x}_0$ and increases for $x > \widehat{x}_0$, we conclude that $\mathcal{I}_0 = (0, \widehat{x}_1) \cup (\widehat{x}_2, x_G^*]$.

**Table S1**
Different cases for the set $\mathcal{I}_0$. All cases assume that $\lambda > -\overline{c_T}$, which is necessary for $E^*$ to exist (Theorem 2).

| Case | Conditions | Set $\mathcal{I}_0$ |
|---|---|---|
| 1 | $-\overline{c_T} < \lambda \leq 0$ | Empty |
| 2 | $0 < \lambda \leq \Lambda_1$ | $(0, \hat{x}_1)$ |
| 3 | $\lambda > \Lambda_1$, and either $\hat{x}_0 \geq x_G^*$ or $\lambda > \Lambda_2$ | $(0, x_G^*]$ |
| 4 | $\hat{x}_0 < x_G^*$ and $\Lambda_1 < \lambda \leq \Lambda_2$ | $(0, \hat{x}_1) \cup (\hat{x}_2, x_G^*]$ |

We may express the conditions of the four cases in $\lambda$ by writing

$$\begin{aligned} f(0,\lambda) - 1 &= \lambda(\overline{c_T})^{-1}, \\ f(x_G^*,\lambda) - 1 &= \frac{\lambda - \Lambda_1}{\overline{c_T}}, \\ f_{\min} - 1 &= \frac{\lambda - \Lambda_2}{\overline{c_T}}, \end{aligned}$$

where

$$\begin{aligned} \Lambda_1 &:= (\overline{c_G}\,\overline{w_G})(2\hat{x}_0 - x_G^*)\,x_G^* = (\overline{c_G}\,\overline{w_G})\,\hat{y}\,x_G^*, \\ \Lambda_2 &:= (\overline{c_G}\,\overline{w_G})\,\hat{x}_0^2. \end{aligned}$$

These yield the following sign equations

$$\begin{aligned} \operatorname{sgn}[f(0,\lambda) - 1] &= \operatorname{sgn}\lambda, \\ \operatorname{sgn}[f(x_G^*,\lambda) - 1] &= \operatorname{sgn}(\lambda - \Lambda_1), \\ \operatorname{sgn}(f_{\min} - 1) &= \operatorname{sgn}(\lambda - \Lambda_2), \end{aligned}$$

in addition to $\operatorname{sgn}(\hat{x}_0 - x_G^*) = \operatorname{sgn}(\hat{y} - x_G^*)$. Table S1 summarizes our analysis on the different intervals (or union of intervals) that $\mathcal{I}_0$ takes.

We now investigate the equation

$$\mathcal{H}(\,\cdot\,, \overline{K_T}) = \mu\,\mathcal{H}_0(\,\cdot\,, \lambda) \text{ on } \mathcal{I}_0 \tag{39}$$

from Eq. (28), where the root(s) determine the viral load size(s) $x^*$ at $E^*$. We have

$$\frac{\partial \mathcal{H}}{\partial x} = \frac{m\overline{K_T}^m \cdot x^{m-1}}{(x^m + \overline{K_T}^m)^2} > 0 \qquad \text{for } x > 0, \tag{40}$$

so $\mathcal{H}(\,\cdot\,,\overline{K_T})$ increases over $(0,x^*_G]$ and particularly over $\mathcal{I}_0$. On the other hand, we can determine the monotone properties of $\mathcal{H}_0(\,\cdot\,,\lambda)$ by applying Eqs. (25) and (26), assuming that $f>0$. Now,

$$\begin{aligned}\frac{\partial f}{\partial x} &= \left(\frac{\overline{c_G}\,\overline{w_G}}{\overline{c_T}}\right)\left[2x-\left(\widehat{y}+x^*_G\right)\right]\\ &= 2\left(\frac{\overline{c_G}\,\overline{w_G}}{\overline{c_T}}\right)(x-\widehat{x}_0).\end{aligned} \tag{41}$$

Then

$$\begin{aligned}\frac{\partial \mathcal{H}_0}{\partial x} &= \frac{\partial}{\partial x}\left(1-\frac{1}{f}\right)=\frac{1}{f^2}\frac{\partial f}{\partial x}\\ &= \frac{2}{[f(x,\lambda)]^2}\left(\frac{\overline{c_G}\,\overline{w_G}}{\overline{c_T}}\right)(x-\widehat{x}_0).\end{aligned} \tag{42}$$

For concavity, we may compute the following second partial derivative:

$$\begin{aligned}\frac{\partial^2 \mathcal{H}_0}{\partial x^2} &= \frac{2}{[f(x,\lambda)]^3}\left(\frac{\overline{c_G}\,\overline{w_G}}{\overline{c_T}}\right)\left[2(\widehat{x}_0-x)\frac{\partial f}{\partial x}+f\right]\\ &= \frac{2}{[f(x,\lambda)]^3}\left(\frac{\overline{c_G}\,\overline{w_G}}{\overline{c_T}}\right)\left[f(x,\lambda)-4\left(\frac{\overline{c_G}\,\overline{w_G}}{\overline{c_T}}\right)(x-\widehat{x}_0)^2\right]\\ &= \frac{8}{[f(x,\lambda)]^3}\left(\frac{\overline{c_G}\,\overline{w_G}}{\overline{c_T}}\right)^2\left[\frac{1}{4}\left(\frac{\overline{c_T}}{\overline{c_G}\,\overline{w_G}}\right)f(x,\lambda)-(x-\widehat{x}_0)^2\right].\end{aligned} \tag{43}$$

Since $f(\,\cdot\,,\lambda)>1>0$ on $\mathcal{I}_0$, Eqs. (41)-(43) yield the following sign equations for all $x\in\mathcal{I}_0$:

$$\operatorname{sgn}\frac{\partial \mathcal{H}_0}{\partial x} = \operatorname{sgn}(x-\widehat{x}_0)=\operatorname{sgn}\frac{\partial f}{\partial x}, \tag{44}$$

$$\operatorname{sgn}\frac{\partial^2 \mathcal{H}_0}{\partial x^2} = \operatorname{sgn}\left[\frac{1}{4}\left(\frac{\overline{c_T}}{\overline{c_G}\,\overline{w_G}}\right)f(x,\lambda)-(x-\widehat{x}_0)^2\right]. \tag{45}$$

In particular, both $\mathcal{H}_0(\,\cdot\,,\lambda)$ and $f(\,\cdot\,,\lambda)$ share the same monotone properties over $\mathcal{I}_0$. We also note that

$$\operatorname{sgn}\mathcal{H}_0(0,\lambda)=\operatorname{sgn}[f(0,\lambda)-1]$$

Armed with these observations, we revisit Cases 2-4 in Table S1. We skip Case 1 because $\mathcal{I}_0$ is empty and thus $E^*$ does not exist. For Cases 2-4, $f(0,\lambda)>1$ and $\mathcal{H}_0(0,\lambda)>0$. Recalling $\mu=\overline{\delta_T}/\overline{r_T}$, the signs of $\mathcal{H}_0$, $\mu\,\mathcal{H}_0$ and their partial derivatives are equal. Eq. (28), we also have

- Under Case 2, we have $\mathcal{I}_0=(0,\widehat{x}_1)\subseteq(0,\widehat{x}_0)$. Hence by eq. (42), $\mathcal{H}_0(\,\cdot\,,\lambda)$ decreases over $\mathcal{I}_0$. Considering eq. (40) and that

$$\mu\,\mathcal{H}_0(\hat{x}_1, \lambda) = 0 < \mu\,\mathcal{H}_0(0, \lambda),$$

Eq. (39) admits exactly one solution $x^*$.

- Under Case 3, we now have $\mathcal{I}_0 = (0, x_G^*]$ so that $\mathcal{H}_0(x, \lambda) > 0$ for $0 < x \le x_G^*$. We proceed with the following subcases:

  - Subcase 3a. If $\hat{x}_0 \ge x_G^*$, then according to Eq. (44) we have $\frac{\partial \mathcal{H}_0}{\partial x} < 0$ and $\mathcal{H}_0(\,\cdot\,, \lambda)$ decreases over $\mathcal{I}_0$. Furthermore, eq. (39) admits a solution if and only if

    $$\mu\,\mathcal{H}_0(x_G^*, \lambda) \le \mathcal{H}(x_G^*, \overline{K_T}) = \mathcal{H}_G^*.$$

    This solution is unique due to the decreasing property of $\mathcal{H}_0(\,\cdot\,, \lambda)$.

  - Subcase 3b. If $0 < \hat{x} < x_G^*$, then $\mathcal{H}_0(\,\cdot\,, \lambda)$ decreases over $(0, \hat{x}_0)$ and increases over $(\hat{x}_0, x_G^*)$. Similar to the Subcase 3a, we conclude that Eq. (39) *restricted to* $(0, \hat{x}_0)$ has a solution (which is unique) if and only if

    $$\mu\,\mathcal{H}_0(\hat{x}, \lambda) \le \mathcal{H}(\hat{x}, \overline{K_T}).$$

    Over $(\hat{x}_0, x_G^*]$, the behavior of Eq. (39) may be intricate due to the changing convexities of both functions $\mathcal{H}(\,\cdot\,, \overline{K_T})$ and $\mu\,\mathcal{H}_0(\cdot, \lambda)$. Future work may need to determine possible numbers of roots of Eq. eq. (39) over $(\hat{x}_0, x_G^*]$.

- Finally, we have $\mathcal{I}_0 = (0, \hat{x}_1) \cup (\hat{x}_2, x_G^*]$ (disjoint union of intervals) in Case 4. Similar to Case 1, we find that Eq. (39) attains a unique solution over $(0, \hat{x}_1)$. Similar to Case 3b, we might expect the same kind of intricate behavior of Eq. (39) over $(\hat{x}_2, x_G^*]$ depending on $\mu$ and the concave properties of $\mathcal{H}$ and $\mathcal{H}_0$.

We are ready to consolidate our analysis on the existence of virus-positive equilibria. Anticipating the possibility of more than one virus-positive equilibrium points, we may write $\mathcal{H} - \mu\,\mathcal{H}_0$ as a rational function in $x$, where both the numerator and the denominator are polynomials in $x$ of degree $m+2$. Hence, we deduce that the equation $\mathcal{H}(\,\cdot\,, \overline{K_T}) = \mu\,\mathcal{H}_0(\,\cdot\,, \lambda)$ admits at most $m + 2$ distinct roots in $\mathcal{I}_0$. We will denote these roots as $x_1^*$, $x_2^*$, ..., $x_\ell^*$, where $\ell \le m + 2$ an $x_j < x_k$ for $j < k$ (provided both $x_j$ and $x_k$ exist). These roots correspond to the existence of the virus-positive equilibrium points

$$E_k^* = (x_k^*, f(x_k^*, \lambda), x_k^*, \overline{w_M} x_k^*, \overline{w_G} x_k^*(x_G^* - x_k^*)), \qquad k = 1, 2, \ldots, \ell.$$

The number $\ell$ determines how many virus-positive equilibrium points exist, so that $\ell = 0$ means that the only equilibrium point is virus-free (i.e., $E_0$). In the simplest case where $m = 2$, we can expect up to four virus-positive equilibrium points ($\ell \leq 4$).

We consolidate our preceding analysis, especially Table S1 and Theorems 2 and 3, into the following summary.

**Theorem 4.** *Dimensionless Model 6, Eqs.* (16)-(20)*, admits virus-positive equilibrium points only if* $\lambda > 0$*. Under this condition, we have the following cases:*

*(a) If* $0 < \lambda \leq \Lambda_1$*, then* $\ell = 1$ *and* $E_1^*$ *is the unique virus-positive equilibrium point. Furthermore,* $x_1^* \in (0, \hat{x}_1)$.

*(b) If* $\lambda > \Lambda_1$ *and* $\hat{x}_0 \geq x_G^*$*, then* $\ell = 1$ *and the virus-positive equilibrium point* $E_1^*$ *exists with* $x_1^* \in (0, x_G^*]$.

*(c) Suppose that* $\lambda > \Lambda_1$ *but* $\hat{x}_0 < x_G^*$*, and* $\lambda > \Lambda_2$*. Then* $E_k^*$ *exists with* $x_k^* \in (\hat{x}_0, x_G^*]$ *for* $k = 2, \ldots, \ell$*. Moreover:*

- *If* $\mu \mathcal{H}_0(\hat{x}, \lambda) \leq \mathcal{H}(\hat{x}, \overline{K_T})$*, then* $x_1^* \in (0, \hat{x}_0)$ *and* $\ell \geq 1$.
- *Otherwise,* $x_1^* \in (\hat{x}_0, x_G^*]$ *and we can only conclude that* $\ell \geq 0$*. Here, the value of* $\ell$ *may depend on the concavity of* $\mathcal{H}$ *and* $\mu \mathcal{H}_0$.

*(d) Suppose that* $\hat{x}_0 < x_G^*$ *and* $\Lambda_1 < \lambda \leq \Lambda_2$*. Then* $\ell \geq 1$ *and* $E_1^*$ *exists with* $x_1^* \in (0, \hat{x}_1)$*. Moreover,* $E_k^*$ *exists with* $x_k^* \in (\hat{x}_2, x_G^*]$ *for* $k = 2, \ldots, \ell$.

*Remark.* Further analysis, either through numerical exploration or advanced mathematical tools, is needed in the following cases:

- $\lambda > \max(\Lambda_1, \Lambda_2)$ and $\hat{x}_0 < x_G^*$.
- $\hat{x}_0 < x_G^*$ and $\Lambda_1 < \lambda \leq \Lambda_2$, only for the equilibrium points $E_2^*$, ..., $E_\ell^*$.

### B. Math Modelling

#### B.1. Model 2

$$
\begin{aligned}
\frac{dV}{dt} &= pV\left(1-\frac{V}{K_V}\right)-c_tVT-\varepsilon MV-\eta GV-cV, && (46)\\
\frac{dT}{dt} &= \delta_t T(0)+rT\left(\frac{V^m}{V^m+k_t^m}\right)-\delta_t T, && (47)\\
\frac{dM}{dt} &= \alpha V(t-\tau_1)-(\gamma+\beta)M, && (48)\\
\frac{dG}{dt} &= \gamma M+\lambda V(t-\tau_2)-\rho G. && (49)
\end{aligned}
$$

#### B.2. Model 4

$$
\begin{aligned}
\frac{dV}{dt} &= pV\left(1-\frac{V}{K_V}\right)-c_tVT-\varepsilon MV-\eta GV-cV, && (50)\\
\frac{dT}{dt} &= \delta_t T(0)+rT\left(\frac{V^m}{V^m+k_t^m}\right)-\delta_t T, && (51)\\
\frac{dB}{dt} &= r_b\left(\frac{1}{1+l_vV}\right)\left(\frac{1}{1+l_tT}\right)(1-B), && (52)\\
\frac{dM}{dt} &= r_mB-(q+\delta_m)M, && (53)\\
\frac{dG}{dt} &= qM+r_gB-\delta_g G. && (54)
\end{aligned}
$$

#### B.3. Model 5

$$
\begin{aligned}
\frac{dV}{dt} &= pV\left(1-\frac{V}{K_V}\right)-c_tVT-\varepsilon MV-\eta GV-cV, && (55)\\
\frac{dT}{dt} &= \delta_t T(0)+rT\left(\frac{V^m}{V^m+k_t^m}\right)-\delta_t T, && (56)\\
\frac{dB}{dt} &= \frac{\delta_v V-B}{\tau_b}, && (57)\\
\frac{dM}{dt} &= r_mB-(q+\delta_m)M, && (58)\\
\frac{dG}{dt} &= q\left(\frac{M^n}{M^n+k_q^n}\right)+r_gB-\delta_g G. && (59)
\end{aligned}
$$

#### C. AIC scores for each severe patients

**Table S2**
AIC scores for severe patients

| Patient | AIC Model 1 | AIC Model 2 | AIC Model 3 | AIC Model 4 | AIC Model 5 | AIC Model 6 |
|---|---|---|---|---|---|---|
| 2 | 7.32 | -5.74 | -13.24 | -42.57 | -57.92 | -63.02 |
| 3 | 35.86 | 12.52 | 23.75 | -12.63 | -36.84 | -15.46 |
| 5 | 21.08 | -6.24 | 1.32 | -32.61 | -11.72 | -28.25 |
| 12 | 14.63 | -1.02 | -32.70 | -30.84 | -20.93 | -18.76 |
| 15 | 27.37 | -4.27 | -14.59 | -19.52 | -5.96 | -19.13 |
| 20 | 42.60 | 4.85 | 4.85 | -31.08 | -29.41 | -9.26 |
| 34 | 16.93 | 7.29 | -1.19 | -28.85 | -25.74 | -34.49 |
| Media | 23.68 | 1.05 | -4.54 | -28.30 | -26.93 | -26.91 |

#### D. AIC scores for each non-severe patients

**Table S3**
AIC scores for each non-severe patients

| Patient | AIC Model 1 | AIC Model 2 | AIC Model 3 | AIC Model 4 | AIC Model 5 | AIC Model 6 |
|---|---|---|---|---|---|---|
| 1 | 32.57 | 25.76 | 6.23 | 4.84 | -36.1 | -13.65 |
| 4 | 4.56 | 13.57 | -20.36 | -59.27 | -24.82 | -166.21 |
| 7 | 57.34 | 34.08 | 5.25 | -8.29 | -3.73 | -13.32 |
| 8 | 25.78 | 11.41 | -16.39 | -2.04 | -14.42 | -24.88 |
| 9 | 9.47 | -2.69 | -18.87 | 4.06 | -25.38 | -29.18 |
| 13 | -5.12 | 7.63 | -48.31 | -7.49 | -16.11 | -61.75 |
| 14 | 12.45 | 19.54 | 3.48 | -8.16 | -17.93 | -36.27 |
| 16 | 41.63 | 23.97 | 4.72 | -2.93 | 11.03 | -9.49 |
| 17 | 6.28 | 25.06 | -1.38 | -17.26 | -6.49 | -19.32 |
| 18 | -3.93 | 17.42 | -19.65 | -11.42 | -28.53 | -25.86 |
| 19 | 5.84 | 19.70 | -3.27 | -2.98 | -26.77 | -14.09 |
| 21 | 34.72 | 2.86 | -25.93 | -36.07 | -21.53 | -48.30 |
| 22 | 23.67 | -4.84 | -2.21 | -12.30 | -9.64 | -7.38 |
| 24 | 11.63 | 10.65 | -23.74 | -2.04 | -35.23 | -27.10 |
| 25 | -7.32 | 38.32 | -43.91 | -55.72 | -85.41 | -173.52 |
| 26 | 17.48 | 3.05 | -16.01 | -28.04 | -21.63 | -37.16 |
| 27 | 9.37 | 23.94 | -2.72 | 2.63 | -37.50 | -18.56 |
| 29 | 17.82 | -7.13 | 12.54 | -28.42 | -26.94 | -30.22 |
| 31 | 16.21 | 4.21 | 3.90 | -12.63 | -33.71 | -45.27 |
| 35 | 8.84 | -12.74 | 11.34 | -27.75 | -52.02 | -63.34 |
| 36 | 19.36 | -18.95 | 2.45 | -9.03 | -5.94 | -12.89 |
| 37 | 26.21 | 11.62 | -47.25 | -2.49 | -16.83 | -6.81 |
| 38 | 21.45 | 4.29 | -15.68 | -24.23 | -26.56 | -30.18 |
| 39 | 12.87 | -2.16 | -14.63 | -9.62 | -8.34 | -24.02 |
| 40 | 35.97 | -3.59 | 2.03 | -2.73 | 3.65 | -7.93 |
| 42 | 23.92 | -1.08 | 4.78 | -8.38 | -36.7 | -6.71 |
| 53 | 31.37 | -8.60 | -2.47 | -23.52 | -21.26 | -24.92 |
| 57 | 48.93 | -2.57 | 11.19 | -2.96 | -7.40 | -9.70 |
| 59 | 27.80 | 4.89 | 1.53 | -13.06 | -2.87 | -19.18 |
| 61 | 15.04 | 3.86 | -23.64 | -5.14 | -27.81 | -31.70 |
| 63 | 48.52 | -4.43 | -9.05 | 2.85 | -42.43 | -20.56 |
| 64 | 4.63 | -10.86 | -14.32 | -20.45 | -8.92 | -17.33 |
| Media | 19.85 | 7.06 | -9.38 | -13.43 | -22.32 | -33.65 |

#### E. Estimated parameters of Model 6 for each severe patients

**Table S4**

Estimated parameters for each severe patients

| Patients | $c_M$ | $c_G$ | $\tau_b$ | $r_M$ | $q$ | $r_G$ | $K_q$ | $r_q$ | $r_B$ |
|---|---|---|---|---|---|---|---|---|---|
| 2 | 1.24E-04 | 1.20E-04 | 2.95E+00 | 5.00E-06 | 1.00E-01 | 7.00E-06 | 2.35E+01 | 1.05E-01 | 2.33E-01 |
| 3 | 6.06E-03 | 2.93E-01 | 1.96E+01 | 1.92E-04 | 5.27E-02 | 4.77E-04 | 9.19E+00 | 7.76E+00 | 2.34E-01 |
| 5 | 2.13E-03 | 1.77E-01 | 3.23E+01 | 1.00E-06 | 7.88E-04 | 1.00E-06 | 2.13E+02 | 1.71E-01 | 1.42E-01 |
| 12 | 1.53E-03 | 1.82E-02 | 4.72E+00 | 6.64E-07 | 9.97E-02 | 1.00E-09 | 3.68E+02 | 1.76E-01 | 1.93E-01 |
| 15 | 3.35E-03 | 2.78E-02 | 3.99E+01 | 4.00E-06 | 1.35E-03 | 1.00E-09 | 3.88E+02 | 6.88E-02 | 2.17E-01 |
| 20 | 7.81E-03 | 2.40E-02 | 3.98E+01 | 2.00E-06 | 9.79E-04 | 1.00E-09 | 3.90E+02 | 1.60E-01 | 2.36E-01 |
| 34 | 8.98E-03 | 2.81E-01 | 1.50E+00 | 3.37E-04 | 7.32E-02 | 8.07E-03 | 3.74E+02 | 2.31E+01 | 1.34E-01 |

#### F. Estimated parameters of Model 6 for each non-severe patients

**Table S5**

Estimated parameters for each non-severe patients

| Patients | $c_M$ | $c_G$ | $\tau_b$ | $r_M$ | $q$ | $r_G$ | $K_q$ | $r_q$ | $r_B$ |
|---|---|---|---|---|---|---|---|---|---|
| 1 | 6.04E-03 | 4.08E-02 | 6.48E+00 | 1.25E-03 | 3.34E-03 | 3.03E-04 | 3.66E+02 | 9.19E-03 | 1.05E-01 |
| 4 | 9.39E-03 | 2.71E-04 | 2.56E+00 | 1.00E-09 | 8.57E-02 | 8.79E-07 | 1.02E+02 | 1.96E-02 | 1.19E-01 |
| 7 | 7.43E-03 | 2.09E-03 | 1.09E+01 | 4.75E-08 | 1.20E-04 | 4.07E-07 | 8.20E+01 | 1.04E-02 | 2.90E-01 |
| 8 | 5.11E-03 | 2.45E-01 | 3.97E+01 | 9.79E-09 | 4.01E-02 | 6.53E-07 | 9.39E+01 | 1.37E+00 | 1.10E-01 |
| 9 | 6.68E-03 | 1.47E-01 | 3.15E+01 | 2.86E-08 | 1.11E-03 | 3.00E-06 | 1.75E+02 | 2.74E-01 | 1.53E-01 |
| 13 | 9.46E-03 | 1.77E-02 | 6.68E+00 | 6.78E-07 | 4.62E-02 | 1.00E-09 | 3.82E+02 | 6.13E-01 | 1.55E-01 |
| 14 | 2.85E-03 | 7.11E-02 | 1.52E+01 | 1.00E-06 | 1.70E-03 | 1.00E-09 | 3.32E+02 | 1.55E-01 | 2.02E-01 |
| 16 | 4.92E-04 | 8.09E-03 | 3.97E+01 | 5.00E-06 | 2.78E-03 | 1.00E-09 | 3.96E+02 | 1.14E-01 | 2.19E-01 |
| 17 | 2.63E-04 | 1.45E-02 | 6.34E+00 | 7.25E-07 | 7.06E-02 | 1.00E-09 | 2.51E+02 | 5.64E-01 | 1.60E-01 |
| 18 | 4.30E-03 | 8.19E-02 | 3.97E+01 | 8.08E-07 | 4.04E-03 | 1.00E-09 | 3.80E+02 | 9.63E-02 | 9.18E-01 |
| 19 | 3.43E-04 | 2.16E-01 | 3.98E+01 | 8.57E-07 | 9.20E-04 | 1.00E-09 | 3.87E+02 | 3.91E-02 | 5.83E-01 |
| 21 | 7.83E-04 | 2.90E-01 | 2.18E+01 | 1.00E-06 | 3.86E-03 | 1.00E-09 | 3.95E+02 | 2.37E-02 | 3.15E-01 |
| 22 | 5.56E-03 | 2.87E-01 | 3.88E+01 | 6.79E-07 | 2.68E-03 | 1.00E-09 | 3.97E+02 | 1.78E-02 | 7.56E-01 |
| 24 | 6.00E-03 | 7.21E-02 | 8.25E+00 | 9.00E-07 | 5.80E-02 | 1.00E-09 | 3.29E+02 | 1.11E-01 | 1.60E-01 |
| 25 | 7.20E-03 | 2.66E-01 | 3.70E+01 | 6.32E-08 | 9.93E-02 | 1.06E-09 | 3.89E+02 | 1.34E+00 | 1.03E-01 |
| 26 | 8.18E-03 | 2.51E-01 | 3.99E+01 | 1.32E-07 | 7.82E-04 | 1.29E-09 | 2.79E+02 | 3.02E-01 | 1.02E-01 |
| 27 | 2.44E-03 | 4.75E-03 | 1.07E+01 | 4.69E-07 | 3.28E-02 | 1.05E-09 | 3.96E+02 | 7.59E-02 | 4.62E-01 |
| 29 | 3.05E-03 | 1.50E-01 | 3.49E+01 | 2.24E-09 | 4.85E-02 | 8.73E-08 | 9.60E+01 | 1.38E+00 | 1.27E-01 |
| 31 | 6.38E-03 | 2.88E-01 | 2.37E+00 | 1.63E-09 | 2.31E-02 | 2.31E-07 | 1.66E+02 | 5.10E+00 | 1.09E-01 |
| 35 | 8.11E-03 | 5.70E-04 | 2.21E+00 | 4.79E-08 | 1.15E-02 | 3.00E-06 | 3.33E+02 | 2.34E+00 | 1.16E-01 |
| 36 | 4.48E-03 | 3.00E-01 | 1.00E+00 | 1.00E-09 | 1.00E-01 | 1.00E-02 | 3.03E+02 | 2.27E+01 | 7.82E-01 |
| 37 | 1.92E-03 | 3.95E-04 | 3.16E+01 | 4.10E-05 | 6.75E-03 | 9.37E-08 | 9.19E+01 | 1.10E+00 | 1.42E-01 |
| 38 | 2.21E-03 | 1.55E-01 | 3.35E+01 | 8.76E-08 | 2.56E-02 | 3.23E-08 | 2.05E+02 | 5.64E-01 | 3.62E-01 |
| 39 | 7.75E-03 | 1.93E-01 | 3.18E+01 | 2.00E-06 | 9.60E-02 | 2.00E-06 | 1.66E+02 | 1.68E+00 | 1.15E-01 |
| 40 | 3.09E-03 | 1.56E-01 | 1.00E+00 | 2.00E-05 | 1.00E-01 | 2.60E-05 | 3.69E+02 | 1.12E+01 | 1.06E-01 |
| 42 | 4.11E-04 | 5.19E-04 | 3.99E+01 | 8.00E-06 | 2.14E-03 | 1.00E-09 | 3.99E+02 | 5.09E-01 | 1.85E-01 |
| 53 | 9.56E-03 | 1.09E-01 | 3.81E+01 | 5.77E-09 | 9.99E-02 | 1.55E-09 | 3.76E+02 | 1.75E+00 | 1.15E-01 |
| 57 | 1.00E-02 | 3.00E-01 | 1.00E+00 | 6.50E-03 | 1.00E-01 | 1.00E-02 | 1.78E+02 | 4.00E+01 | 1.00E-01 |
| 59 | 8.97E-03 | 2.04E-01 | 1.12E+00 | 1.00E-09 | 1.00E-01 | 7.93E-04 | 3.23E+02 | 3.08E+01 | 1.12E-01 |
| 61 | 7.89E-03 | 2.70E-01 | 3.04E+01 | 3.94E-08 | 5.35E-02 | 4.97E-07 | 3.39E+02 | 3.49E+00 | 1.05E-01 |
| 63 | 4.09E-03 | 5.71E-03 | 3.18E+00 | 1.00E-06 | 1.94E-03 | 6.21E-07 | 1.80E+02 | 5.04E-01 | 1.24E-01 |
| 64 | 8.80E-03 | 2.95E-01 | 4.08E+00 | 1.14E-03 | 3.14E-02 | 7.29E-04 | 4.81E+00 | 9.86E+00 | 1.48E-01 |

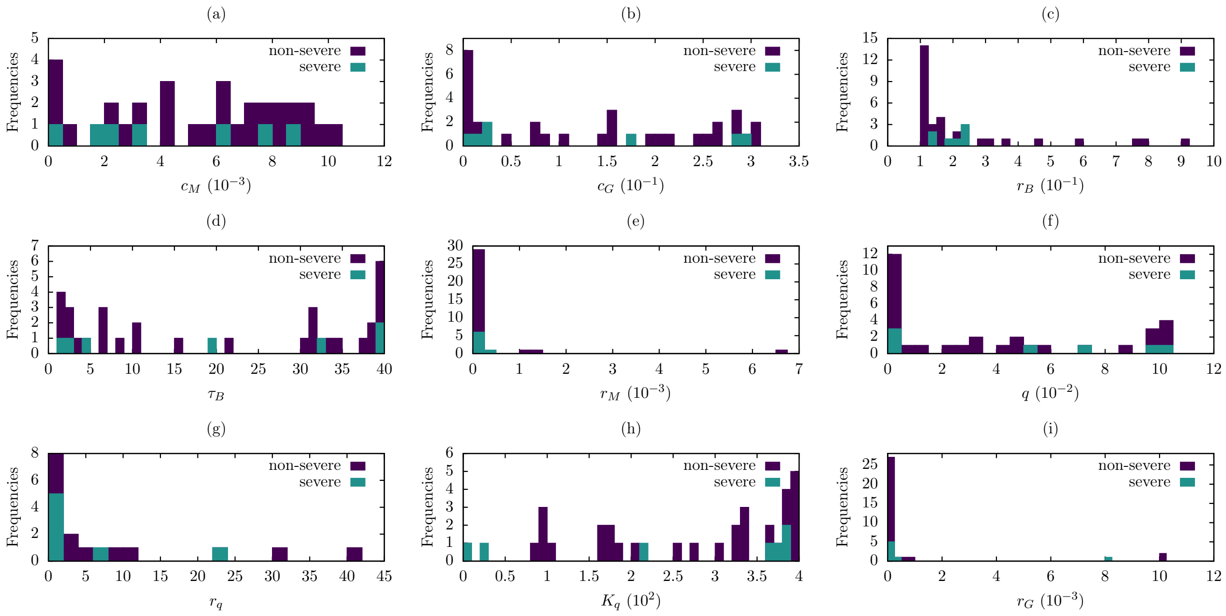


**Figure S1:** Parameter distributions for 7 severe patients (green) and 32 non-severe patients (purple).

#### G. Dynamics of Model 6 for each patient

Dynamics of viral load and immune response for each patient using the Model 6 and the parameter estimated from IgG antibody data of each patient reported in [21]. Severe D patients are presented in yellow color, severe E patients are presented in green color, and non-severe patients are presented in purple color. In each figure: (a) Viral load, (b) T cell level, (c) B cell level, (d) IgM antibody level, (e) IgG antibody level; solid line is the model output and the dots are the data.

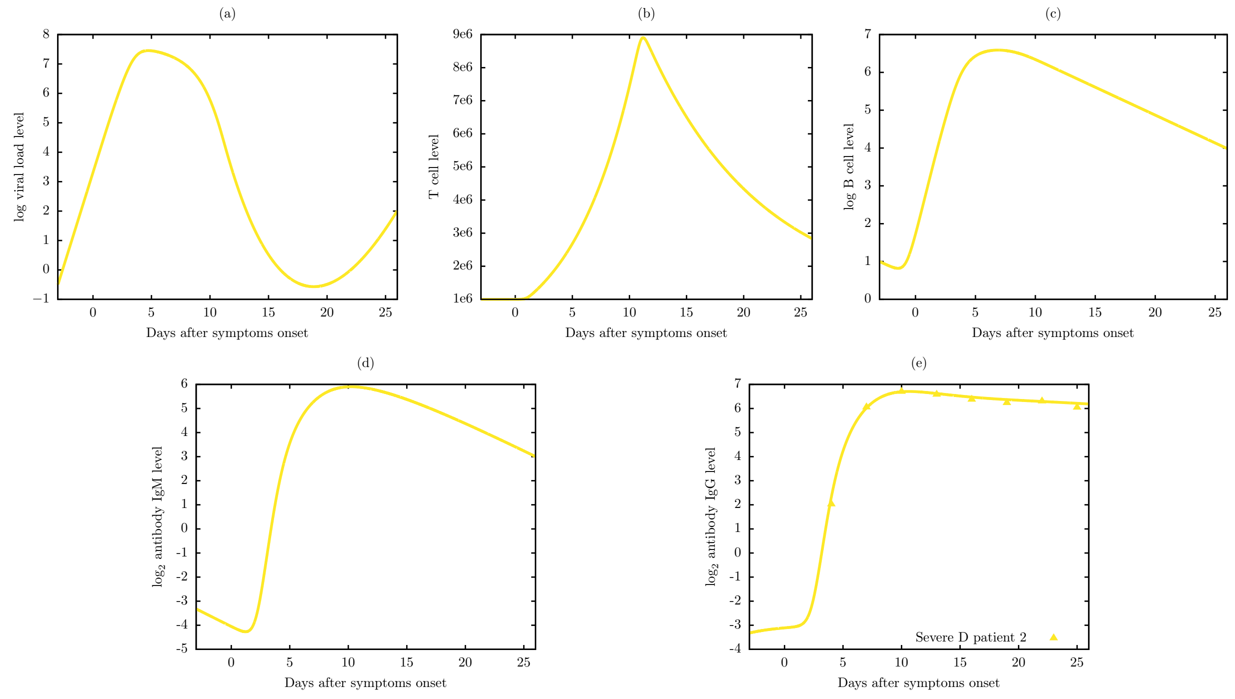


**Figure S2:** Viral dynamic and immune response of Model 6 for severe D patient 2.

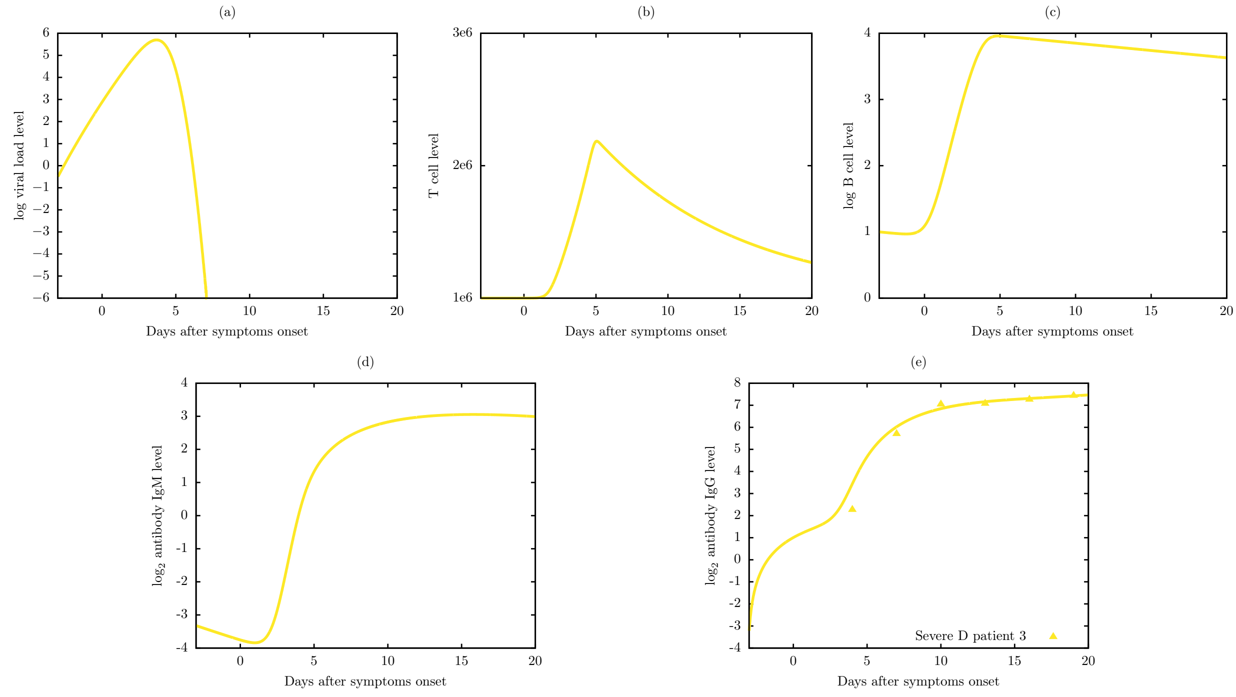


**Figure S3:** Viral dynamic and immune response of Model 6 for severe D patient 3.

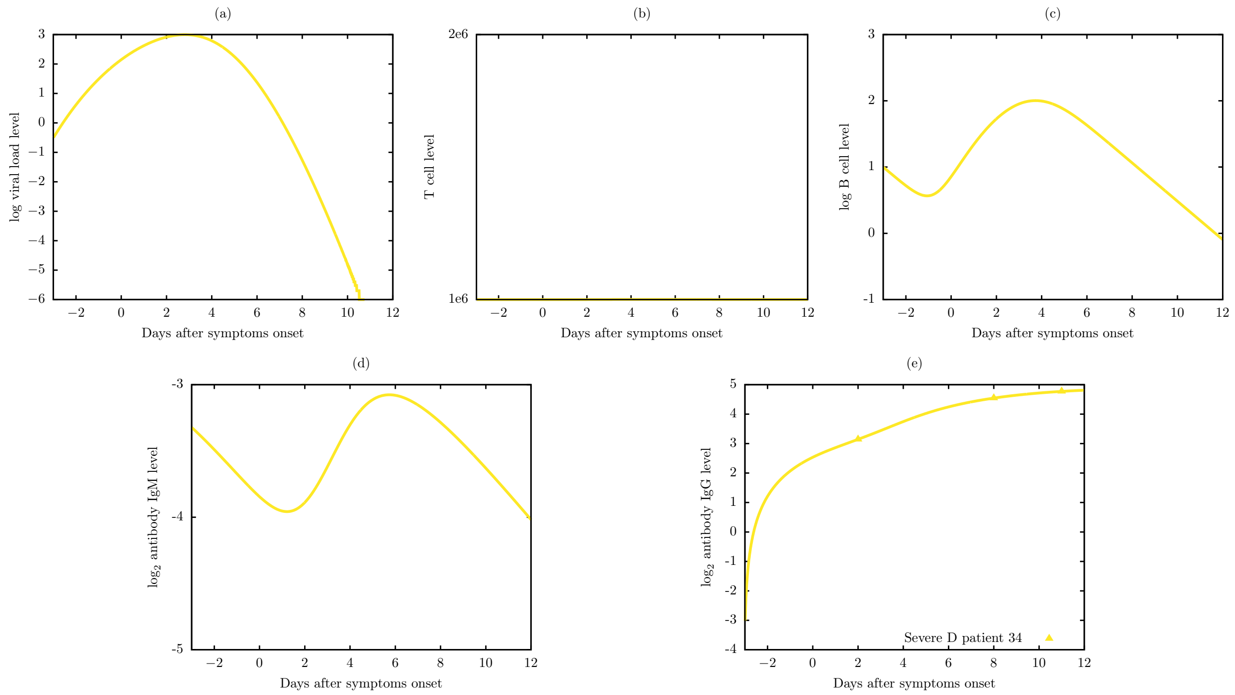


**Figure S4:** Viral dynamic and immune response of Model 6 for severe D patient 34.

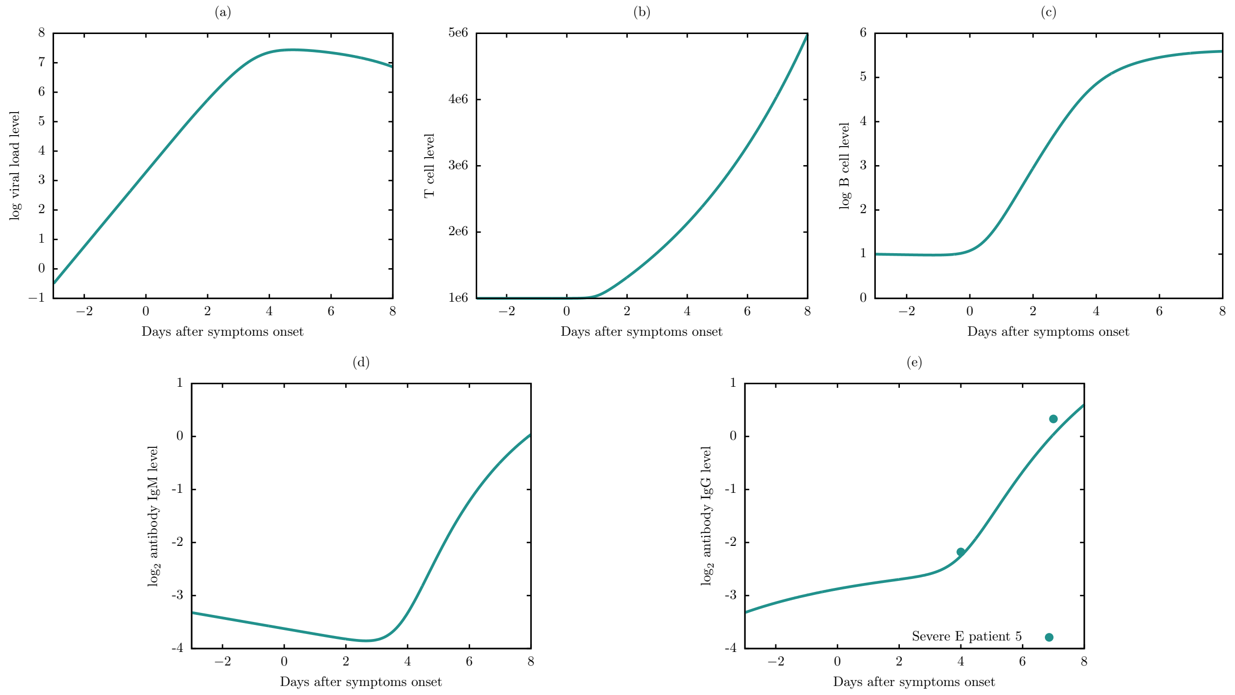


**Figure S5:** Viral dynamic and immune response of Model 6 for severe E patient 5.

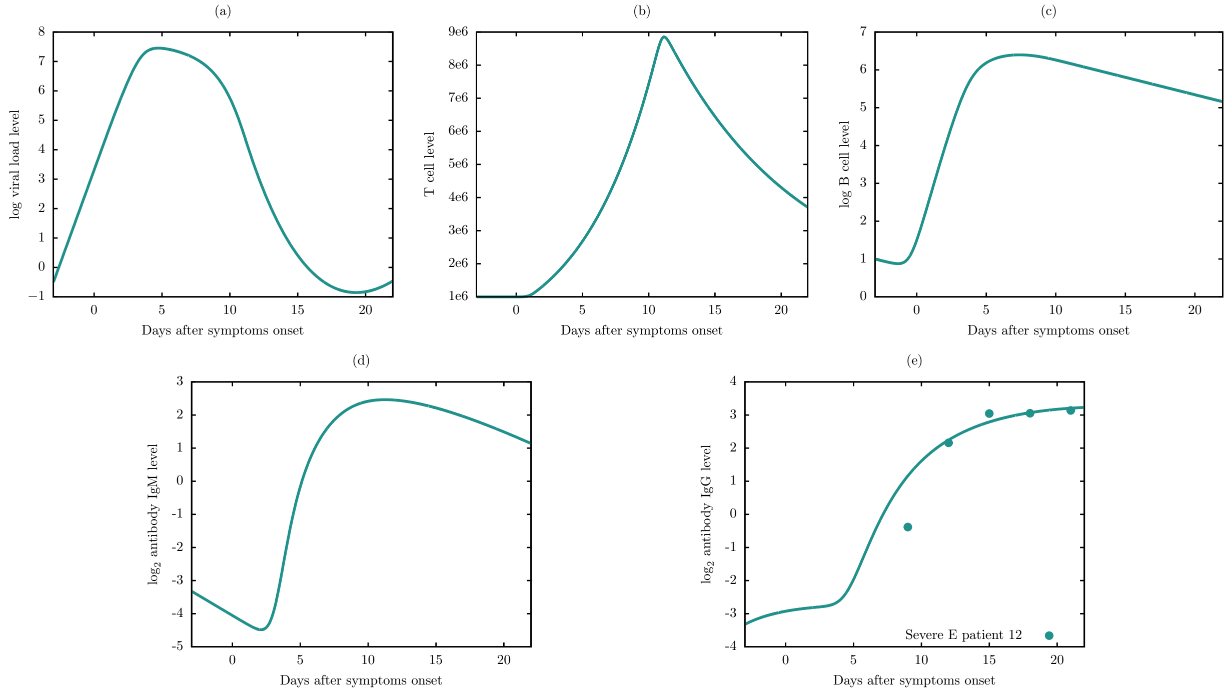


**Figure S6:** Viral dynamic and immune response of Model 6 for severe E patient 12.

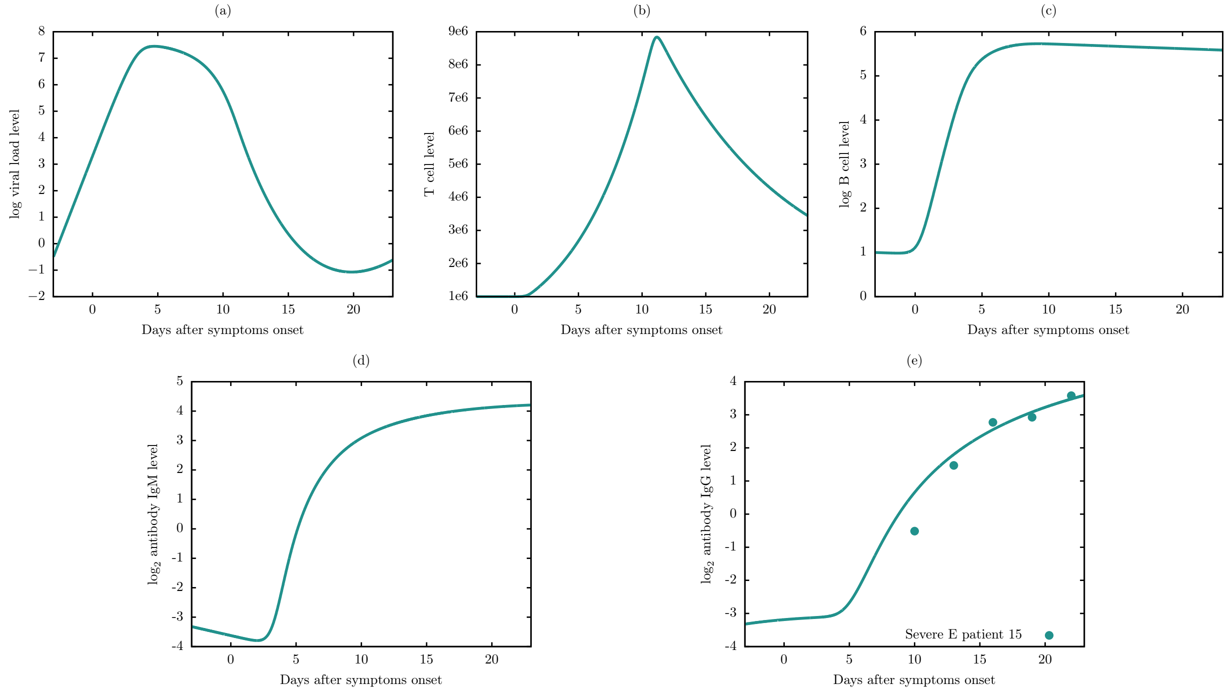


**Figure S7:** Viral dynamic and immune response of Model 6 for severe E patient 15.

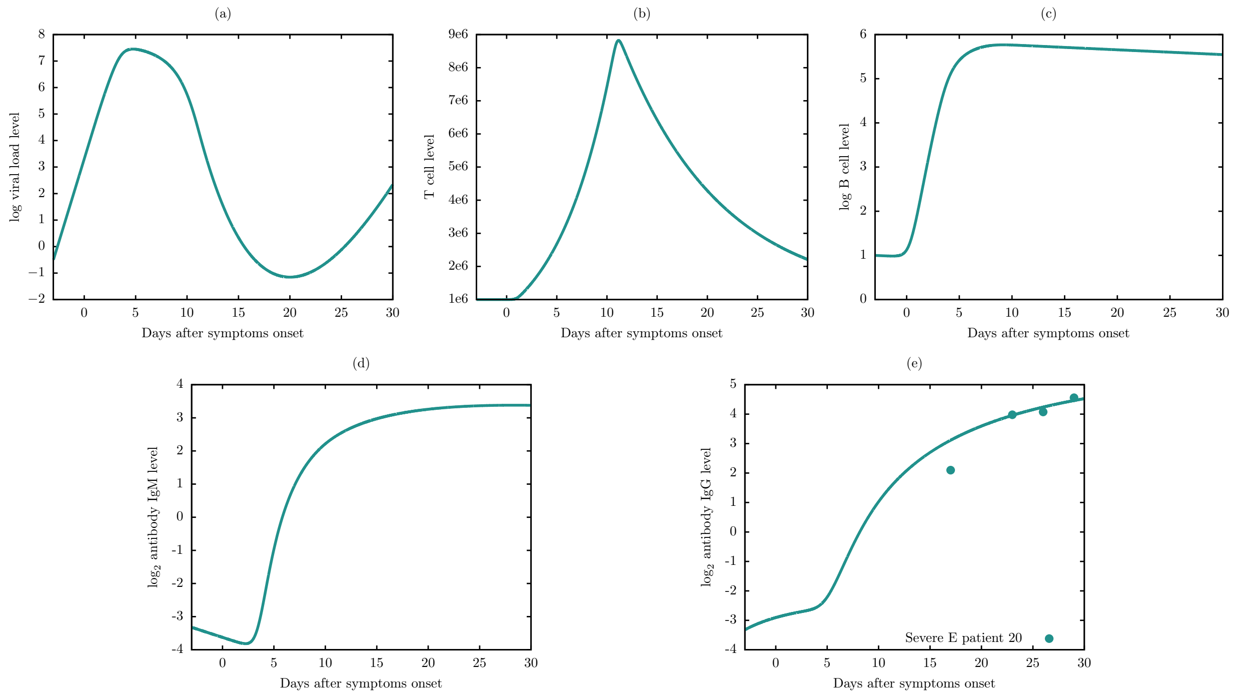


**Figure S8:** Viral dynamic and immune response of Model 6 for severe E patient 20.

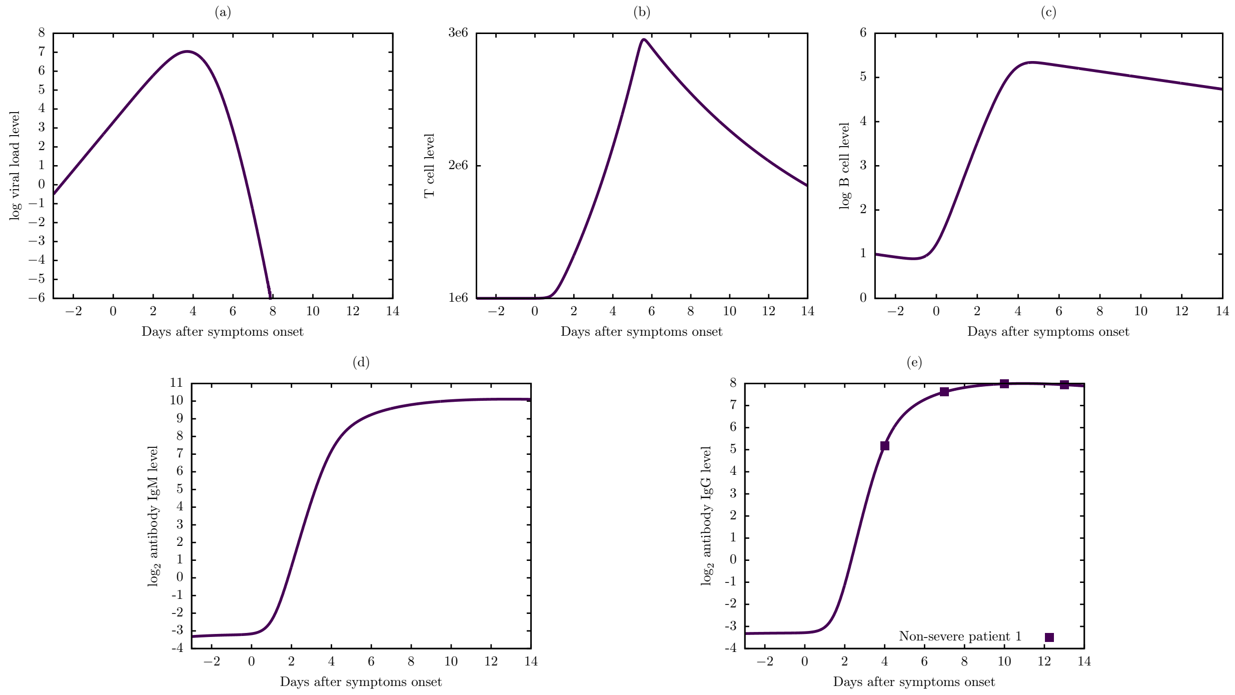


**Figure S9:** Viral dynamic and immune response of Model 6 for non-severe patient 1.

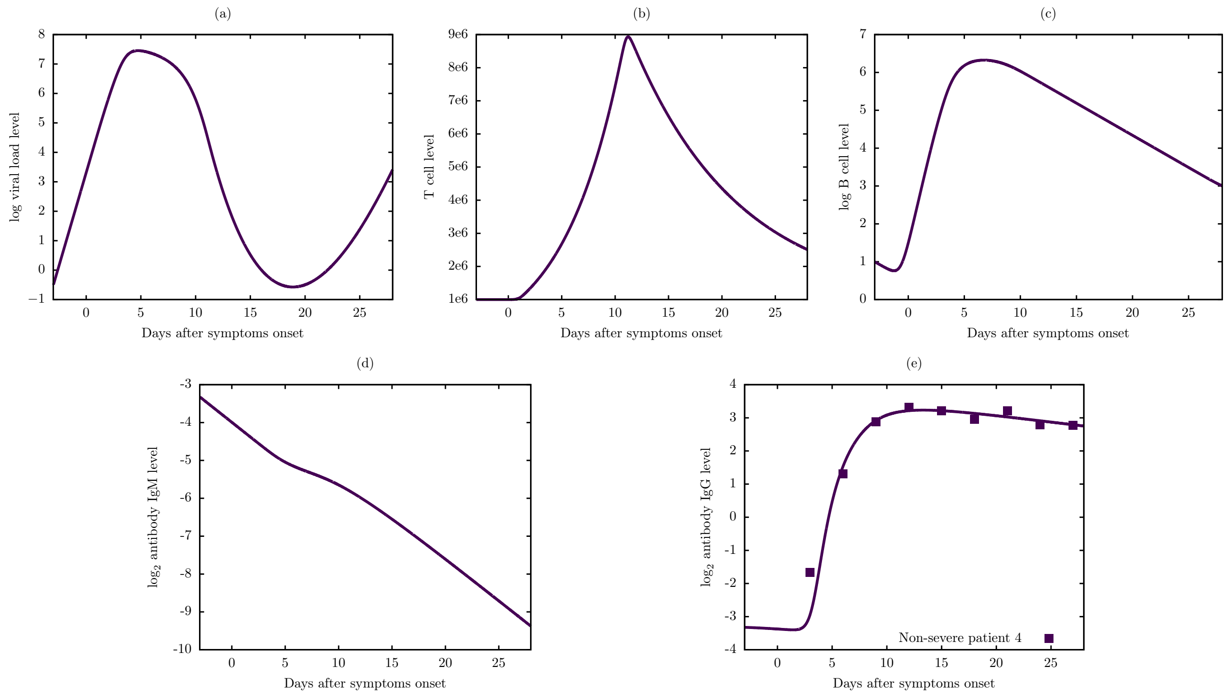


**Figure S10:** Viral dynamic and immune response of Model 6 for non-severe patient 4.

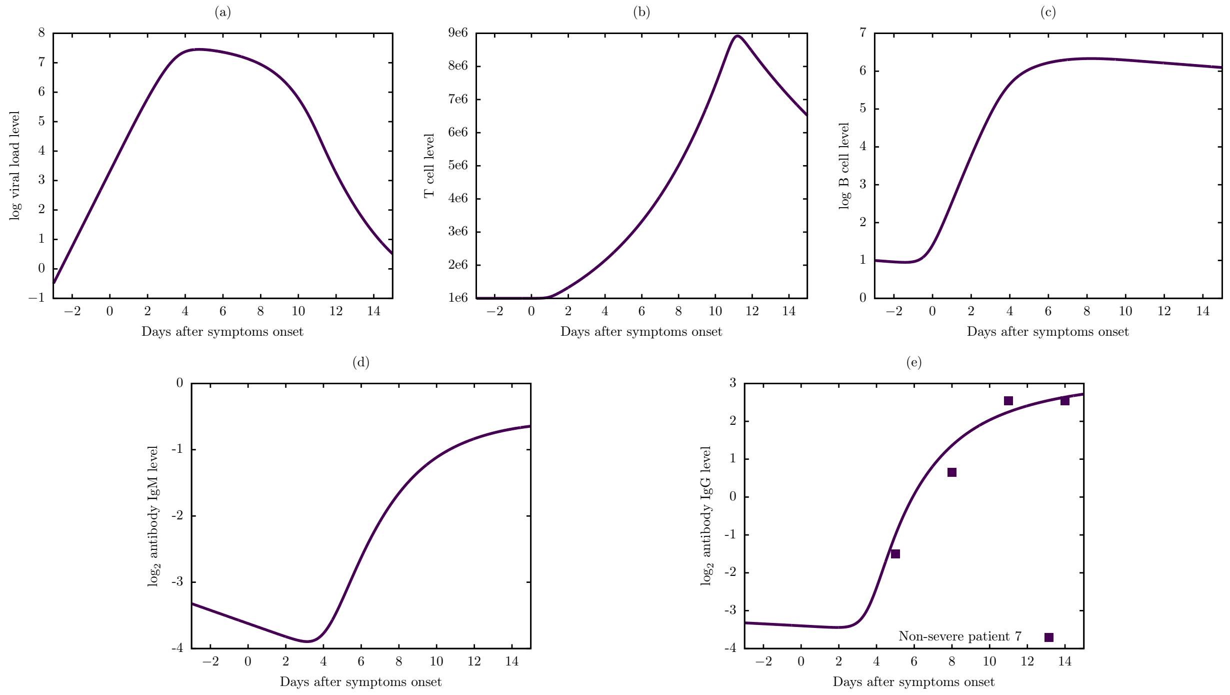


**Figure S11:** Viral dynamic and immune response of Model 6 for non-severe patient 7.

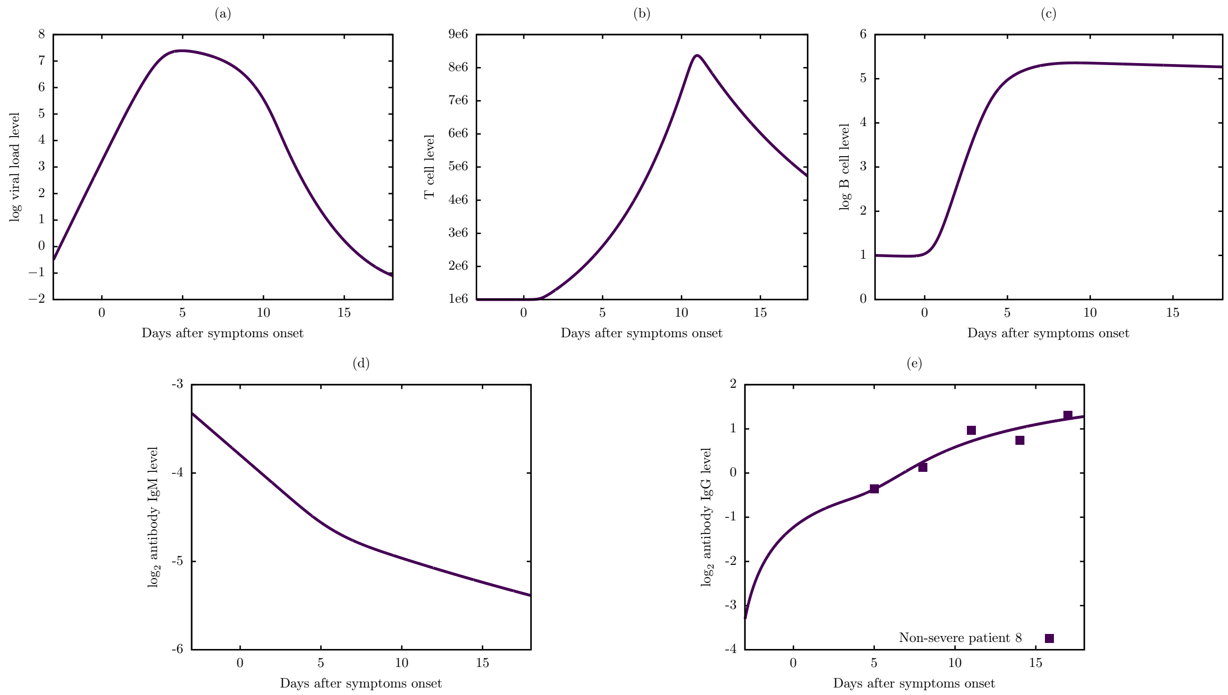


**Figure S12:** Viral dynamic and immune response of Model 6 for non-severe patient 8.

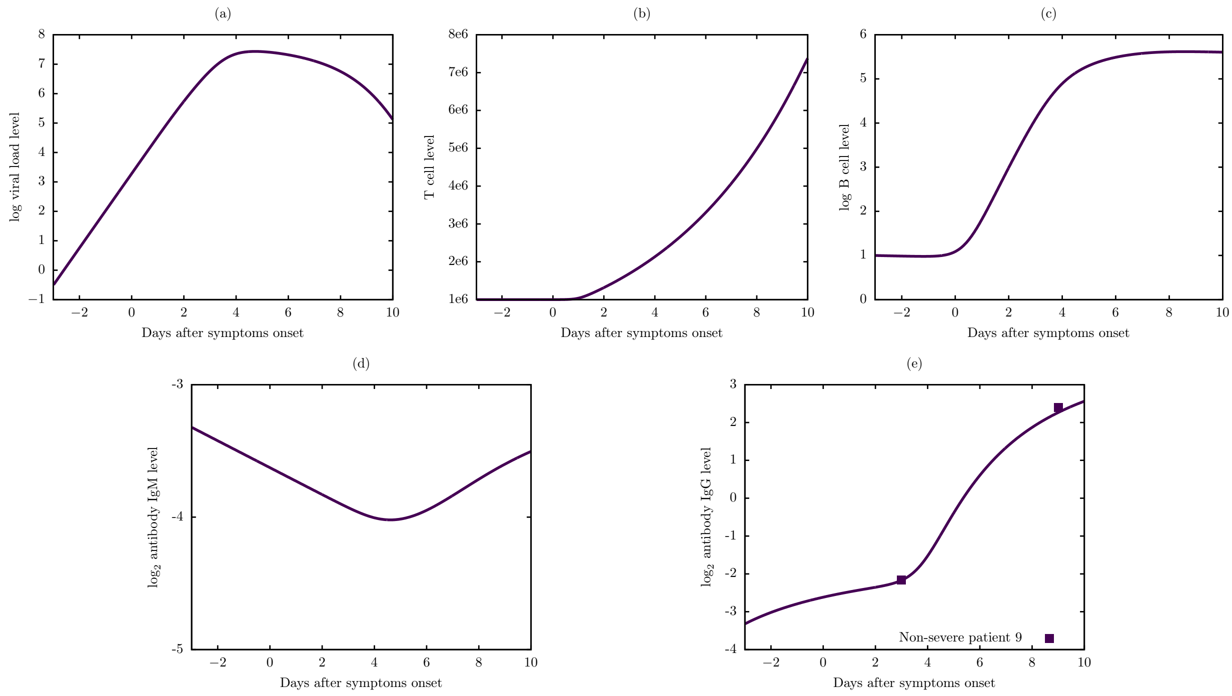


**Figure S13:** Viral dynamic and immune response of Model 6 for non-severe patient 9.

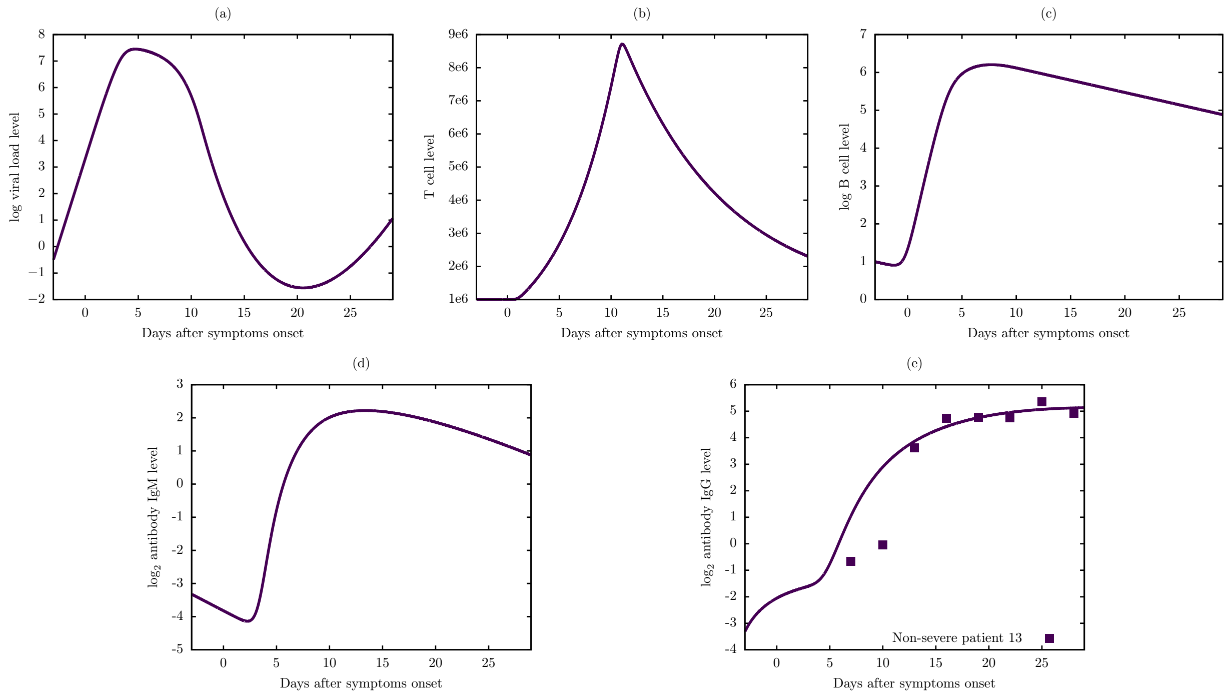


**Figure S14:** Viral dynamic and immune response of Model 6 for non-severe patient 13.

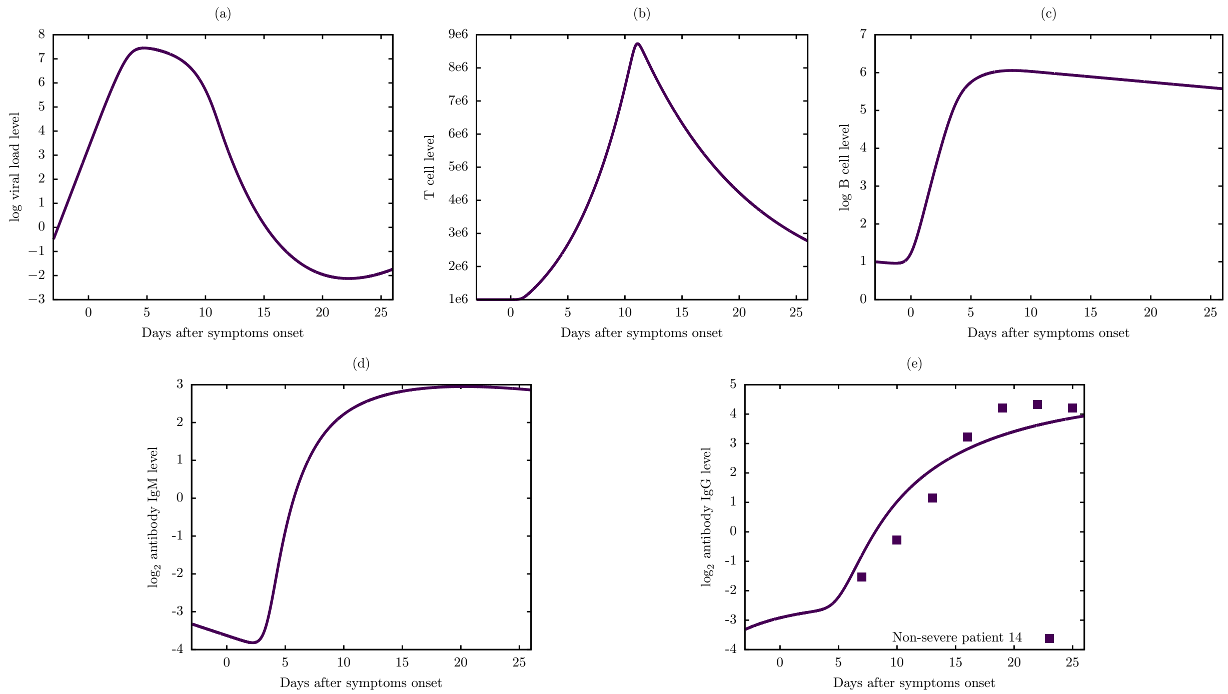


**Figure S15:** Viral dynamic and immune response of Model 6 for non-severe patient 14.

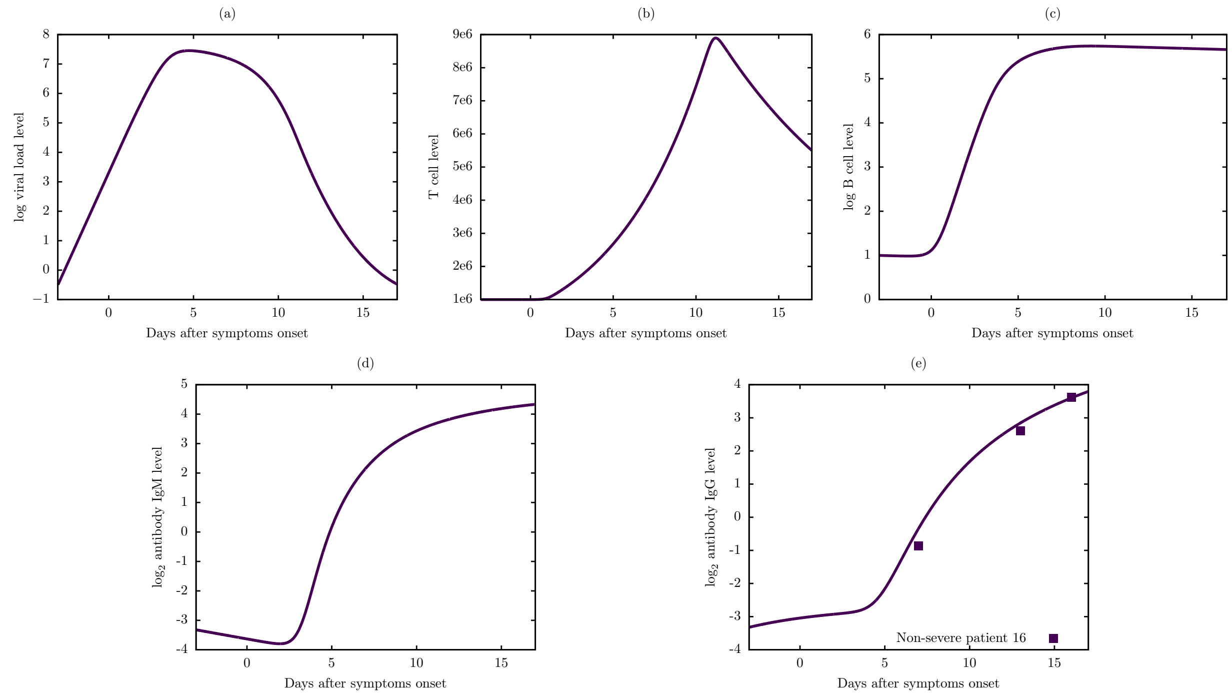


**Figure S16:** Viral dynamic and immune response of Model 6 for non-severe patient 16.

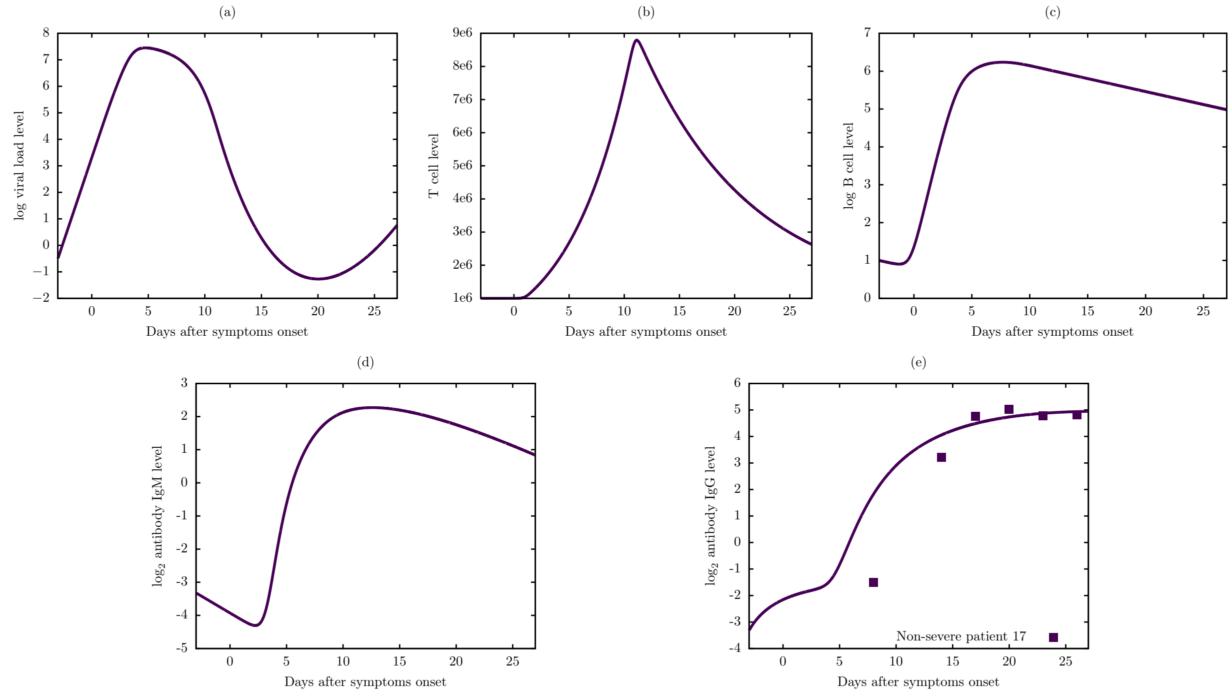


**Figure S17:** Viral dynamic and immune response of Model 6 for non-severe patient 17.

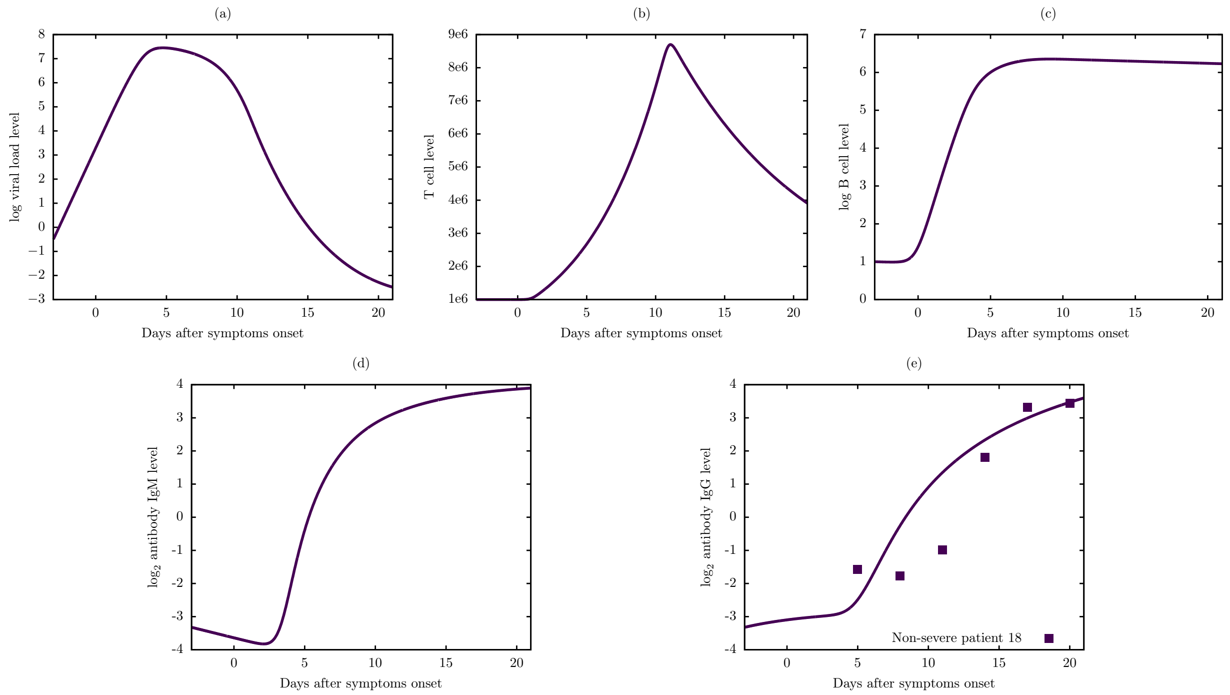


**Figure S18:** Viral dynamic and immune response of Model 6 for non-severe patient 18.

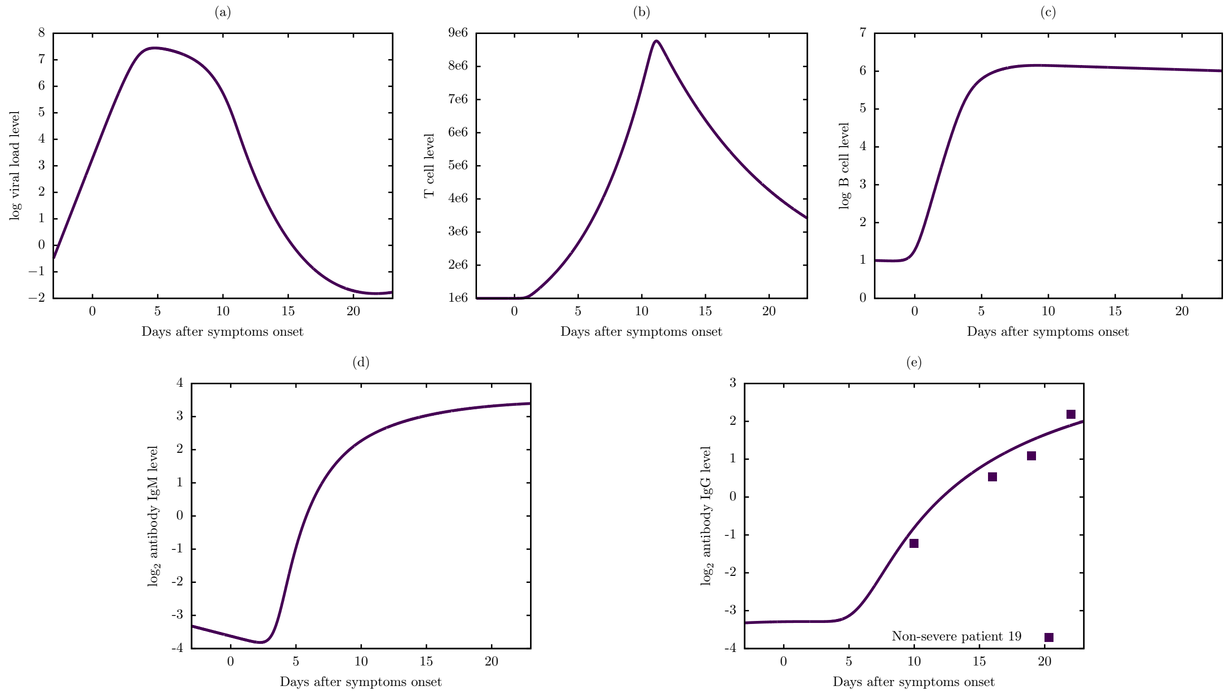


**Figure S19:** Viral dynamic and immune response of Model 6 for non-severe patient 19.

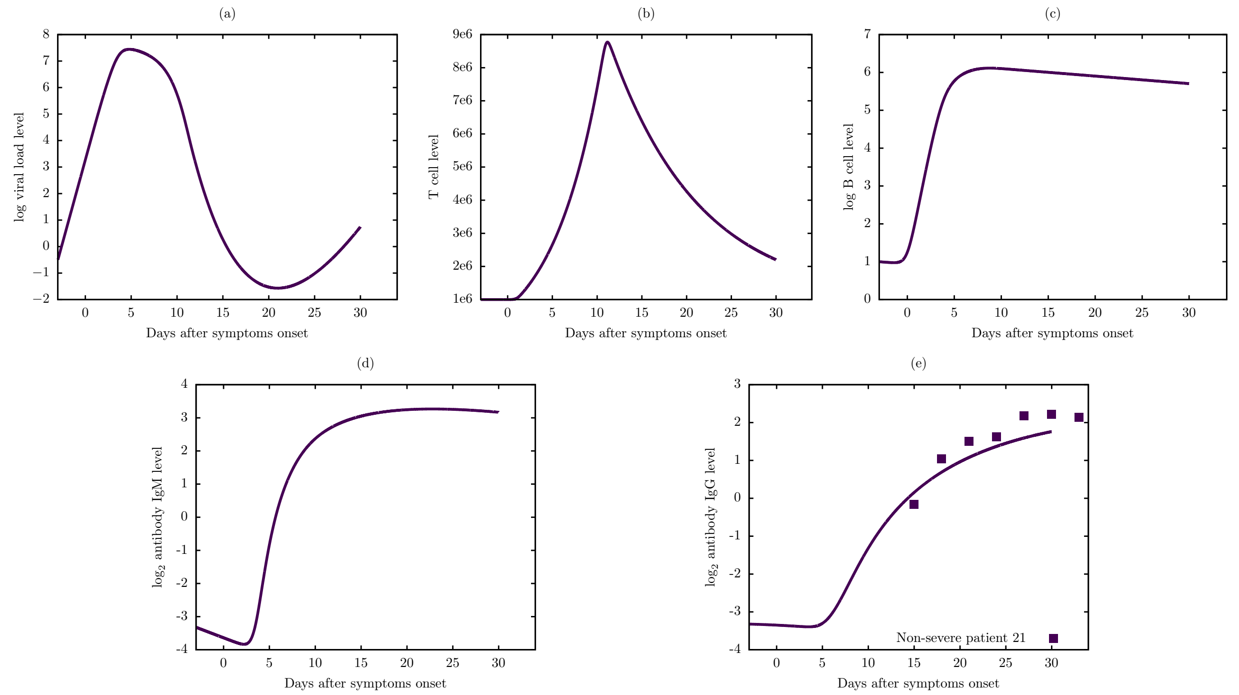


**Figure S20:** Viral dynamic and immune response of Model 6 for non-severe patient 21.

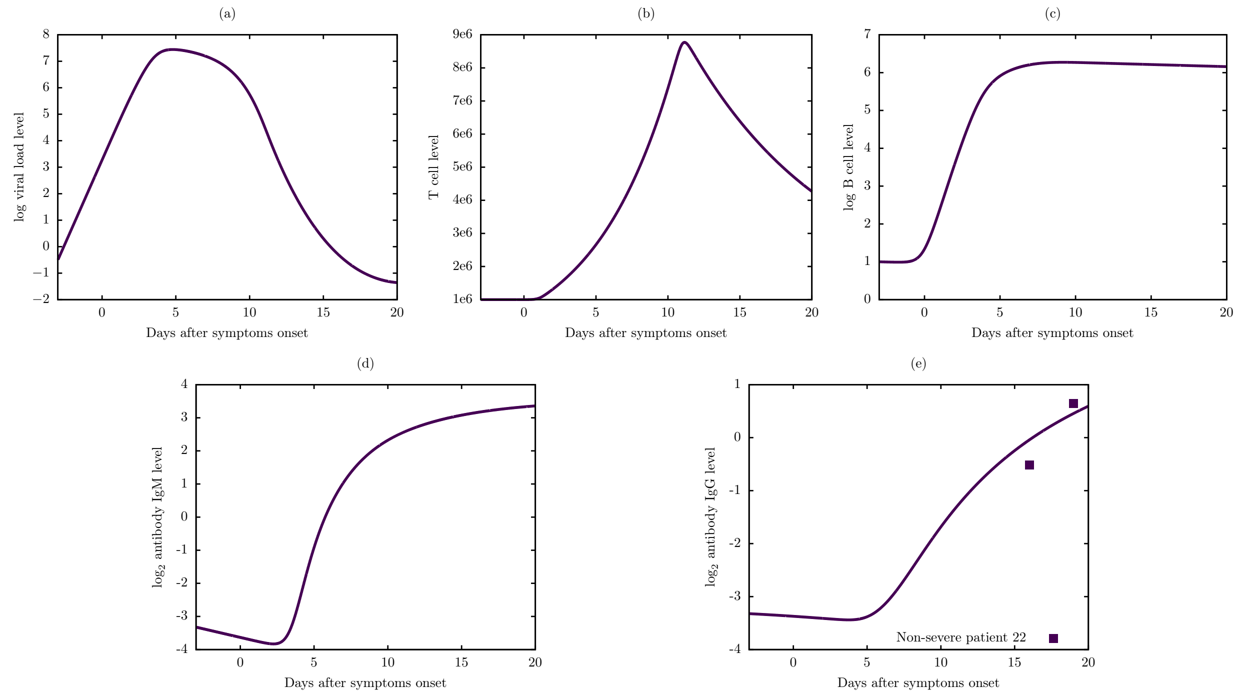


**Figure S21:** Viral dynamic and immune response of Model 6 for non-severe patient 22.

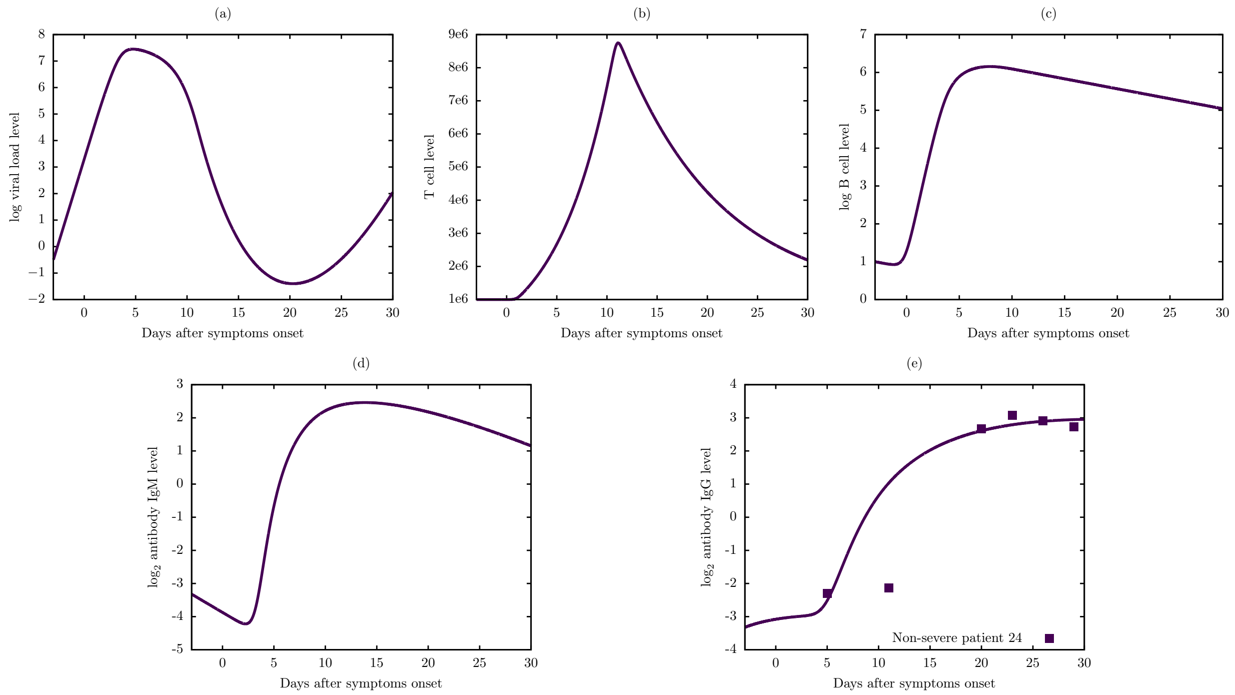


**Figure S22:** Viral dynamic and immune response of Model 6 for non-severe patient 24.

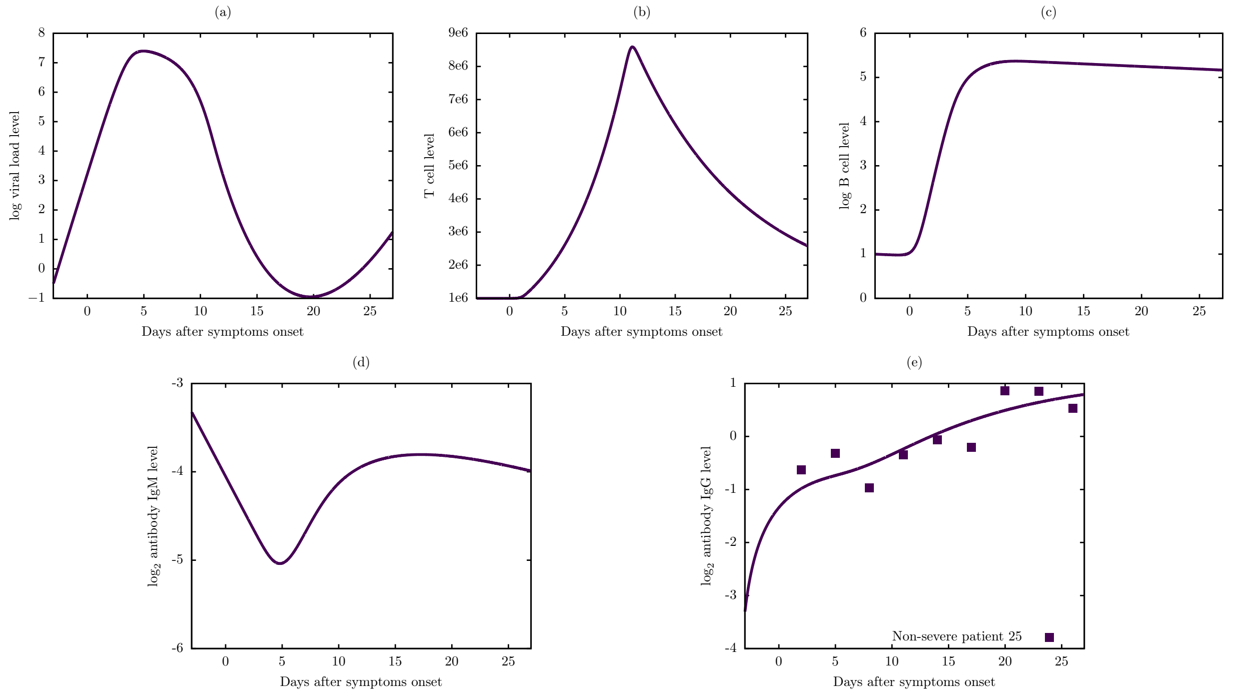


**Figure S23:** Viral dynamic and immune response of Model 6 for non-severe patient 25.

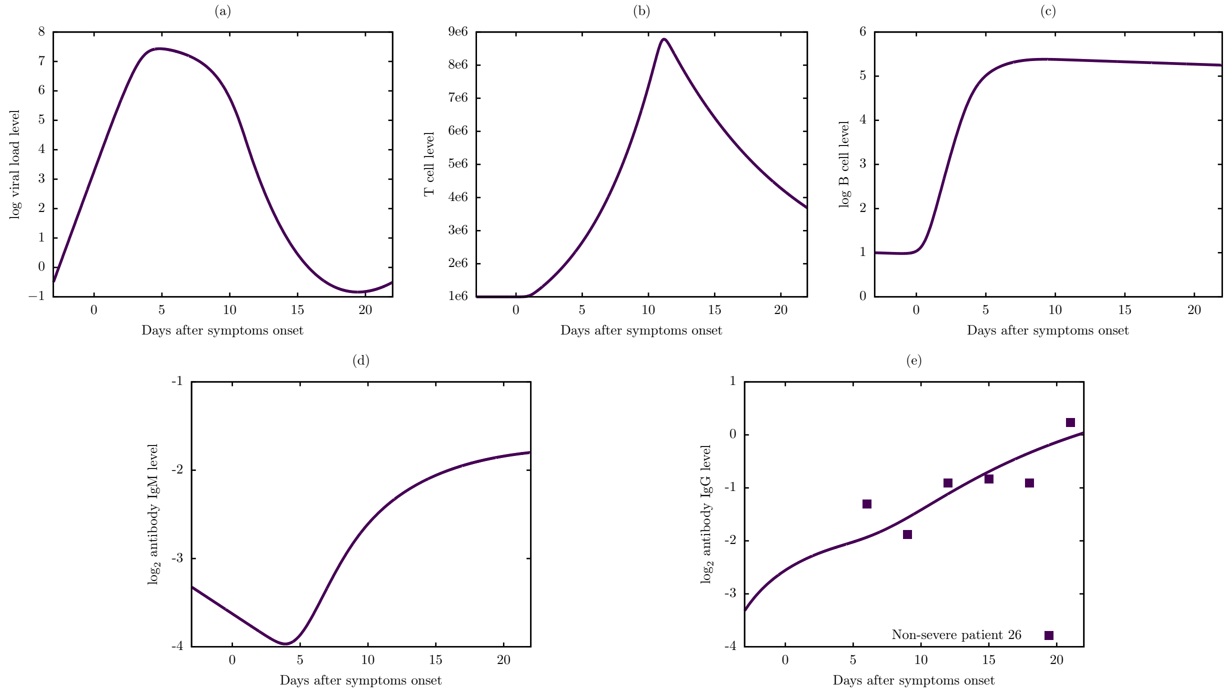


**Figure S24:** Viral dynamic and immune response of Model 6 for non-severe patient 26.

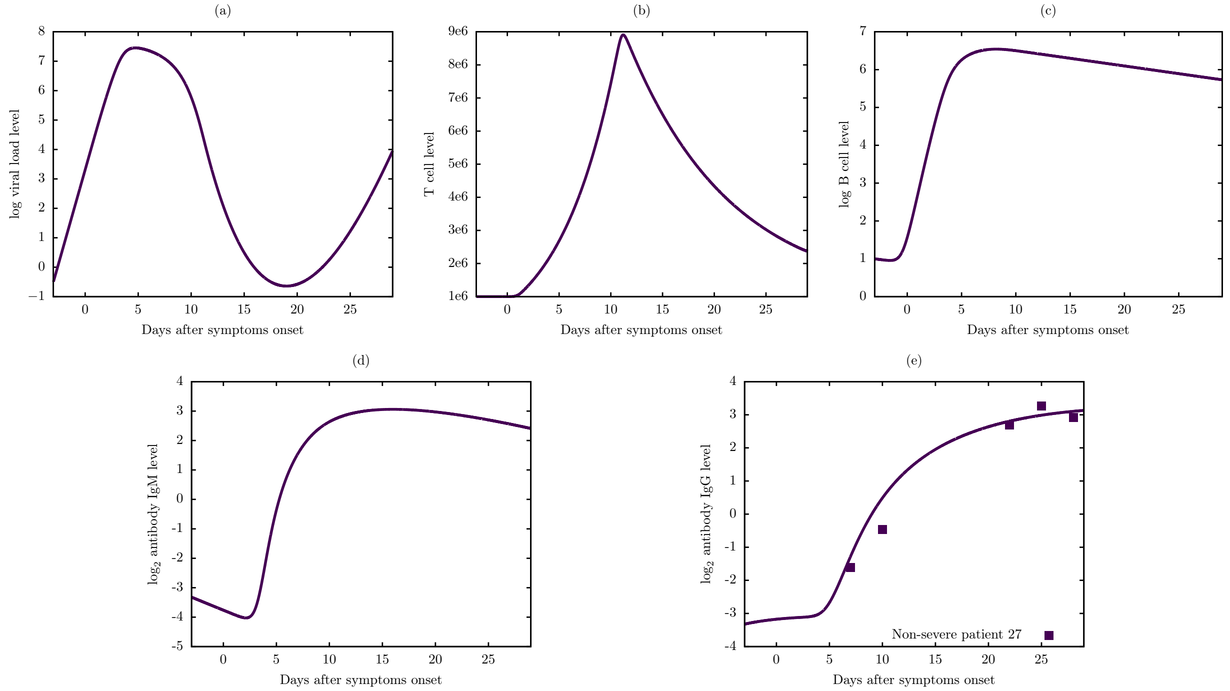


**Figure S25:** Viral dynamic and immune response of Model 6 for non-severe patient 27.

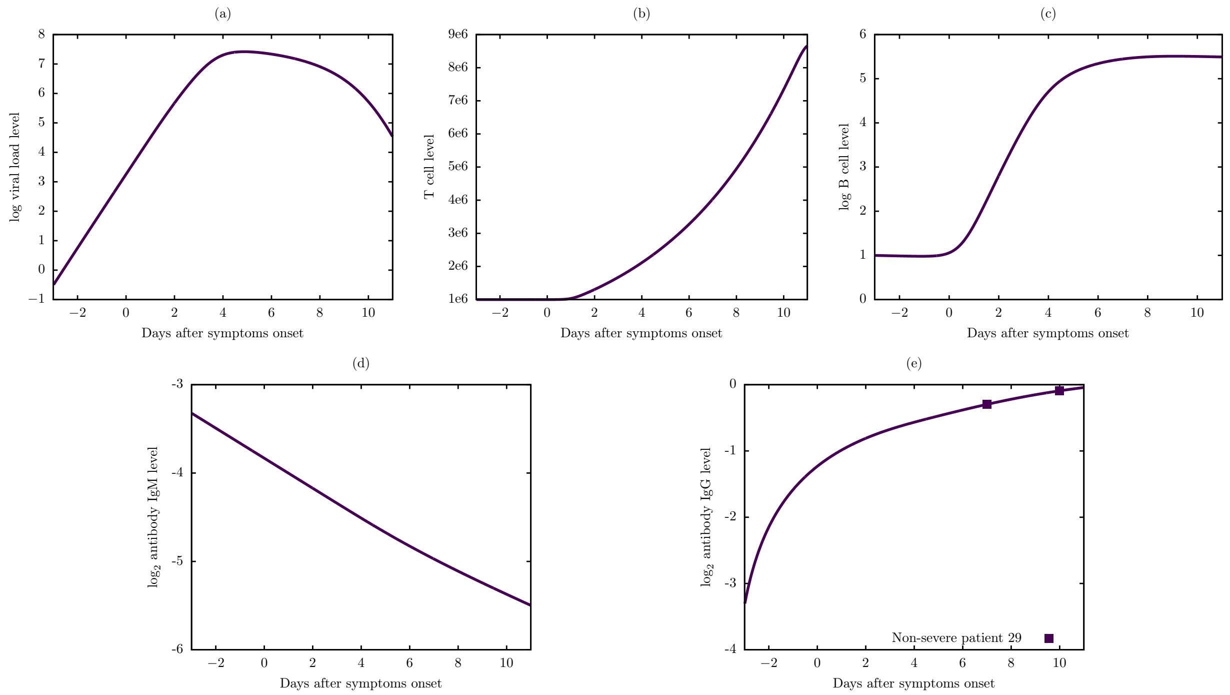


**Figure S26:** Viral dynamic and immune response of Model 6 for non-severe patient 29.

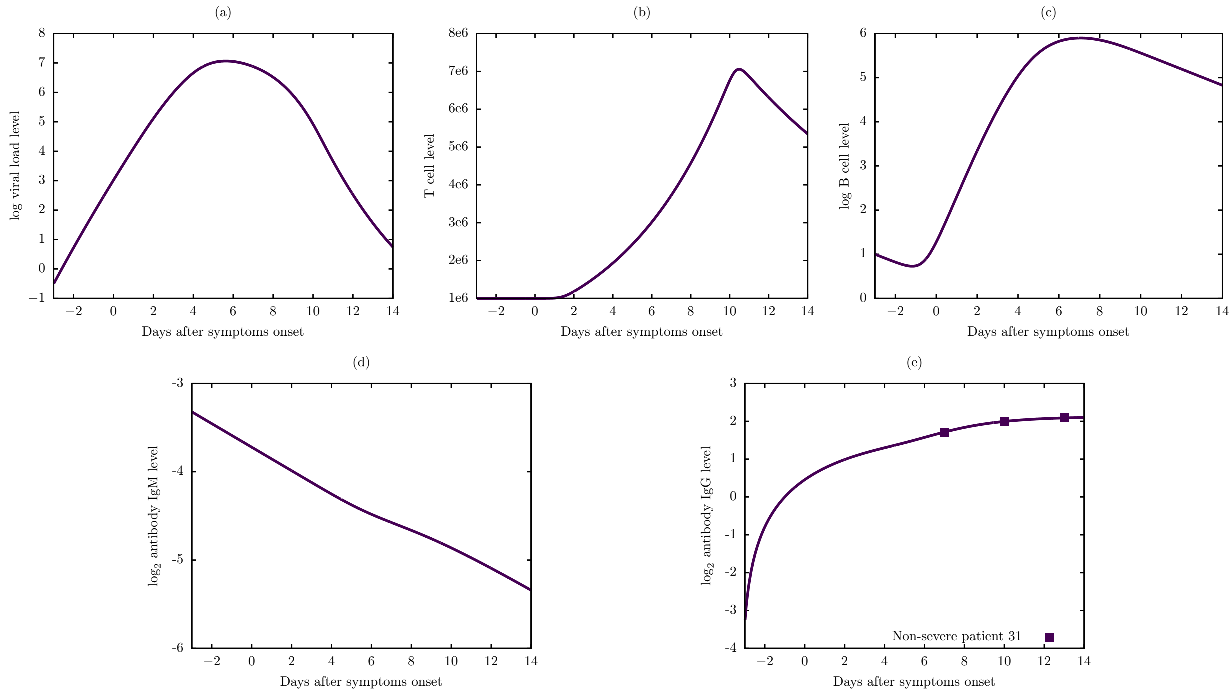


**Figure S27:** Viral dynamic and immune response of Model 6 for non-severe patient 31.

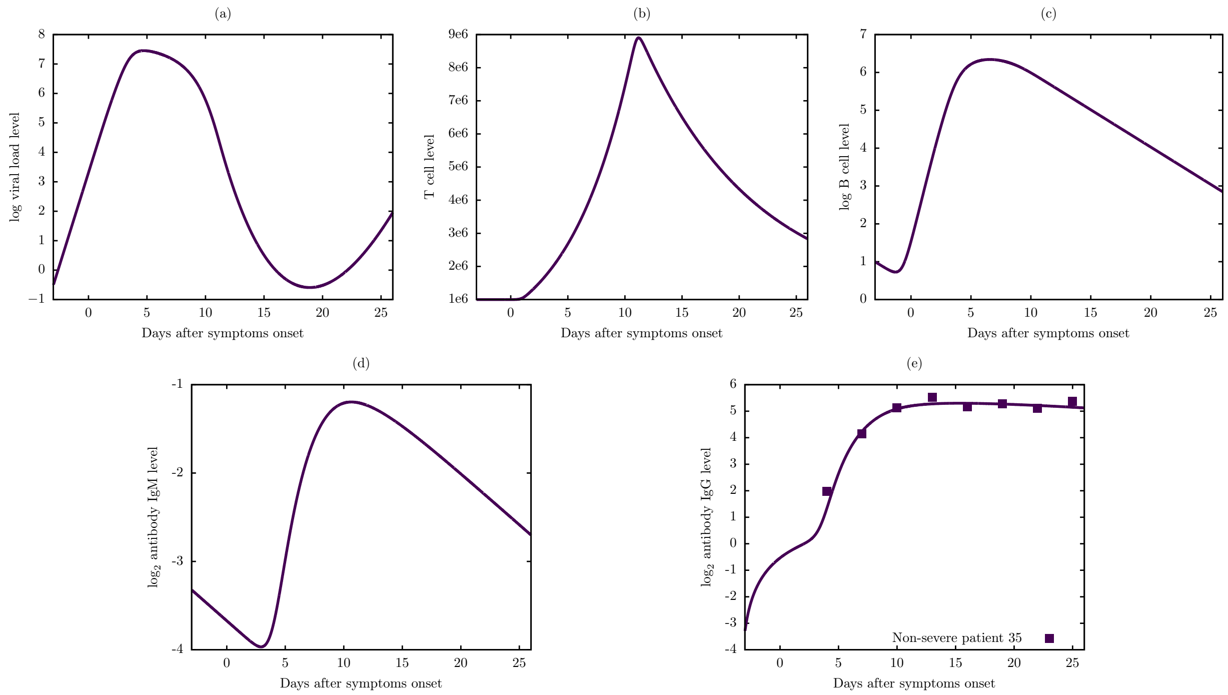


**Figure S28:** Viral dynamic and immune response of Model 6 for non-severe patient 35.

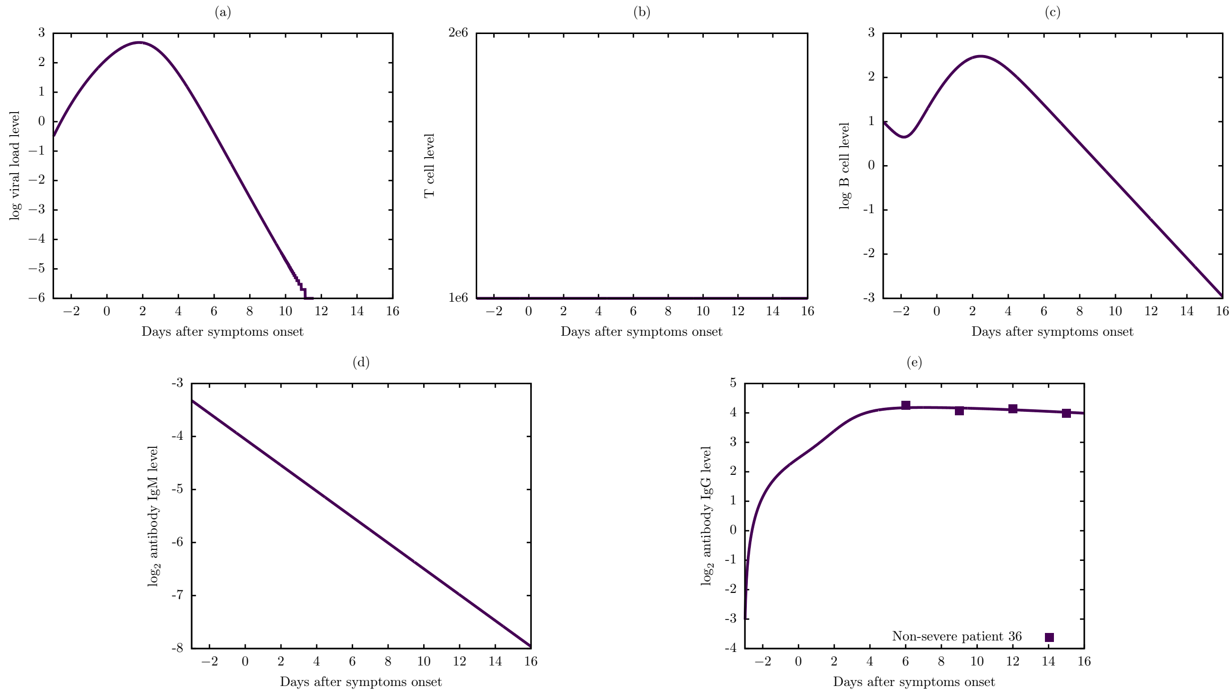


**Figure S29:** Viral dynamic and immune response of Model 6 for non-severe patient 36.

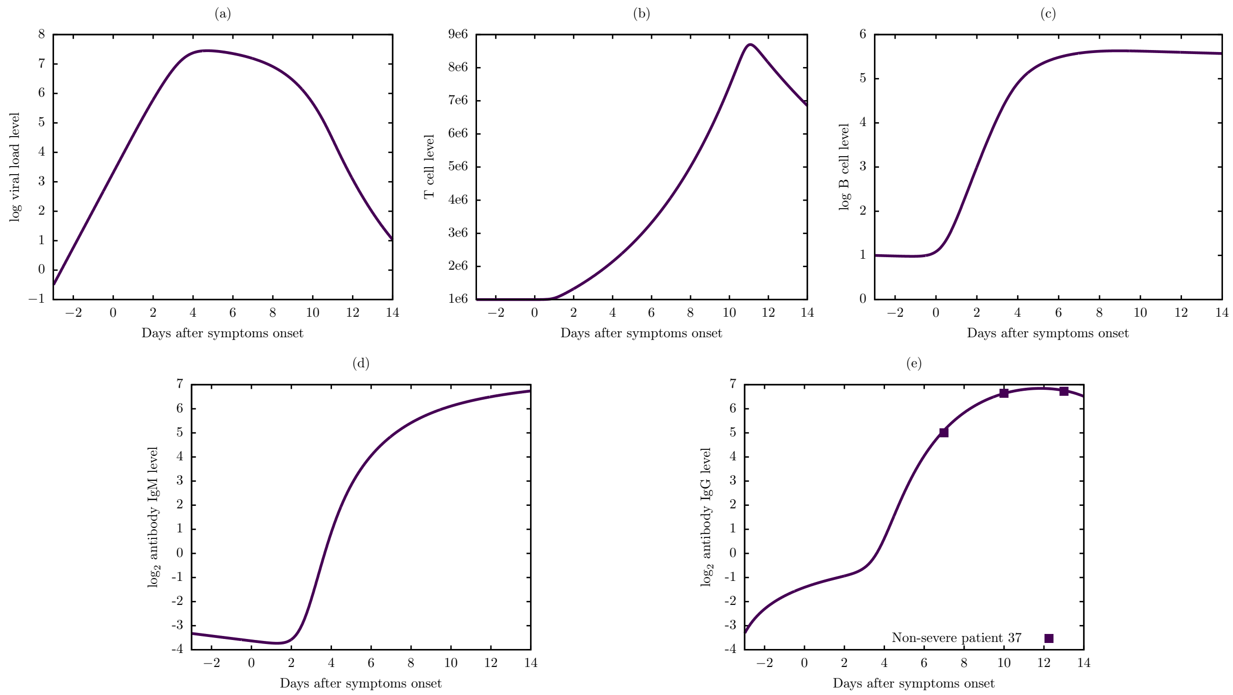


**Figure S30:** Viral dynamic and immune response of Model 6 for non-severe patient 37.

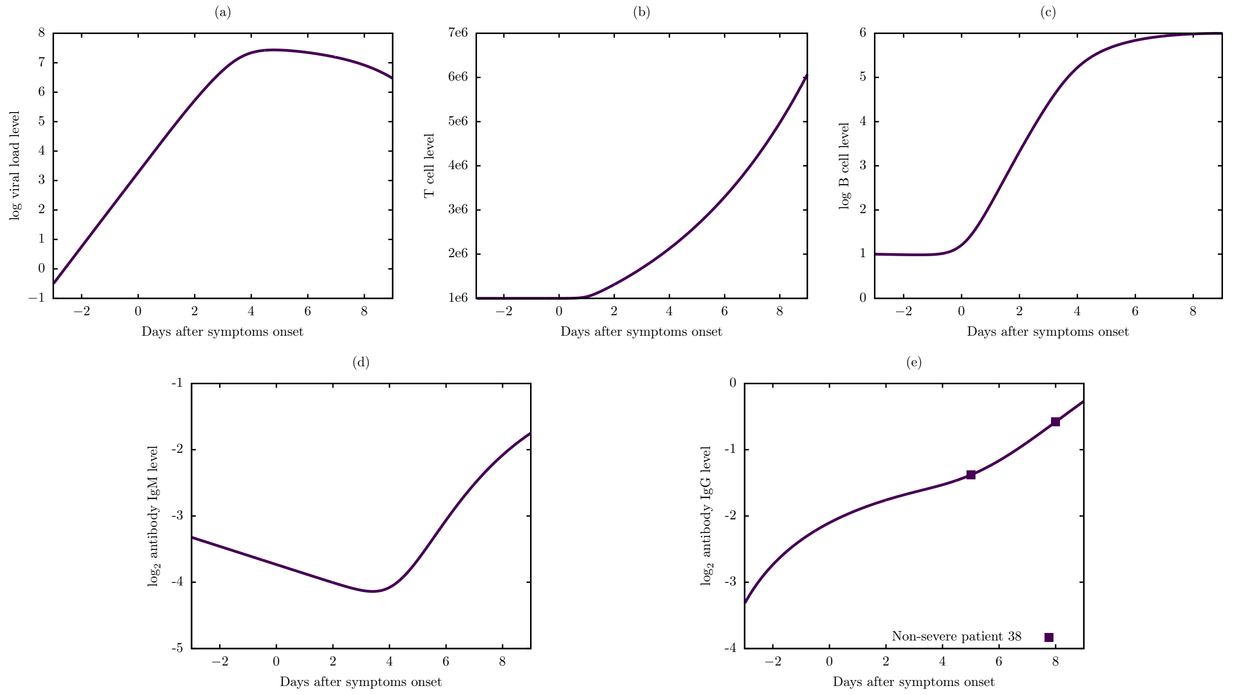


**Figure S31:** Viral dynamic and immune response of Model 6 for non-severe patient 38.

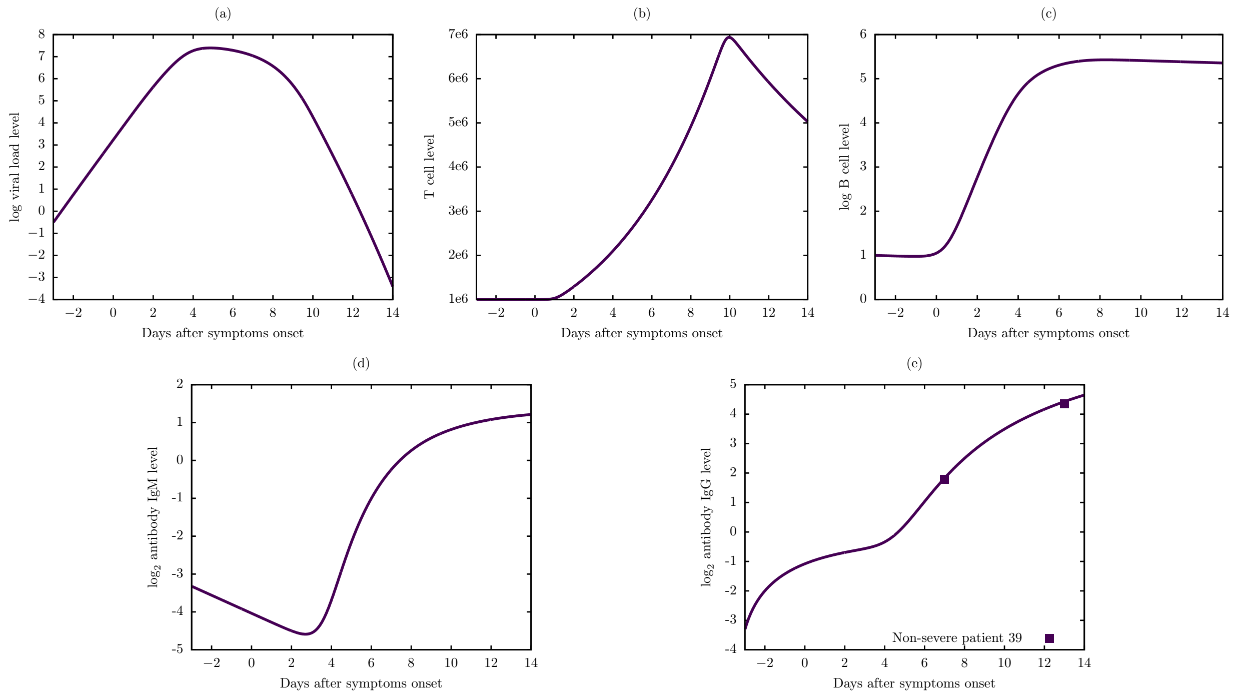


**Figure S32:** Viral dynamic and immune response of Model 6 for non-severe patient 39.

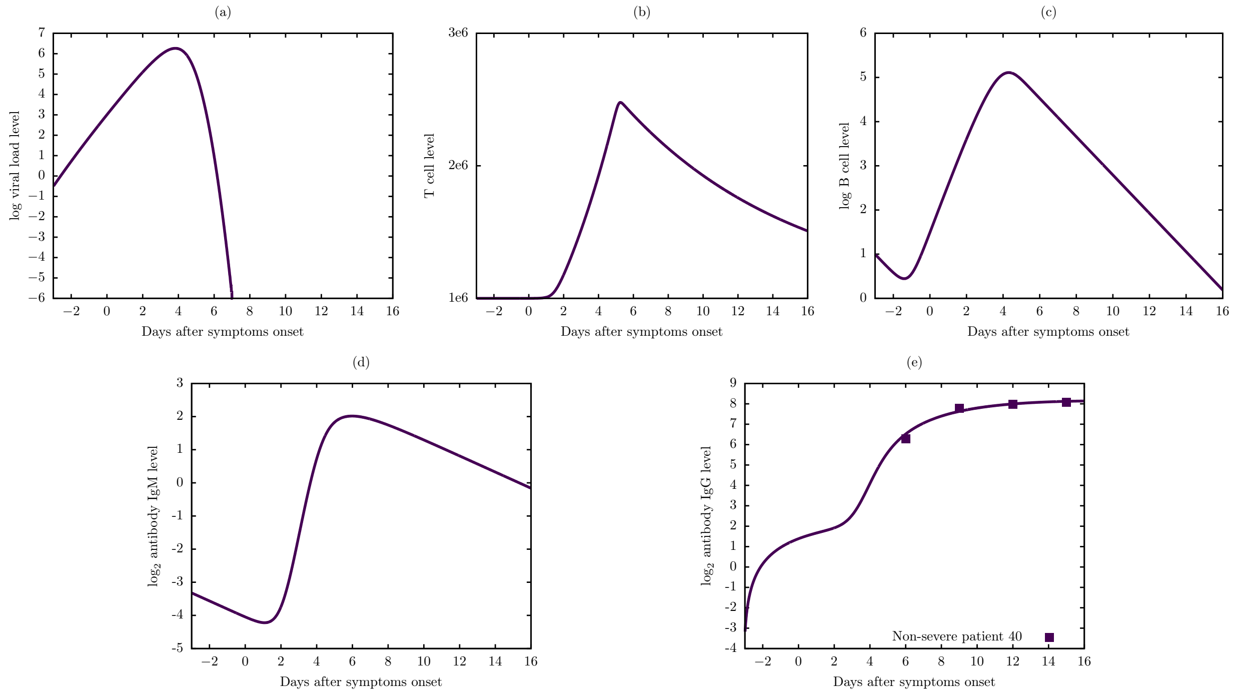


**Figure S33:** Viral dynamic and immune response of Model 6 for non-severe patient 40.

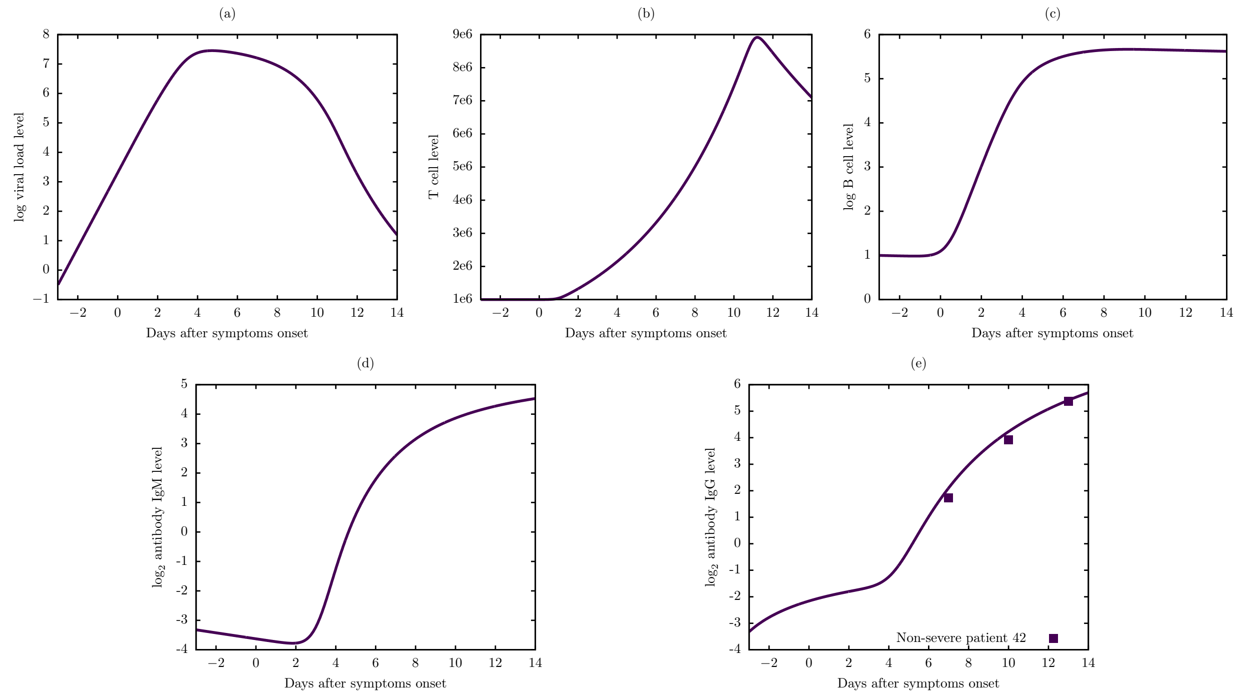


**Figure S34:** Viral dynamic and immune response of Model 6 for non-severe patient 42.

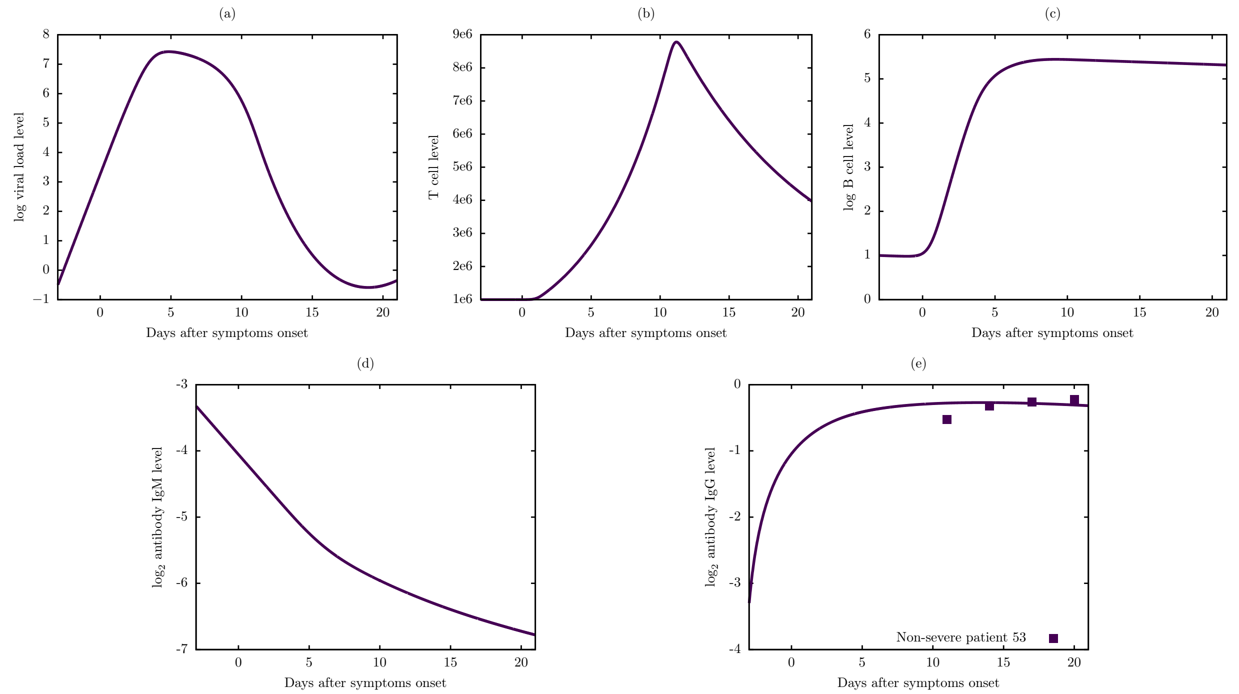


**Figure S35:** Viral dynamic and immune response of Model 6 for non-severe patient 53.

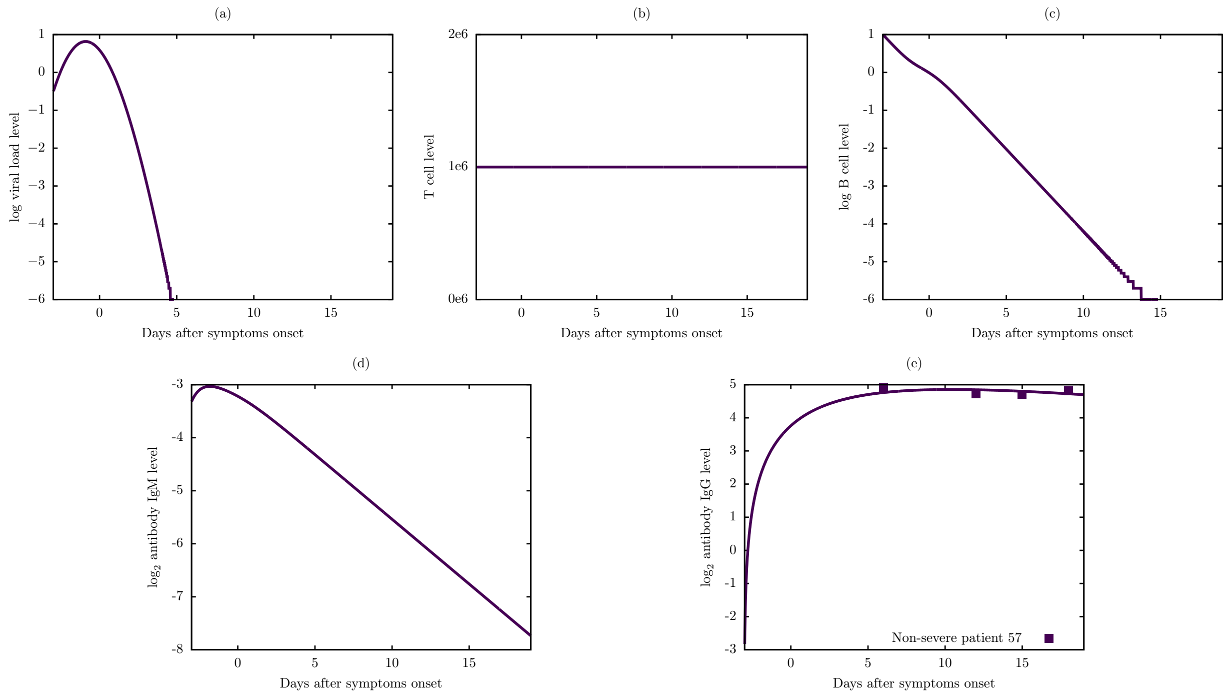


**Figure S36:** Viral dynamic and immune response of Model 6 for non-severe patient 57.

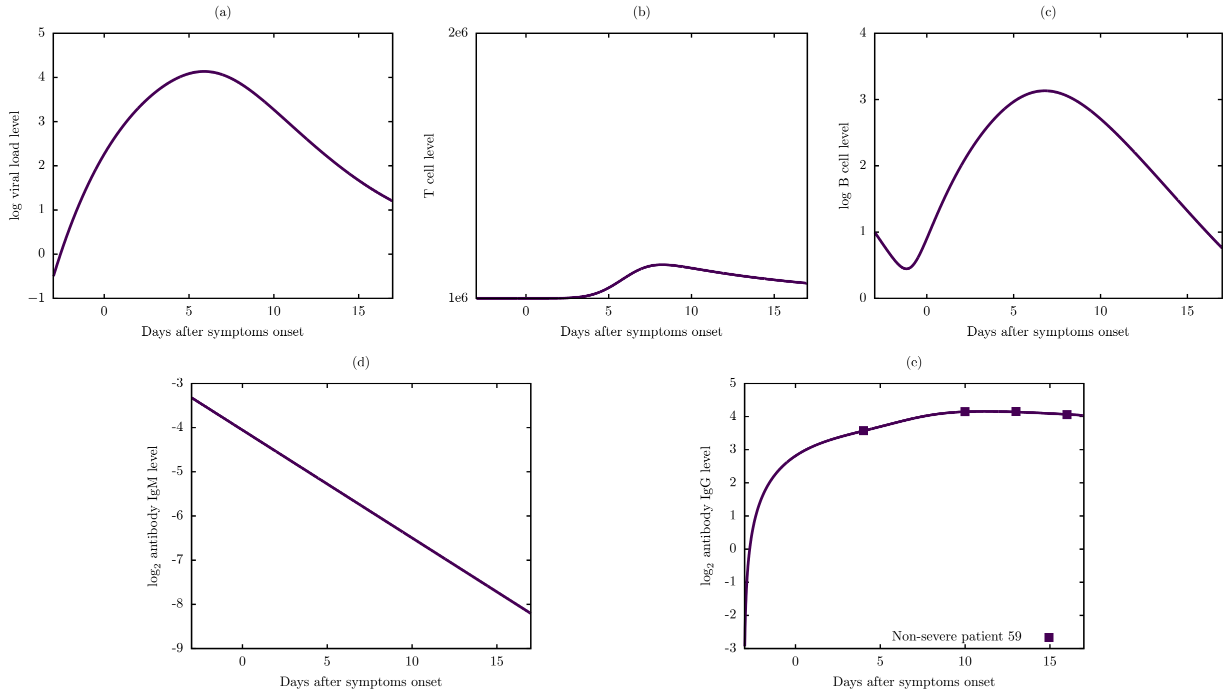


**Figure S37:** Viral dynamic and immune response of Model 6 for non-severe patient 59.

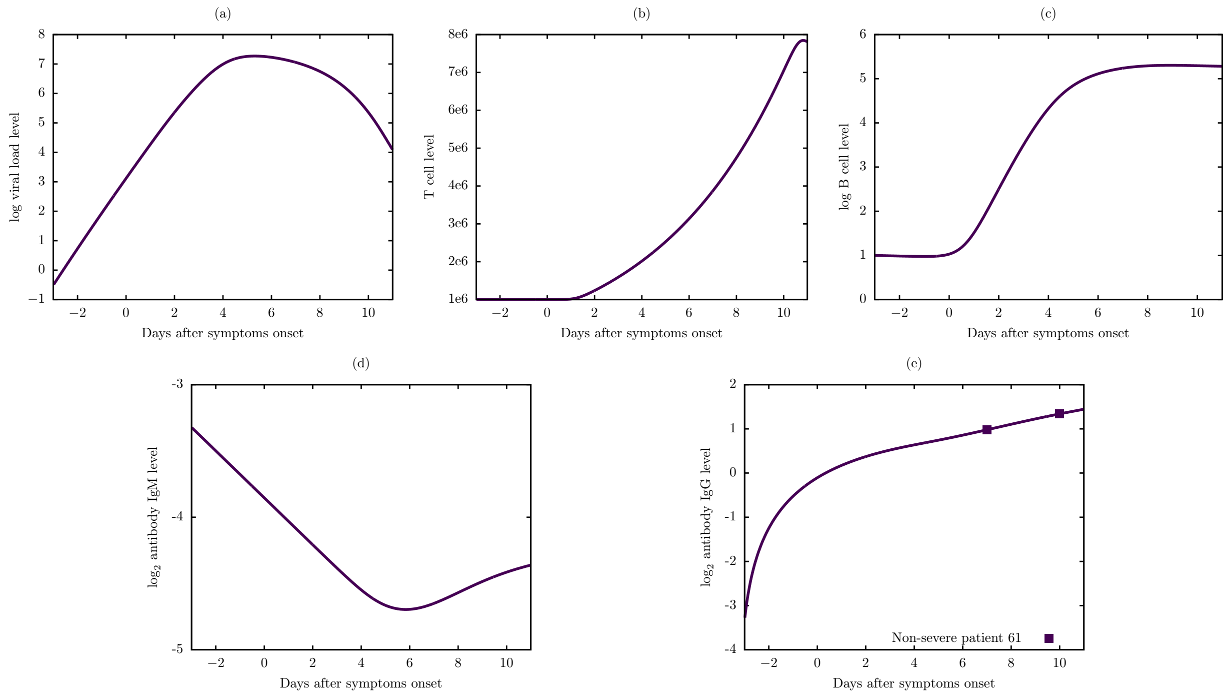


**Figure S38:** Viral dynamic and immune response of Model 6 for non-severe patient 61.

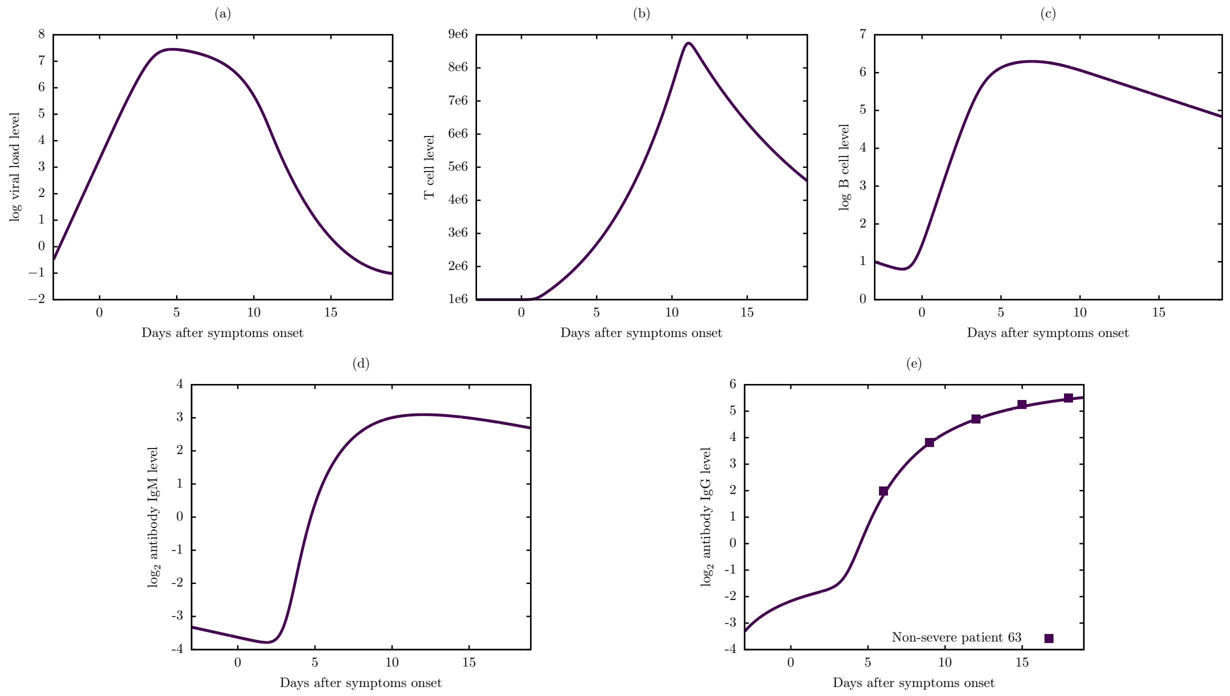


**Figure S39:** Viral dynamic and immune response of Model 6 for non-severe patient 63.

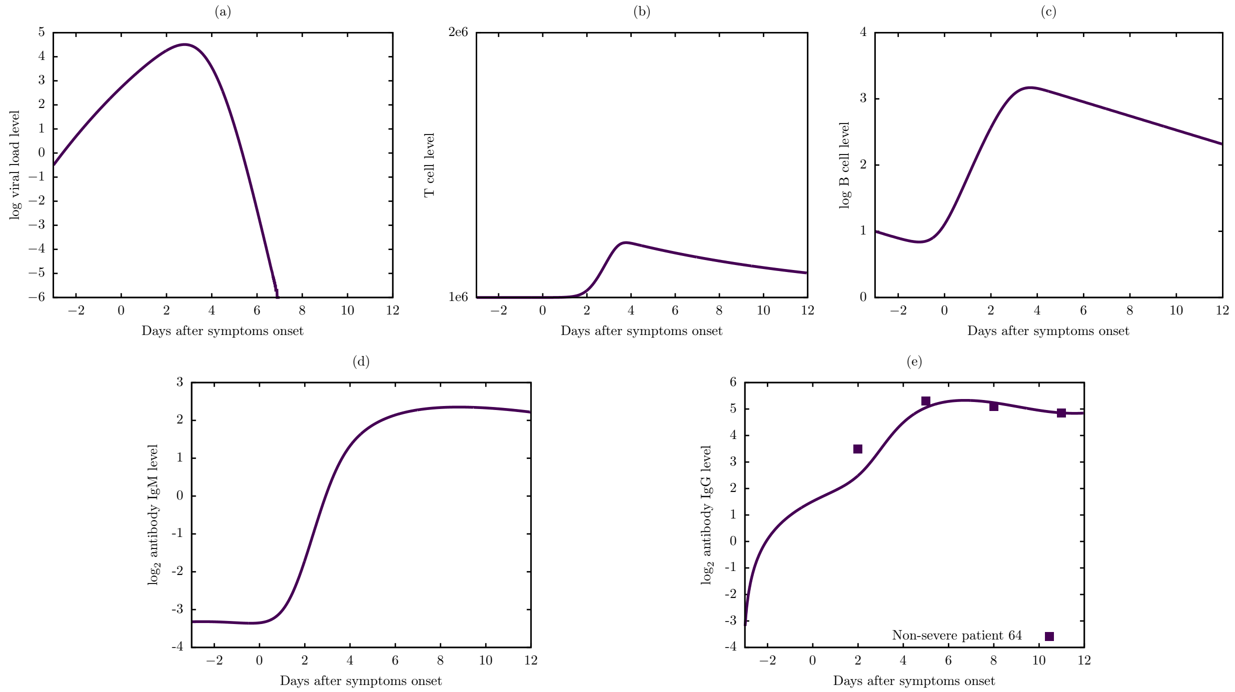


**Figure S40:** Viral dynamic and immune response of Model 6 for non-severe patient 64.
